# Assessing the Impact of Timing and Coverage of United States COVID-19 Vaccination Campaigns: A Multi-Model Approach

**DOI:** 10.64898/2026.04.07.26349269

**Authors:** Anjalika Nande, Soren L Larsen, James Turtle, Jessica T. Davis, Shraddha Ramdas Bandekar, Bryan Lewis, Shi Chen, Lucie Contamin, Sung-mok Jung, Emily Howerton, Katriona Shea, Clara Bay, Michal Ben-Nun, Kaiming Bi, Anass Bouchnita, Jiangzhuo Chen, Matteo Chinazzi, Spencer J. Fox, Alison L. Hill, Harry Hochheiser, Joseph C. Lemaitre, Sara L. Loo, Madhav Marathe, Lauren Ancel Meyers, Carl A. B. Pearson, Przemyslaw Porebski, Emily Przykucki, Claire P. Smith, Srinivasan Venkatramanan, Alessandro Vespignani, Timothy C. Willard, Katie Yan, Cecile Viboud, Justin Lessler, Shaun Truelove

**Affiliations:** Johns Hopkins University, Baltimore, Maryland, USA; University of Oxford, Oxford, UK; University of Illinois, Urbana Champaign, Urbana, Illinois, USA; University of California, Berkeley, Berkeley, California, USA; Predictive Science Inc., San Diego, California, USA; Northeastern University, Boston, Massachusetts, USA; University of Texas at Austin, Austin, Texas, USA; University of Virginia, Charlottesville, Virginia, USA; University of North Carolina at Charlotte, Charlotte, North Carolina, USA; University of Pittsburgh, Pittsburgh, Pennsylvania, USA; National University of Singapore, Singapore, Singapore; Princeton University, Princeton, New Jersey, USA; The Pennsylvania State University, University Park, Pennsylvania, USA; The University of Texas Health Science Center at Houston, Houston, Texas, USA; University of Texas at El Paso, El Paso, Texas, USA; Northern Arizona University, Flagstaff, Arizona, USA; University of North Carolina at Chapel Hill, Chapel Hill, North Carolina, USA; National Institutes of Health Fogarty International Center, Bethesda, Maryland, USA

**Keywords:** ‘COVID-19’, ‘Scenario modeling’, ‘Ensemble projections’, ‘Vaccination timing and coverage strategies’, ‘Seasonal dynamics of respiratory infections’, ‘Mathematical modeling’, ‘Multi-model aggregation’

## Abstract

Six years after its emergence, SARS-CoV-2 continues to have a substantial burden, however, the impact of vaccination and the optimal timing of its rollout remain uncertain. To explore these uncertainties, the US Scenario Modeling Hub convened its 19th round of ensemble projections for COVID-19 hospitalizations and deaths in the United States. Eight teams provided outcomes for each US state and nationally from April 2025 to April 2026 under five scenarios regarding vaccine recommendations and timing. We assessed recommendations with two eligibility scenarios (high-risk individuals only and all-eligible) and two timing scenarios (classic start: mid-August, earlier start: late June). These were crossed to create four scenarios and were compared against a counterfactual scenario with no vaccination. We found that compared to no vaccination, our ensemble projections estimated 90,000 (95% PI 53,000-126,000) hospitalizations averted in the high-risk and classic timing scenario across the US. Expanding coverage averted an additional 26,000 (95% PI 14,000-39,000) hospitalizations, which when coupled with earlier vaccination timing further reduced national hospitalizations by 15,000 (95% PI −3,000-33,000). These findings estimate significant benefits from a broad all-eligible vaccination recommendation, and suggest an additional benefit is likely to be gained from an earlier vaccination campaign.

## Introduction

Six years after the first reported case of COVID-19 in the United States,^1^ SARS-CoV-2 remains a substantial source of disease burden, with an estimated 390,000 to 550,000 hospitalizations due to COVID-19 in the US 2024-25 respiratory season.^2^ During this period, policy questions have evolved alongside disease dynamics and variants of concern,^3^ and in particular, the impact of vaccination campaigns at the population level has been an area of continued exploration.^4,5,6^ Substantial heterogeneity across age in transmission^7^ and risk of severe disease ^8^ has necessitated evaluation of vaccine prioritization and eligibility, for example in the context of resource limitations when vaccines were first approved,^9^ and later on, expansion of vaccine approval to younger age groups.^10^ Several factors have the potential to influence COVID-19 booster impacts – and consequently, vaccine policy recommendations – in future years. These include that the majority of the US population is no longer immunologically naive,^11^ the propensity to get vaccinated has decreased, the increased prevalence of hybrid immunity^11,12^ may decrease the rate of immune waning,^13^ and the landscape of variants continues to evolve^14^. Further, for the past several years, both a summer and winter wave have been observed, in contrast to influenza and RSV dynamics, which only show a winter season wave.^15^ In light of these considerations, the population impacts of vaccine campaigns in 2025-26, and subsequent seasons remains uncertain. To navigate these uncertainties and support evidence-based policy decisions, scenario modelling can compare alternative futures and enable projections informing vaccine policy decisions under various epidemiological and immunological assumptions.

Since December 2020, the US Scenario Modeling Hub (SMH) has generated 18 rounds of ensemble-based projections at the state and national level to inform COVID-19 policy.^16,17^ These projections are ensembled by aggregating outcomes from multiple mathematical models, solicited in an open call.^18^ This approach has been shown to enhance the reliability of predictions, compared to those from individual models.^19^ For each round, SMH defined a set of scenarios in collaboration with decision makers, aimed at answering specific questions for policy or about critical epidemiological uncertainty; these have included vaccine effectiveness, vaccination coverage, timing of vaccine rollout, waning immunity, emergence and characteristics of new variants, and non-pharmaceutical interventions.

This study presents the results from SMH’s 19th round of COVID-19 projections conducted in early 2025, examining five scenarios (A-E) across a 52-week period for the 2025-26 season (April 27, 2025 to April 25, 2026) that vary vaccination campaign timing and target population coverage (Table 1). Unlike seasonal influenza and RSV, since 2022, seasons of COVID-19 in the US have been characterized by two annual waves, making vaccination campaign planning and evaluation challenging. While winter peaks remain common, substantial summer activity has been observed in the last three years, potentially driven by a combination of climate factors, behavioral responses, waning immunity, and changes in susceptibility driven by viral evolution.^20,21, 22^ To account for this dual-wave pattern, SMH evaluated the potential impact of starting the vaccination campaign 1.5 months earlier than in previous years, alongside different coverage recommendations, on projected COVID-19 hospitalizations and deaths. These scenario projections are not intended as forecasts or to predict exact epidemic target outcomes, but rather to explore plausible epidemic trajectories under alternative vaccination strategies.

**Table 1.** COVID-19 projection scenario definitions for Round 19 of the US Scenario Modeling Hub, 27 April 2025–25 April 2026. Scenario specifications focus on the impact of different target group recommendations for updated COVID-19 booster vaccinations (rows) combined with different timing of booster vaccination (columns) for the 2025-2026 season. Modelling teams were instructed to provide projections for hospitalizations and deaths in 50 states, D.C., and at the national level under these five scenarios.

|  | Vaccine recommended, same timing of immunization as last year campaign | Vaccine recommended, early timing of immunization campaign |
| --- | --- | --- |
| **No vaccine recommendation** | **Scenario A**<br>No new vaccinations during 2025-26 season |  |
| **Annual vaccination recommended for high-risk groups (65+ and those with underlying risk factors)** | **Scenario B**<br>• Vaccination starts 15 August 2025<br>• Vaccine has 45% VE against hospitalization at time of initial roll-out<br>• Uptake in high risk groups (individuals 65+ or with underlying risk factors) is the same as seen for 2024-25<br>• Uptake in all other groups is negligible | **Scenario C**<br>• Vaccination starts 29 June 2025 (1.5 month earlier than in 2024-25)<br>• Vaccine has 45% VE against hospitalization at time of initial roll-out<br>• Uptake in high risk groups (individuals 65+ or with underlying risk factors) is the same as seen for 2024-25<br>• Uptake in all other groups is negligible |
| **Annual vaccination recommended for all currently eligible groups** | **Scenario D**<br>• Vaccination starts 15 August 2025<br>• Vaccine has 45% VE against hospitalization at time of initial roll-out<br>• Coverage saturates at levels of the 2024-25 booster for all population | **Scenario E**<br>• Vaccination starts 29 June 2025 (1.5 months earlier than in 2024-25)<br>• Vaccine has 45% VE against hospitalization at time of initial roll-out<br>• Coverage saturates at levels of the |
|  | subgroups (approximately 22% of adults nationally, 44% of 65+, 26% of high risk individuals 18-64 yrs) | 2024-25 booster for all population subgroups (approximately 22% of adults nationally, 44% of 65+, 26% of high risk individuals 18-64 yrs) |

## Results

### Projected national-level burden of hospitalizations and deaths

The national-level hospitalization ensemble of eight models projected two distinct periods of increased COVID-19 activity: a summer wave peaking in August 2025 and a winter wave peaking in January 2026 (Figure 1A), with similar timings projected for peak deaths (Figure S2). Based on the 50% projection interval, both peaks were expected to approach those observed during the previous winter season (2024–2025), peaking at approximately 20,000 weekly hospitalizations nationally. While projections for the winter wave were relatively consistent across models, uncertainty around the summer wave size was greater, with two of eight participating models projecting insubstantial summer activity (Figure S3). At the national level, there is limited variance across scenarios. Peak timing is largely consistent, however the peak magnitude and cumulative burden varies slightly depending on the vaccine schedule and coverage (Figure 1C,D). State level projections were more heterogeneous, with some states projected to experience much larger winter waves (e.g., California, Hawaii) and others to have larger summer waves (e.g., Pennsylvania, Ohio) (Figure S4).

**Figure 1.**
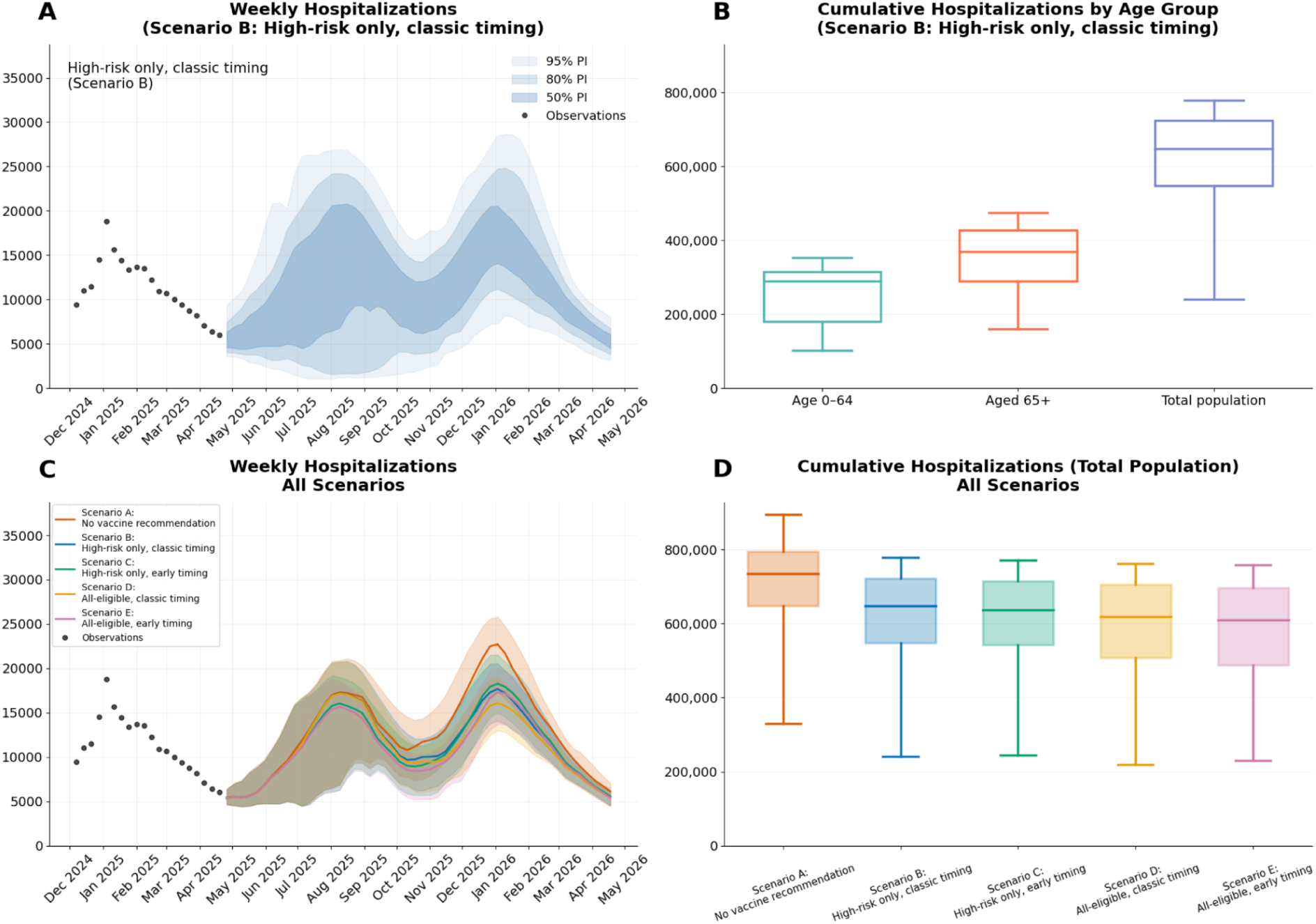
National-level ensemble predictions of COVID-19 hospitalizations in the US, 27 April 2025–25 April 2026. A) Projected incident hospitalizations across Scenario B (high risk group vaccination and classic timing). Shaded regions represent the 50%, 80%, 95% projection intervals. B) Projected cumulative hospitalizations by age-group and total population of Scenario B. The outer boxes represent the IQR and the whiskers are the 95% projection interval. C) Projected incidence of hospitalizations across all scenarios. Shaded regions represent the 50% projection interval and the solid line represents the median value. D) Projected cumulative hospitalizations for all scenarios. The outer boxes represent the IQR and the whiskers are the 95% projection interval.

Projections indicated substantial continuing COVID-19 hospitalization and death burdens, with an estimated 648,000 cumulative hospitalizations (95% prediction interval [PI] 241,000–778,000) and 40,000 deaths (95% PI 25,000-59,000) during the 2025-2026 season under Scenario B (vaccination recommended for high-risk individuals only, with classic timing consistent with the previous season; Figures 1B, S2F). The majority of the burden was projected among adults aged 65 years and older, accounting for 57% of hospitalizations and 81% of deaths.

### Impact of vaccination coverage and timing

Across both national and state levels, cumulative pooled results showed that vaccination recommendations, whether limited to high-risk groups or extended to all eligible individuals, were projected to substantially reduce hospitalizations and deaths compared to the counterfactual Scenario A with no vaccination recommendation, regardless of campaign timing (Figure 2A). Relative to Scenario A, recommending vaccination for high-risk groups (Scenario B) was projected to reduce cumulative national hospitalizations by 13% (95% confidence interval [CI] 8-17%) and deaths by 16% (95% CI 11-21%) over the projection period with classic vaccination timing. This corresponds to approximately 90,000 (95% CI 53,000–126,000) fewer hospitalizations and 7,000 (95% CI 5,000-9,000) fewer deaths nationally.

**Figure 2.**
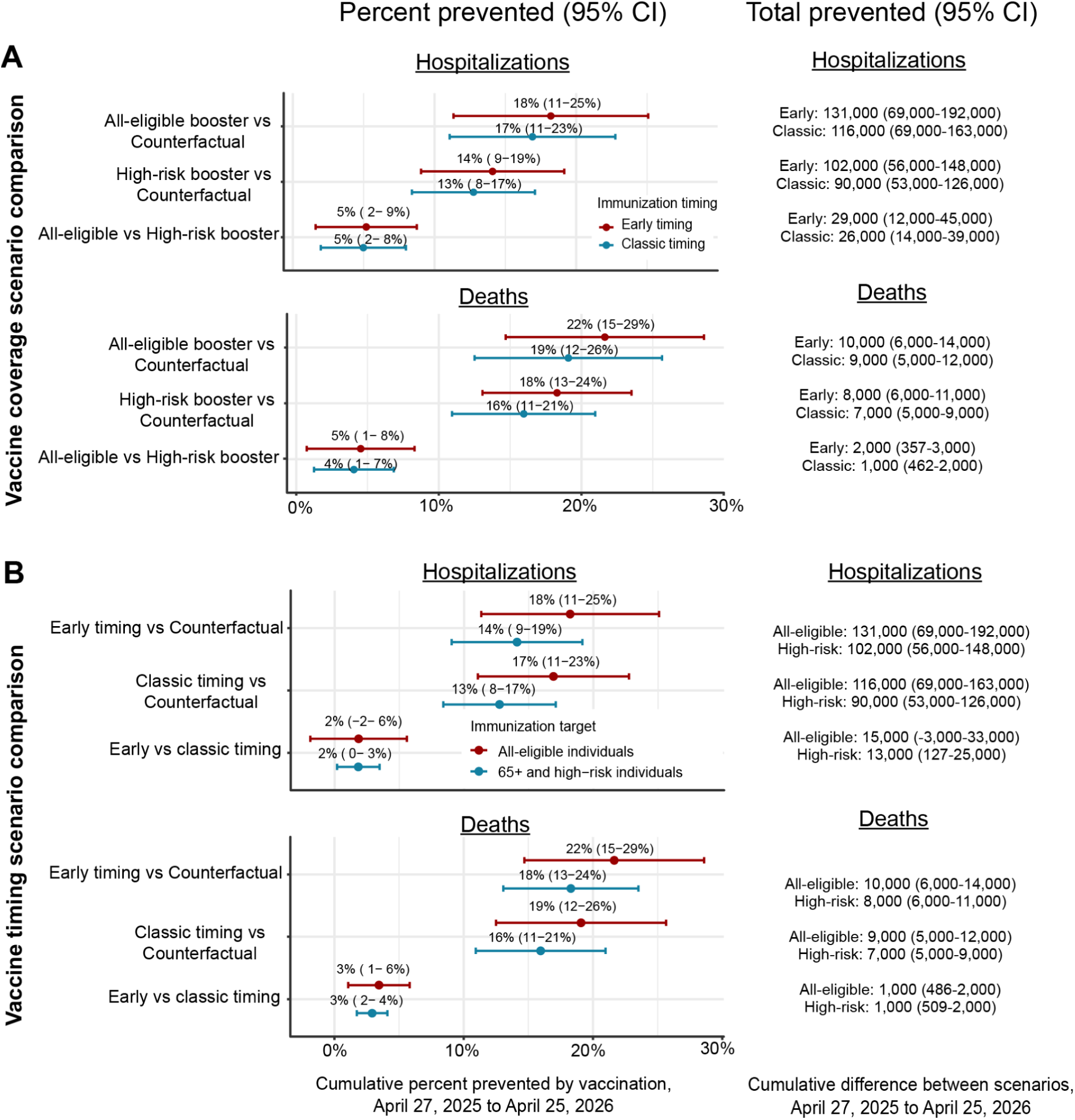
Projected national-level impacts of COVID-19 vaccination recommendation and timing scenarios in the United States, 27 April 2025 to 25 April 2026. A) Projected percentages and values of hospitalizations and deaths averted due to either all-eligible or high-risk only COVID-19 booster vaccination recommendations, under either early or classic timing assumptions. B) Projected percentages and values of hospitalizations and deaths averted due to early (starting 29 June) or classic (starting 15 Aug) COVID-19 vaccination timing, across the projection period among all age-groups, nationally. These projections suggest significant value of continued COVID-19 booster vaccinations, for both high-risk and all-eligible recommendations, as well as potentially significant impacts of shifting the vaccination campaign earlier.

Extending vaccination recommendations to all eligible groups (Scenario D) was projected to avert about 116,000 (95% CI 69,000-163,000) hospitalizations and 9,000 (95% CI 5,000-12,000) deaths, preventing an additional 26,000 hospitalizations and 2,000 deaths compared with the high-risk–only recommendation. Advancing the start of vaccination campaigns by 1.5 months (Scenarios C and E) was projected to additionally reduce hospitalizations by 2% (95% CI −2-6%) and deaths by 3% (95% CI 1-6%) relative to the classic timing (Figure 2B).

Most of the vaccination benefits were projected to occur among adults aged ≥65 years, the result of both direct reductions in outcomes and indirect effects through decreased transmission. In this group, Scenario B was projected to avert 67,000 (95% CI 38,000-96,000) hospitalizations and 6,000 (95% CI 4,000-7,000) deaths compared with Scenario A (Figure S5). Despite equal vaccine coverage among those aged ≥65 years across scenarios, all-eligible vaccination recommendations (Scenario D) were projected to avert an additional 12,000 (95% CI 6,000-18,000) hospitalizations and 1,000 (95% CI 395-2,000) deaths among this group, reflecting significant indirect benefits of vaccinating the full population. While these patterns are consistent across models, individual model estimates varied considerably, reflecting differences in model assumptions on seasonality, vaccine effectiveness, and waning immunity (Figures S6-S7).

### Relationship between vaccination timing, epidemic timing and vaccination benefits

Projected national-level epidemic timing of individual model trajectories was characterized by the week at which 50% of hospitalizations were reached—which we refer to as the median hospitalization week (MHW; Figure 3A). Focusing on the all-eligible vaccination coverage and classic timing scenario (Scenario D), the majority of MHWs fell earlier than EW 43 (week of 2025-10-19) for four out of eight models, while two models exhibited trajectory timings concentrated between EW 44-51, and the densities for the remaining two models were concentrated during EW 50 and later. This heterogeneity persisted across all scenarios (Figure S8), therefore all timing interaction analyses were broken out by model—rather than taking results from the ensemble as a whole. In addition to MHW, trajectory timings were classified by which wave was larger: summer, winter, or similar (see Supplementary Results 2A and Figures S9, S10).

**Figure 3.**
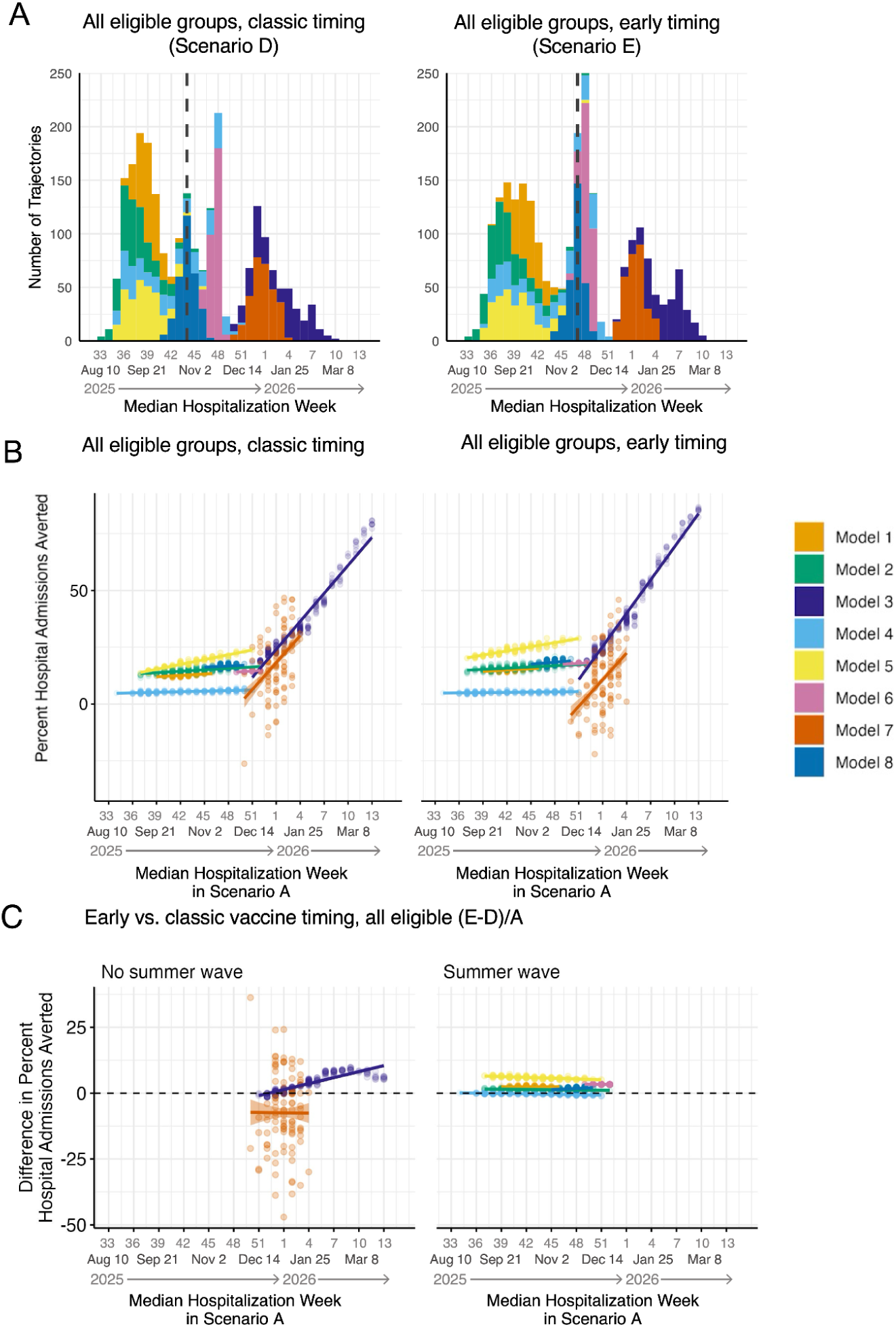
Relationship between projected COVID-19 trajectory timing, vaccination timing, and estimated vaccine benefits, 27 April 2025 to 25 April 2026, United States. A) Distribution of the timing of COVID-19 hospitalizations for Scenario D (left) and Scenario E (right), as characterized by the median hospitalization week (MHW), and colored by model. The median MHW across all models is indicated by a vertical dashed line. B) Percent hospitalizations averted in Scenarios D (all eligible, classic vaccine timing) and E (all eligible, earlier vaccine timing), relative to the counterfactual Scenario A, vs. trajectory mean hospitalization week (MHW) in Scenario A. C) The difference in percent averted between early timing (Scenario E) and classic timing (Scenario D) vs trajectory MHW, calculated as 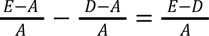. Models have been grouped into those with a summer wave (right subpanel), and those without (left subpanel). Solid lines correspond to results of linear regression; coefficients are provided in Tables S5-S10. Dashed lines denote the threshold above (below) which early (classic) timing is better. Note: under classic vaccine timing, at least 50% of the final vaccination coverage has been attained for all age groups by epidemic week 46.

Earlier vaccination timing relative to epidemic timing has the potential to reduce COVID-19 burden, but the actual impacts of a 1.5-months earlier start time were not conclusive. First, when regressing across all paired trajectories and models, we observed a consistent positive relationship between later trajectory timing and greater relative vaccine impacts, regardless of scenario (Figure S11). Later trajectory timing also appeared to increase the relative effect of 1.5-months earlier vaccine timing overall, though this association was nonmonotonic (Figure S12). However, because each model covers only a portion of the MHW space (Figure 3A), it was challenging to analyze the interaction of epidemic timing and vaccination timing in the ensemble. To address this issue, within-model timing trends were analyzed. When looking at continuous trajectory timings at national (Figure 3B) and sub-national (Figures S13,14) levels, the percent hospitalizations averted was consistently higher with later epidemic timing across all models and scenarios.. Comparing the percent averted by early versus classic vaccination timing in scenarios with the same vaccination recommendation groups (C and B, D and E), earlier vaccination timing averted more hospitalizations regardless of trajectory timing for six out of eight models across national-level (Figure 3C) trajectories and five out of eight models across state-level (Figures S15,16) trajectories. However, models do not agree on how trajectory timing affects the relative hospitalizations averted (in the classic timing versus early timing scenarios). There was a consistent positive association in two out of eight models across national trajectories, and three out of eight models across state-level trajectories. This means that later epidemic timing would increase the advantage of early over classic vaccine timing in these cases. Subnationally, the percent of hospitalizations averted was positively associated with MHW across 98.2% of location, model, and scenario combinations (Figure S17). In 66.2% of location-model combinations, later epidemic timing was associated with reduced effectiveness of early vaccination against classic timing scenarios, but the strength and direction of this association was largely model dependent (Figure S18). Note, these trends were largely preserved when considering a discrete characterization of trajectory timings (summer-dominant, winter-dominant, and similar) instead (Suppl. Results, Figures S19-S21).

### Effects of vaccination scenario on summer and winter waves

We investigated the effect of the vaccination scenarios on the summer and winter wave sizes in the ensemble (Figure 4) and the association between them for each model and location (Supplementary Methods 1D, Results 2C, Figure S22). Comparing the two scenario axes, vaccination timing had a larger effect on the summer wave size compared to vaccination coverage level (Figure 4B): earlier vaccination averted 5.30 (per 100,000) more summer hospitalizations (Scenario C vs B) on average, whereas increased vaccination coverage (Scenario D vs B) averted only 0.63 (per 100,000) more hospitalizations. In contrast, vaccination coverage had a greater impact on the winter wave size (Figure 4C): increased vaccination averted 7.52 more winter hospitalizations per 100,000 (Scenario D vs B) on average, while earlier vaccination increased winter hospitalizations by 1.64 per 100,000, on average (Scenario C vs B). Overall, expanded eligibility and earlier timing increased averted hospitalizations by 12.95 per 100,000, on average (Scenario E vs B) over the entire outbreak (Figure 4A). Full results of the analysis are provided in Table S11. These trends were maintained when the analysis was repeated for each model individually, separating out locations by size of the summer and winter waves, or by historical vaccination coverage levels (Figures S23-S26, Tables S12-S22 and Supplementary Results 2D).

**Figure 4.**
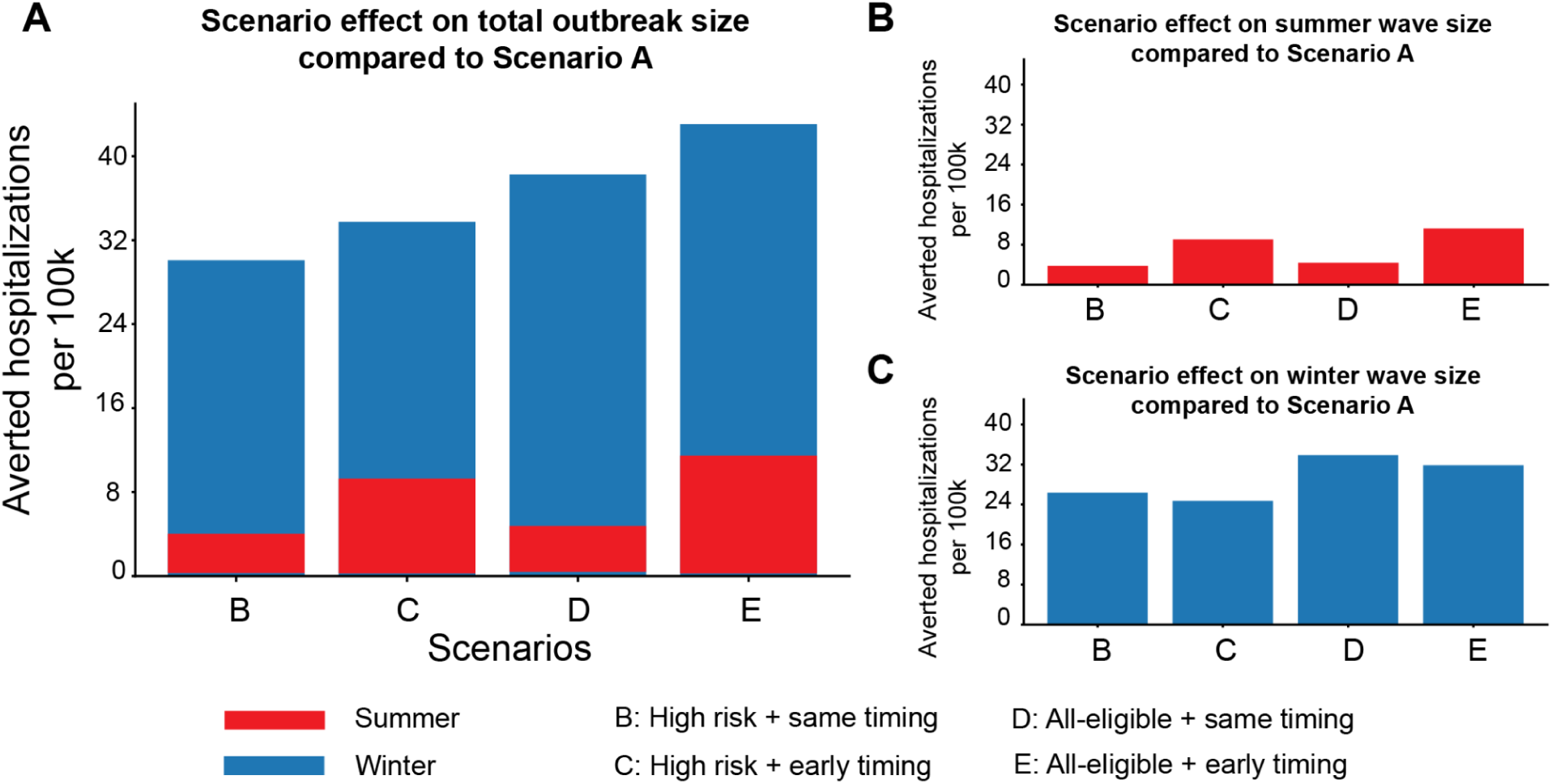
Averted COVID-19 hospitalizations under the vaccination scenarios compared to the no vaccination Scenario A, 27 April 2025 to 25 April 2026, United States. Mean vaccination effect, defined as the number of hospitalizations averted per 100,000, on the A) total, B) summer and C) winter wave sizes for the ensemble model with scenarios as the fixed effect and states and teams as the random effects. The effect on the total outbreak is divided into effects on the summer (red) and winter waves (blue). Comparison between scenarios indicates that vaccination timing has a larger effect on the summer wave, whereas vaccination coverage has a larger effect on the winter wave. Across all scenarios, vaccination has a larger effect on the winter wave compared to the summer wave.

## Discussion

In the nineteenth round of US Scenario Modeling Hub COVID-19 projections, we found the need and value for continued vaccination to curb COVID-19 burdens (hospitalizations and deaths) during the 2025-2026 season. Further, we found novel results that indicate earlier vaccination timing is likely to provide additional population-level benefit. Specifically, vaccination recommendations for all eligible groups and an earlier vaccination campaign combination was the most impactful in averting COVID-19 hospitalizations (18%, 95% CI: 11-25%) and deaths (22%, 95% CI: 15-29%), compared to the counterfactual scenario without vaccination recommendations. While these effects varied substantially across models, the diversity of model structures, approaches, and assumptions allowed us to further explore questions about vaccine timing, epidemic timing, and the drivers of summer vs. winter wave sizes.

While segregation of epidemic timings across individual models hindered ensemble analysis of the relationship between epidemic timing and vaccine impacts, analysis of within-model trends yielded multiple key insights. First, there were overwhelmingly positive associations between later MHW and higher percent of averted hospitalizations across individual models, scenarios, and spatial scales, suggesting strong evidence of modest benefits from earlier vaccination relative to epidemic timing, presumably because more individuals were immunized by the time of high epidemic activity. While not directly observed, there is likely a point of diminishing returns when an early vaccination campaign (i.e., 2.5-mo earlier than classic timing) is too early. Second, early vaccination timing generally reduced COVID-19 burden more than classic vaccination timing, but the relationship between the scale of these impacts and the timing of the epidemic was inconsistent and inconclusive. Moreover, we find optimal vaccination timing may fall outside the range of the discrete vaccine timing scenarios, indicated by the larger trend line slope for models with greater period between vaccination and epidemic wave (i.e., Figure 3C). This suggests the need for a more granular and broader vaccination timing axis to effectively capture this optimum at both national and state levels. A follow up study with greater vaccination timing variability, including in the context of the immune landscape and waning of immunity, could further explore this tradeoff, and provide more granular evidence to support optimal timing of vaccination campaigns at the state level.

Despite model variabilities, effects of vaccination scenarios on the summer and winter wave sizes were qualitatively similar across individual models, with more averted hospitalizations in states dominated by winter waves. Additionally, we found that the summer wave is more responsive to an earlier vaccination campaign than to increased vaccination coverage, the result of shifting new vaccinations to time periods with high infection burden. In contrast, the winter wave benefits more from increased vaccination coverage than an earlier campaign. These results provide new insights into optimal vaccination strategies to tackle the strong biannual seasonality of COVID-19 burdens^15^ across different regions and should be considered for maximum impact of vaccination, particularly given variability of summer/winter burden across the US. It is also worth noting that while this work focuses on COVID-19 in the US, the underlying framework and insights are more broadly applicable as COVID-19 in other countries ^23^ as well as many other respiratory pathogens ^24–26^ exhibit spatiotemporal heterogeneity in transmission and burden, and would benefit from tailored vaccination strategies rather than a single uniform schedule.

Our work has several limitations. First, our scenario design doesn’t consider the effects of increasing vaccine uptake. Scenario E specifies the highest national coverage based on historical data, 42.8% in high-risk individuals and 24.6% overall (Table S2). While higher uptake would likely reduce acute disease the most, changing vaccine hesitancy is often quite difficult and therefore was not a focus.

Second, our ensemble model has high uncertainty on the projected summer wave size. This is partially due to the lack of robust calibration data available for summer and fall 2024, during which NHSN hospitalization reporting was paused^27^, and variability in how individual models handled this issue.

Despite these uncertainties, national projections accurately captured the seasonal distribution of reported hospitalizations in 2025–2026 (Figure S2), lending support to our conclusion regarding vaccination timing. Although we cannot directly evaluate the accuracy of these scenario projections due to reality not perfectly matching any single scenario, our projections appear to be somewhat inflated. This overestimation is possibly a result of changes not accounted for in models, including reductions in reporting and continued declines in susceptibility and severity, as COVID-19 transitions to endemicity. Fortunately, this overestimation should primarily affect estimates of absolute vaccination benefits, and relative effects (percent averted) should be less sensitive to these issues. Third, we remain in the early stages of understanding the mechanisms of COVID-19 biannual seasonality. There is substantial spatial heterogeneity in the summer wave across the US, including in timing and intensity.^22^ A more comprehensive characterization of this biannual seasonality is needed to design and implement more effective COVID-19 interventions, such as precise vaccination timing and coverage for different locations. Finally, our projections share the common limitation of long-term models that overlook continual viral evolution where emerging variants or subvariants with distinct epidemiological characteristics may exhibit changing dynamics.

Given rapid viral evolution, SARS-CoV-2 is likely to continue to cause substantial disease burden, including hospitalizations and deaths in the US and around the world. In this work we found that the 2025-26 season, and possibly future seasons, would likely continue to be characterized by two waves nationally, in summer and winter, though the exact drivers of these dynamics remain uncertain. With this biannual burden, COVID-19 presents an important question for policy and public health: should we shift away from the traditional fall vaccination campaign timing, and when would be the ideal timing to maximize the value of the COVID-19 vaccine? While the evidence from this work points to earlier vaccination campaigns in locations that experience greater relative burden of disease in the summer, further investigation is needed. This work highlights the importance of empirically-relevant modelling efforts that can provide essential insight to assess and guide policy, potentially helping to maximize the benefits of declining resources like vaccination.

## Methods

Eight teams contributed weekly projections of COVID-19 hospitalizations and deaths by state and nationally during a 52-week projection horizon from 27 April 2025 to 25 April 2026; projections and aggregated results were released in June 2025. Teams calibrated independent models using weekly COVID-19 hospitalizations and deaths from the National Healthcare Safety Network (NHSN)^27^ and National Centre for Health Statistics (NCHS),^28^ with calibration specifics (e.g., timeframe for calibration or inclusion of additional data sources) at the teams’ discretion ensuring that no data from the projection period was used, and submitted 150-300 trajectories (e.g., randomly sampling from a range of parameter values or stochastic iterations) per target, scenario, location, and age-group.

### Trajectory pairs

To ensure comparability between scenarios, teams were required to ‘pair’ trajectories across scenarios, targets, horizons and age groups. Pairing dictates that all model parameters outside of the scenario assumptions remain the same, such that an individual trajectory in a scenario is comparable to its paired trajectory in the other scenarios. The pairing requirement plus a counterfactual Scenario A (no vaccination recommendation) enables estimation of the number and percent of averted hospitalizations and deaths.

### Ensemble methodology

Individual team’s model projections were combined to produce ensemble projections using a trimmed linear opinion pool method,^29^ and estimates of burden averted under different mitigating scenarios were obtained using meta-analysis approaches.^6^ For both hospitalizations and deaths, eight models contributed to the ensemble.

### Scenario design

Five scenarios (Table 1) were considered across two axes: vaccination group recommendations and timing of the vaccination campaign. For vaccination groups, three levels were considered: i) no recommendations; ii) recommendations for high-risk group only, including individuals over 65 years and those under 65 years with pre-existing medical conditions, such as chronic obstructive pulmonary disease (COPD), diabetes, chronic kidney diseases;^30^ and iii) universal recommendation for all eligible individuals across age groups. Vaccination uptake was set to 2024-25 coverage levels for vaccination groups in each scenario (Figure S1). For vaccination timing, we considered two start times for all locations: i) classic timing, with the vaccine roll-out aligned with the 2024-25 campaign (i.e., 15 August 2025), and ii) an earlier timing, where the vaccination campaign would start 1.5 months earlier (i.e., 29 June 2025). In all scenarios, updated vaccines were expected to match the predominant SARS-CoV-2 variants circulating on 30 June 2025. The vaccine effectiveness (VE) against COVID-19 hospitalization was assumed to be 45% at the start of the vaccination campaign, consistent with a recent analysis of US COVID-19 hospitalizations.^31^ VE against infection and death and the pace of immune escape were left to the scientific discretion of the teams.^32^ We did not consider the impact of a new variant with rapid, significant antigenic change, nor substantial increase in transmissibility (akin to Delta or Omicron variants). Severity of future circulating variants in naive populations was assumed to remain similar to the Omicron lineages.

### Trajectory timing analysis

To understand the relationship between epidemic and vaccination timings, and how it affects COVID-19 burden–for example, understanding whether the combination of an earlier epidemic and vaccination campaign leads to lower burden–we characterized timings of individual model projections at the level of the trajectories. Trajectory timings were quantified in two ways: 1) continuous - median hospitalization week (MHW, defined as the first epidemic week reaching ≥50% cumulative hospitalizations) and 2) categorical - using the percentage of cumulative hospitalizations occurring by epidemic week (EW) 43, as on average, it is the midpoint between summer and winter waves. Models (model number: 4, 5-8) that provided fewer than 300 trajectories were upsampled for a consistent comparison across all models (Supplementary Methods 1C). Trajectories were defined as “summer-dominant” if more than 55% of cumulative hospitalizations occurred by EW 43, “winter-dominant” if fewer than 45%, or “similar” if between 45-55% accrued. Frequency of trajectory timing classifications at both national and state levels were analyzed. Then, using Scenario A as the reference group, we systematically quantified the percent hospitalization averted in Scenarios B through E. We also computed the percent of averted hospitalizations in earlier vs. regular vaccine timing in scenarios with the same vaccine eligibility criteria (i.e., Scenario C vs. B, and D vs. E). We focus on hospitalizations for this analysis as deaths are highly correlated with it.

### Analysis of vaccination scenario effects on summer and winter waves

We also investigated the effect of the vaccination scenarios on the summer and winter wave sizes. A mixed-effects model was fitted to the cumulative ensemble estimates of the total, summer and winter wave sizes, with scenarios as a fixed effect and locations (i.e., states) and teams as a random effect. The coefficients of the fixed effects (i.e., the vaccination scenarios) correspond to the number of hospitalizations averted per 100,000 population relative to the reference Scenario A. Supplementary analyses were conducted at the level of individual models to control for model-specific effects, state specific effects related to historically higher summer or winter outbreaks, and variation in vaccination coverage (Supplementary Methods 1E,F).

All analyses were performed in *Python* and *R* and the analysis code is freely available at: https://github.com/midas-network/covid19-scenario-hub_r19_manuscript. Authors used ChatGPT and Claude to assist with writing some of the analysis code. After using these tools, the authors reviewed and edited the content as needed and take full accountability for the final content.

## Author contributions

CV, EH, HH, JL, KS, KY, SJ, and ST contributed to scenario concept and design; AB, ALH, AN, AV, BL, CB, CABP, CPS, CV, JC, JTD, JCL, JT, KB, LC, LAM, MB, MC, PP, SRB, SC, SJF, SL, SLL, ST, and SV contributed to acquisition, curation, analysis, or interpretation of data; AB, ALH, AN, AV, BL, CB, CABP, CPS, JC, JTD, JCL, JT, KB, LAM, MB, MC, MM, PP, SRB, SC, SJF, SL, SLL, ST, and SV contributed to model design, calibration, or analysis; AN, BL, EH, JC, JTD, JL, JT, KS, LC, PP, SRB, SJ, SL, and ST contributed to ensemble post-processing, analysis, or interpretation; AN, BL, JTD, JT, SC, SB, SL, and ST drafted the manuscript; AB, ALH, AN, AV, BL, CB, CABP, CPS, CV, EH, HH, JC, JTD, JL, JCL, JT, KB, KS, KY, LC, LAM, MB, MC, MM, PP, SRB, SC, SJF, SJ, SL, SLL, ST, and SV contributed to manuscript critical review; ALH, BL, HH, JL, KS, LAM, MB, MM, SJF, and ST obtained funding; CV, EP, HH, JL, KY, MM, SJ, ST, and TCW contributed to administrative, technical, or material support; ALH, AN, BL, HH, JL, JT, MM, and ST provided supervision.

## Data Availability

All data used in this work is available online at: https://github.com/midas-network/covid19-scenario-modeling-hub. And all the analysis code is available at: https://github.com/midas-network/covid19-scenario-hub_r19_manuscript

## Acknowledgments

We extend our gratitude to the Center for Forecasting and Outbreak Analytics-Scenario Modeling Group for their valuable contribution in generating and submitting model results to this effort.

## Sources of Funding and Support

ALH, AN, CABP, EP, JCL, TCW, SLL, LC, HH, JL, ST was supported by the CDC-CFA award for the Atlantic Coast Center for Infectious Disease Dynamics and Analytics (ACCIDDA):

NU38FT000012-01-00 (UNC).

HH and LC were supported by NIH Grants U24GM132013 and R24GM153920.

KY and KS were supported by NSF RAPID awards DEB-2126278, and DEB-2220903, and KY was also supported by NSF Grant No. DGE1255832. MC acknowledges support from CDC-JHU-2005702123. EH was supported by federal funds from the National Cancer Institute, National Institutes of Health, under Prime Contract No. 75N91019D00024, Task Order No. 75N91023F00016. The content of this publication does not necessarily reflect the views or policies of the Department of Health and Human Services, nor does mention of trade names, commercial products or organizations imply endorsement by the U.S. Government.

PP, SV, BL, JC, MM acknowledge support from SMH Supplement via UNC ACCIDDA CDC-CFA: NU38FT000012-02-02, SMC Fellowship 75D30121F00005-2005604290, NSF Grant No. OAC-1916805,

NSF Expeditions in Computing Grant CCF-1918656, DTRA subcontract/ARA S-D00189-15-TO-01-UVA, and UVA strategic funds. This material is based upon work supported by the NAVAL SEA SYSTEMS COMMAND under Contract No. N00024-22-D-6404 JT, MBN were supported by a subcontract of the CDC-CFA award for the Atlantic Coast Center for Infectious Disease Dynamics and Analytics (ACCIDDA): NU38FT000012-01-00 (UNC).

SL was supported by the National Institute of General Medical Sciences of the National Institutes of Health under award number R35GM156856.

SRB and LAM were supported by CDC-CFA award: NU38FT000008 and CDC award U01IP001136.

## Supplementary Materials

### 1. Supplementary Methods

#### 1A. Modeling Team Descriptions

A total of eight models submitted to this scenario modeling challenge (Table S1). The types of models ranged from mechanistic compartmental approaches to hybrid Fourier transformation and compartmental approaches. While the scenario design required the same assumptions about vaccine timing, coverage, and efficacy against hospitalization, modeling teams had different assumptions around population structure, natural history, immunity against infection, seasonality, and calibration process. Full details on model descriptions and assumptions can be found here: https://docs.google.com/spreadsheets/d/12VFgTtOoyTN806E_I6q1ufUX-vXv07MvdVcmXFtAjBY/edit?gid=0#gid=0

**Table S1:** Table mapping the description of the models to the labels used in the analysis in the main text.

| Model Label | Model Name | Institution | Model Type | Reference |
| --- | --- | --- | --- | --- |
| Model 1 | CFA-Scenarios | CDC CFA | Meta-population, compartmental model |  |
| Model 2 | JHU_UNC-flepiMoP | Johns Hopkins University and University of North Carolina at Chapel Hill | Mechanistic age-specific compartmental model | 1 |
| Model 3 | MOBS_NEU-GLEAM_COVID | Northeastern University | Meta-population, age-specific, stochastic compartmental model | 2 |
| Model 4 | PSI-M_CoV_2025 | Predictive Sciences | Mechanistic age-specific compartmental model |  |
| Model 5 | UIUC-IMMCYC | University of Illinois - Urbana-Champaign | Mechanistic age-stratified compartmental model | 3 |
| Model 6 | UNCC-Hierbin | University of North Carolina - Charlotte | Fourier transformation and compartmental hybrid model | 4 |
| Model 7 | UT-ImmunoSEIRS | University of Texas at Austin | Mechanistic age-stratified compartmental model | 5 |
| Model 8 | UVA-adaptive | University of Virginia | Meta-population, compartmental model | 6 |

#### 1B. Quantifying national vaccination coverages

In this study, we consider scenarios with three different levels of vaccination: no vaccination (counterfactual - scenario A), high-risk (scenarios B and C), and all-eligible (scenarios D and E). In addition to scenario and location differences, vaccine uptake time-series are stratified by age group and high/low risk populations (see Figure S1). Table S2 summarizes these differences by aggregating total doses (millions) across locations and age groups. Percent population coverage appears in parenthesis. The counterfactual scenario A is excluded from the table because it specifies no doses administered.

**Table S2:** National vaccine doses (millions) by scenario and risk population. For each scenario specification, millions of doses are listed for each risk group. Additionally, the risk-population percentage uptake appears in parentheses.

| Scenario | Low Risk | High Risk | Totals |
| --- | --- | --- | --- |
| B - high risk, classic timing | 0 (0.0%) | 38.5 (39.8%) | 38.5 (11.8%) |
| C - high risk, early timing | 0 (0.0%) | 41.4 (42.8%) | 41.4 (12.7%) |
| D - all eligible, classic timing | 35.4 (15.4%) | 38.5 (39.8%) | 73.9 (22.7%) |
| E - all eligible, early timing | 38.8 (16.9%) | 41.4 (42.8%) | 80.3 (24.6%) |

#### 1C. Upsampling model trajectories for timing analysis

The scenario specification stated that teams must submit 150 to 300 trajectories for each location and scenario. As a result, some teams submitted 150 trajectories, and some submitted 300. For the analysis of trajectory timing by model, we upsampled such that each team had equal representation in the distribution of trajectory timings. Because we only had two trajectory counts (150 and 300), upsampling was implemented as a simple duplication of trajectories from models with a 150 count.

#### 1D. Association between the sizes of the summer and winter waves

To investigate the association between winter and summer wave sizes, we split each trajectory at EW 43 for each model, location and scenario. Summer wave size was calculated as the cumulative number of hospitalizations that occurred until the end of EW 42. Similarly, winter wave size was the cumulative number of hospitalizations from EW 43 through the end of the simulation horizon. As an exploratory analysis of the biannual seasonality of COVID-19 hospitalizations, associations between the summer and winter wave sizes for each team, location, and scenario were determined by the Pearson’s correlation coefficient.

#### 1E. Mixed effects analysis per model for summer and winter leaning locations

We investigated how sizes of summer and winter waves differed across scenarios. A location was tagged as summer-leaning or winter-leaning based on which wave had more than 50% of cumulative hospitalizations on average across all the Scenario A trajectories. A mixed-effects model was fitted separately to summer and winter leaning locations, with scenarios as a fixed effect and locations (i.e., states) as a random effect. The ‘effect’ was the difference in number of hospitalizations per 100,000 population relative to the reference Scenario A. We report the number of hospitalizations averted per 100,000 relative to Scenario A.

#### 1F. Division of states into low, medium and high coverage

In our scenario analysis, the baseline vaccination coverage level for each state was assumed to equal that of the 2024-2025 vaccination campaign. Given that there was significant variation in the baseline coverage levels ranging from less than 10% of a state receiving the booster vaccine to greater that 35% (Figure S24), for each team, we repeated the mixed-effects analysis separately for states grouped into low, medium, and high vaccination coverage. This grouping was based on the historical coverage levels and states were divided into tertiles (so that each group contained one-third of the locations). Note, as in the main text analysis, states were also further divided into the summer and winter dominant categories resulting in six separate mixed-effects models that were fit for each team for each variable of interest (for e.g., the total epidemic size) using Scenario A as baseline, the other scenarios as fixed effects, and the particular subset of states as a random effect (Figures S25-S26).

### 2. Supplementary Results

#### 2A. Characterization of trajectory timing

To characterize the timings of model trajectories we used median hospitalization week (MHW) and a seasonality classification where trajectories are classified as having a stronger summer wave, stronger winter wave, or similarly sized waves. Applying the same methods to national observed hospitalization data for the previous four seasons yields the results in Table S3. Reporting was voluntary—and significantly lower—during a long period of 2024, so the true timing of the 2024-25 season is likely earlier than the observed timings. However—ignoring the 2024-25 season—we see MHWs varying from epidemic week 40 to 48, and one season each classified as Summer, Similar, and Winter.

**Table S3:** National NHSN observed MHW for confirmed COVID-19 hospital admissions. *NHSN reporting was not mandatory from May 1, 2024 (EW 18) until October 30, 2024 (EW 48). The metrics presented here for 2024-25 are useful for comparison to the observed data, but likely not a good characterization of the actual season timing.

| Season | MHW | Seasonality |
| --- | --- | --- |
| 2021-22 | 46 | Similar |
| 2022-23 | 40 | Summer |
| 2023-24 | 48 | Winter |
| 2024-25* | 51 | Winter |

In the main text we generally avoided drawing timing-related conclusions from the ensemble, but it is interesting to note how scenarios affected trajectory timing. Table S4 contains the median MHW across all models for national projections, by scenario. Scenarios with early vaccine timing (C and E) reduced the summer wave, resulting in a slightly later MHW. The scenario with no vaccination (A) results in larger winter waves, and thus later MHW.

**Table S4:** National ensemble median trajectory MHW for each scenario.

| Scenario | Median MHW |
| --- | --- |
| A | 47 |
| B | 45 |
| C | 46 |
| D | 44 |
| E | 47 |

When classifying national trajectory timings by summer, similar, and winter, all scenarios produced more winter-than summer-dominant ensemble trajectories (Figure S9). However, the proportion of winter-dominant trajectories was substantially lower in the classic (B: 42% and D: 40%) vs early (C: 50% and E: 50%) vaccination scenarios. Winter-dominant trajectories were also more prevalent than summer-dominant at the subnational level (Figure S10). Looking specifically at the counterfactual (Scenario A), 44 of 51 locations exhibited a higher proportion of winter-dominant trajectories.

#### 2B. Categorical epidemic timing analysis

This section contains extended results using categorization of epidemic timings. At the national level, Figure S19 illustrates percent hospitalization averted for each model and scenario, grouped by epidemic timings: summer-dominant, winter-dominant, and similar. Looking more directly at the effect of vaccine timing, Figure S20 depicts the difference between percent hospitalizations averted for early and classic vaccination timing with one panel each for high-risk (scenario C minus scenario B) and all-eligible (scenario E minus scenario D) vaccine strategies. For each panel, 14 of 16 model-timing combinations show an increase in percent averted in earlier vaccination timing scenarios. Four out of five models with coverage across more than one timing group show a positive median averted-difference for earlier trajectory timings.

Extending the analysis to state-level, Figure S21 compares the impact of vaccine timing vs categorized epidemic timing for the full ensemble of models. The difference in percent hospitalizations averted (early minus classic vaccine timing) is shown for each subnational location. The top panel takes the difference of the two high-risk vaccine scenarios (C-B), and the bottom panel compares the all-eligible scenarios (E-D). In almost every location-timing combination the median outcome results in more hospitalizations averted under the early vaccine scenario. For high-risk vaccination (top panel), median outcomes are positive for 49 (50, 51) of 51 locations for summer (similar, winter) epidemic timings. For all-eligible vaccination (bottom panel), median outcomes are positive for 49 (49, 51) of 51 locations for summer (similar, winter) epidemic timings.

#### 2C. Association between summer and winter wave sizes

The association between summer and winter wave sizes was assessed using Pearson’s correlation coefficient *r* for each model at national and state levels for each scenario (Figure S22). Here, summer wave size corresponds to the number of hospitalizations up to (including) EW 42 for each trajectory and winter wave size is the number of hospitalizations from EW 43 till the end of the projection period. Three out of eight models (2, 3, and 6) reported a predominantly positive association between the two waves, i.e., a large summer wave implies a large winter wave, although variability exists at the state level. Sizes of the two waves were predominantly negatively associated for the other five models. These patterns are consistent across all five scenarios.

We speculate this variability in the sizes of the summer and winter wave results from a complicated interplay between the force of infection (primarily driven by intrinsic viral transmissibility and seasonality) and immune dynamics, for example, waning of prior infection- or vaccine-induced immunity. While the exact contribution of each mechanism is difficult to disentangle, as models implement them differently, these results suggest immune dynamics likely drive the winter wave for those models with a predominantly negative association. This could be the result of a large summer wave depleting the susceptible population, leading to a smaller successive winter wave. Wave sizes can be explained by the force of infection for models with a positive association between the two waves, with both waves either high or low depending upon intrinsic viral transmissibility and/or assumptions about immunity waning in the model.

#### 2D. Vaccination scenario effects on hospitalizations per team

For each team’s projections, a mixed effects model (with the scenarios as a fixed effect and the state as a random effect) was deployed to investigate vaccination scenario effects on total, summer and winter wave sizes, with Scenario A (no vaccination) as the reference (Figure S23). These effects were calculated separately for states with historically larger summer or winter waves (see Suppl. Methods 1E). Vaccination effect on total epidemic size was larger for winter leaning states, driven by larger reductions in winter wave size across scenarios and models, except Model 4. There were 13.78 (average range across scenarios: 11.91-15.78) more hospitalizations per 100,000 during the winter waves in winter leaning states than summer leaning ones. Scenario effects on summer wave size were similar for summer and winter leaning states, with an average 2.34 (range: 1.35-3.72) more averted hospitalizations per 100k in the winter leaning states. A similar analysis was also done where states were instead divided by historical vaccine coverage levels (see Figure S24 and Suppl. Methods 1F) and the qualitative trends were unaffected (Figures S25-26).

### 3. Supplementary Figures

**Figure S1.**
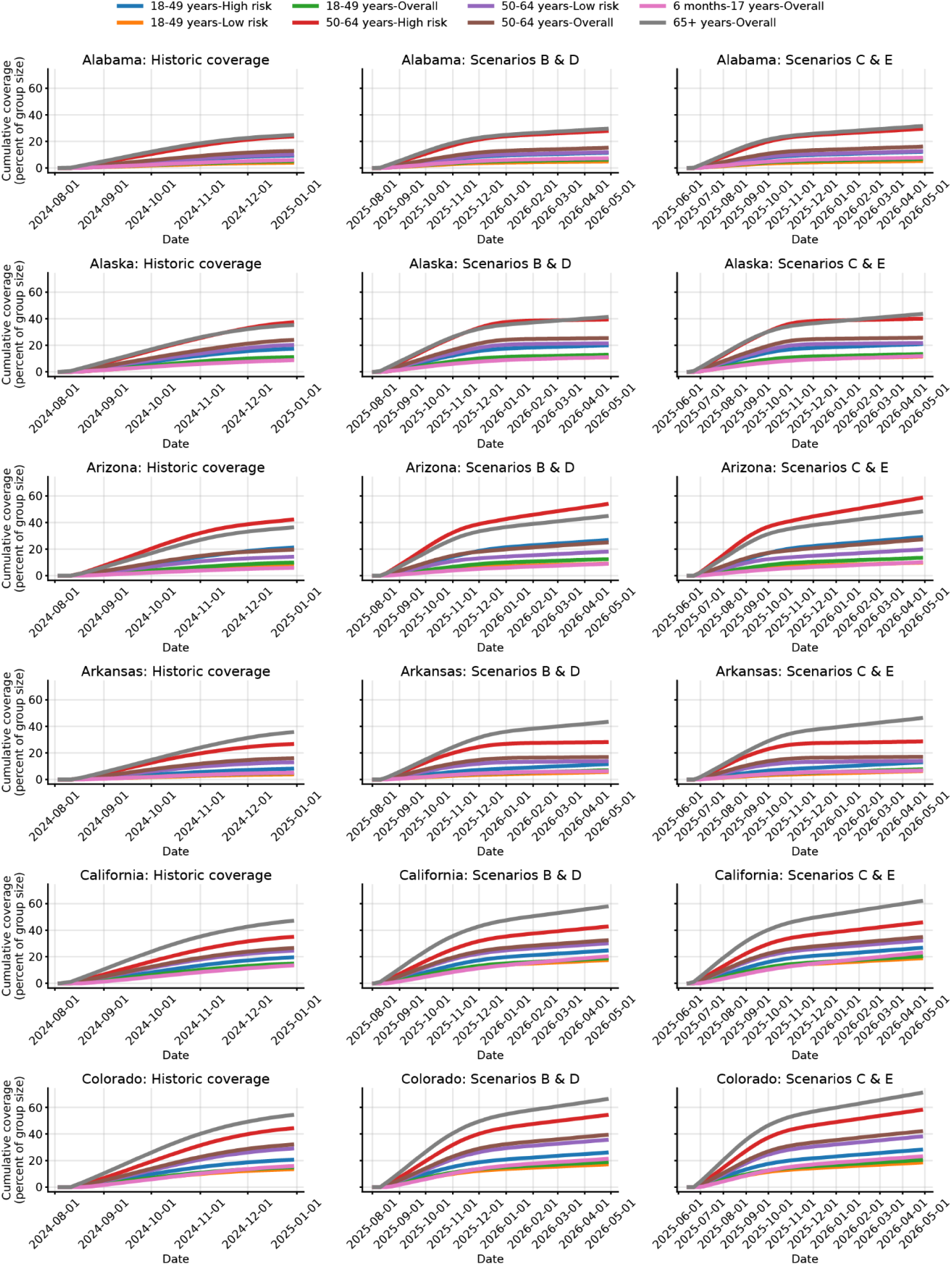
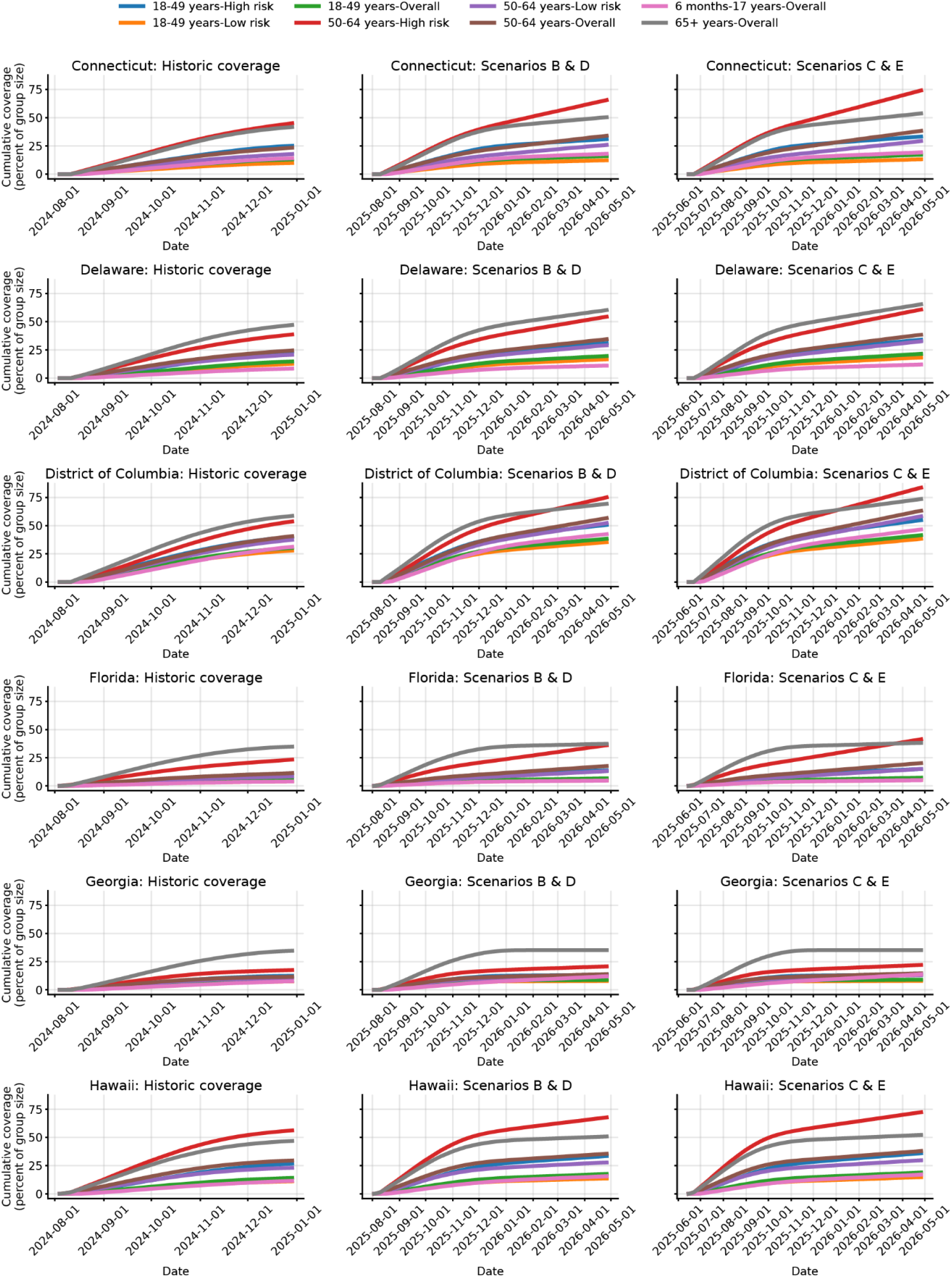
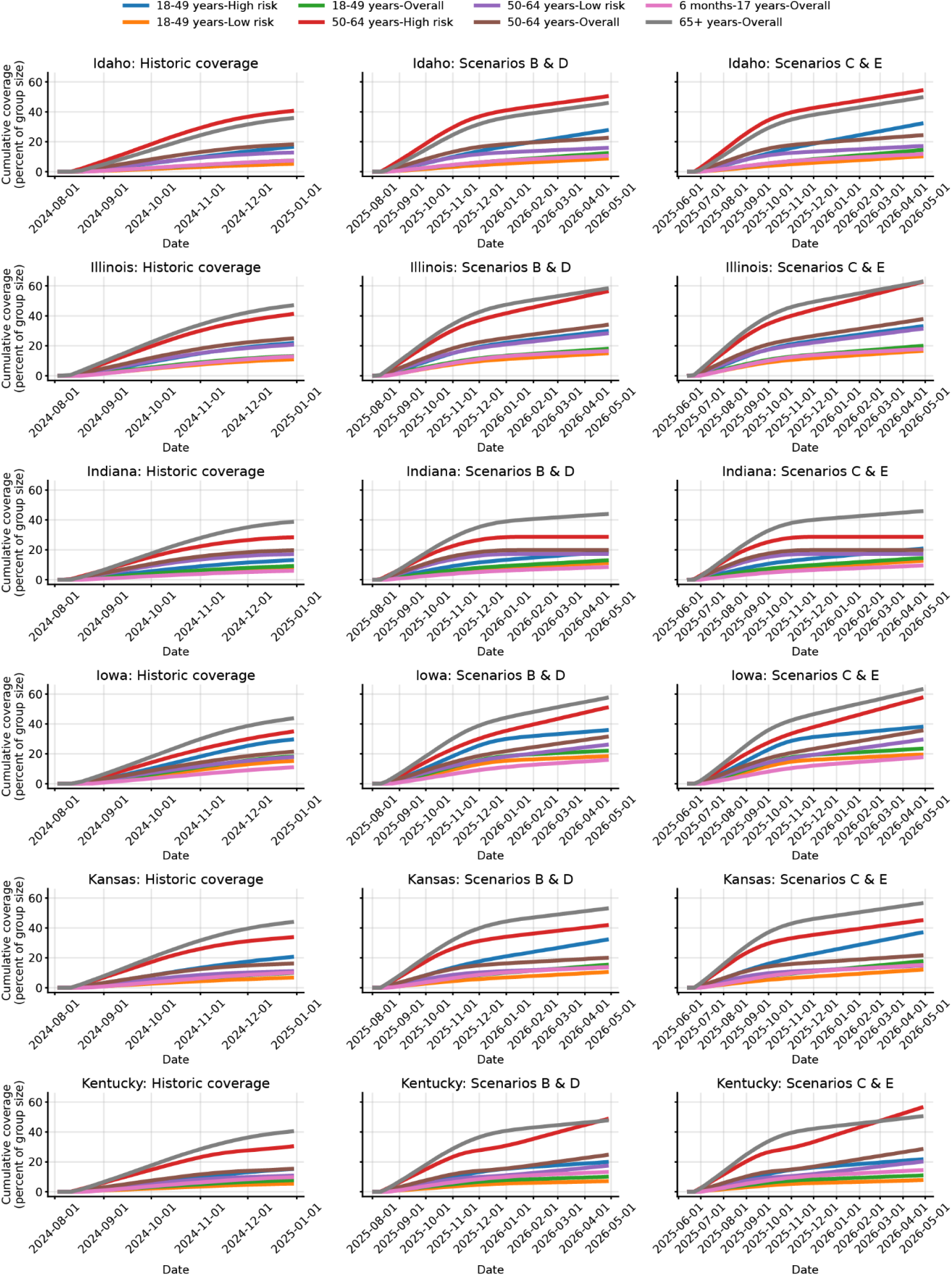
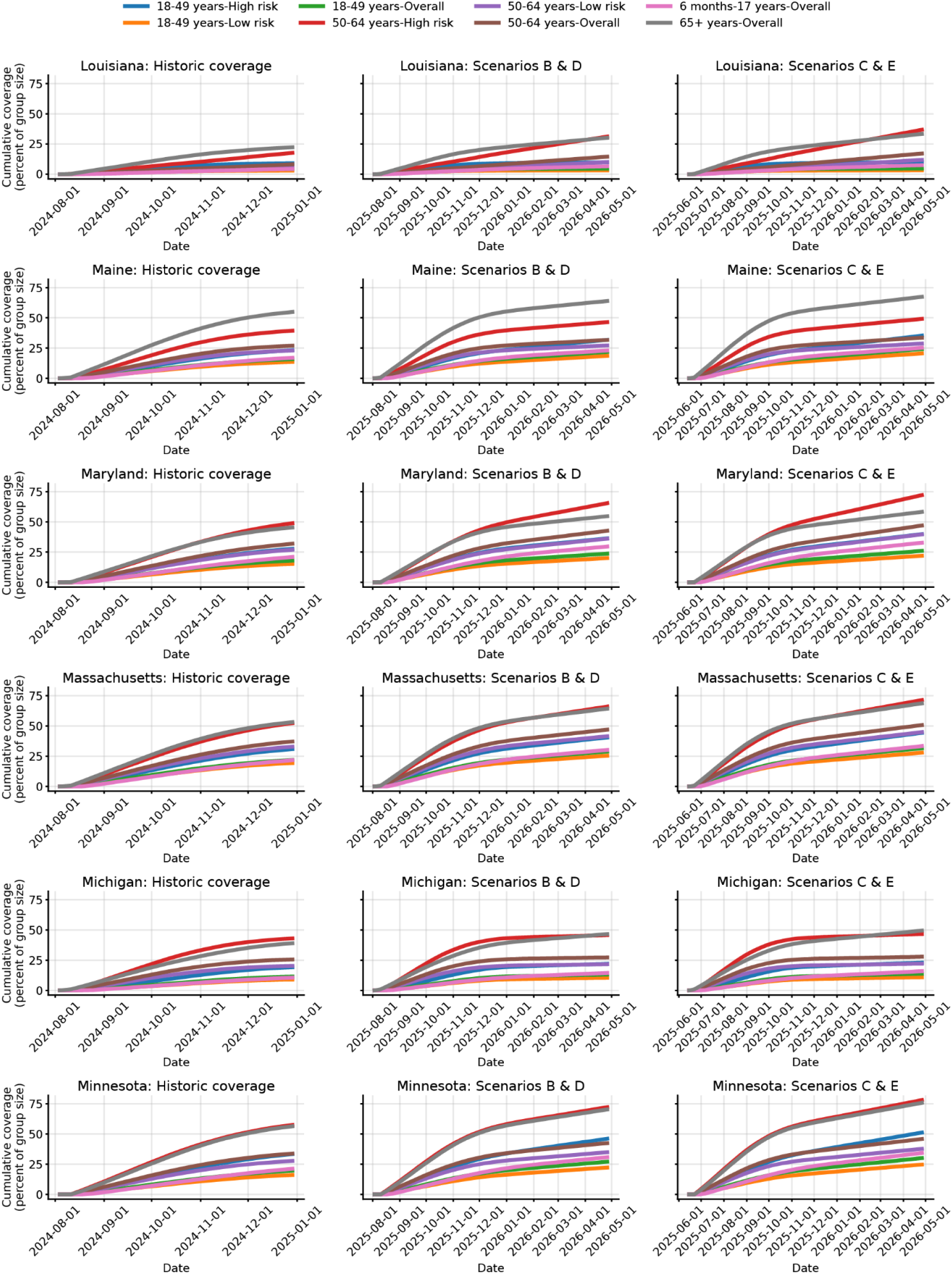
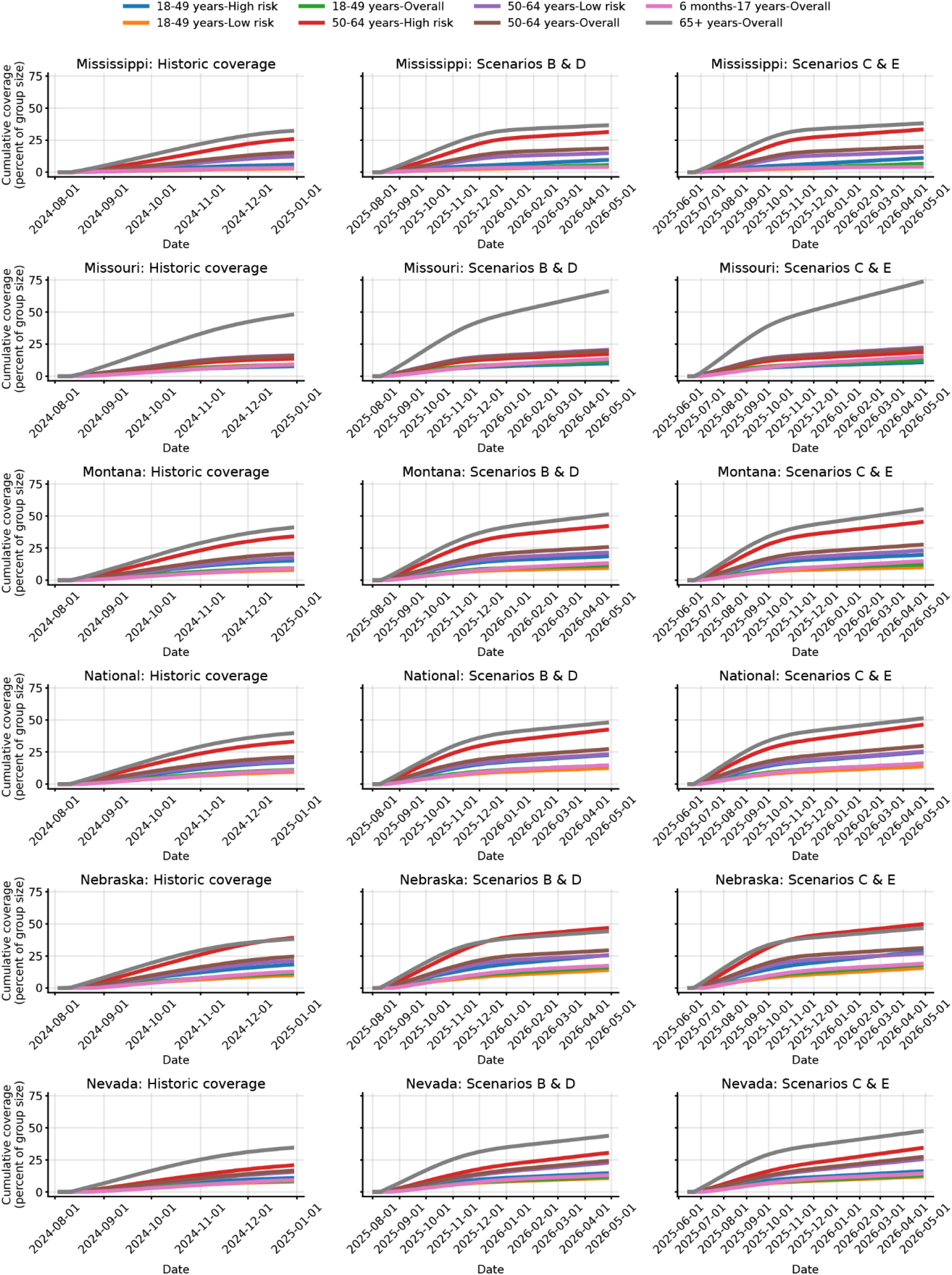
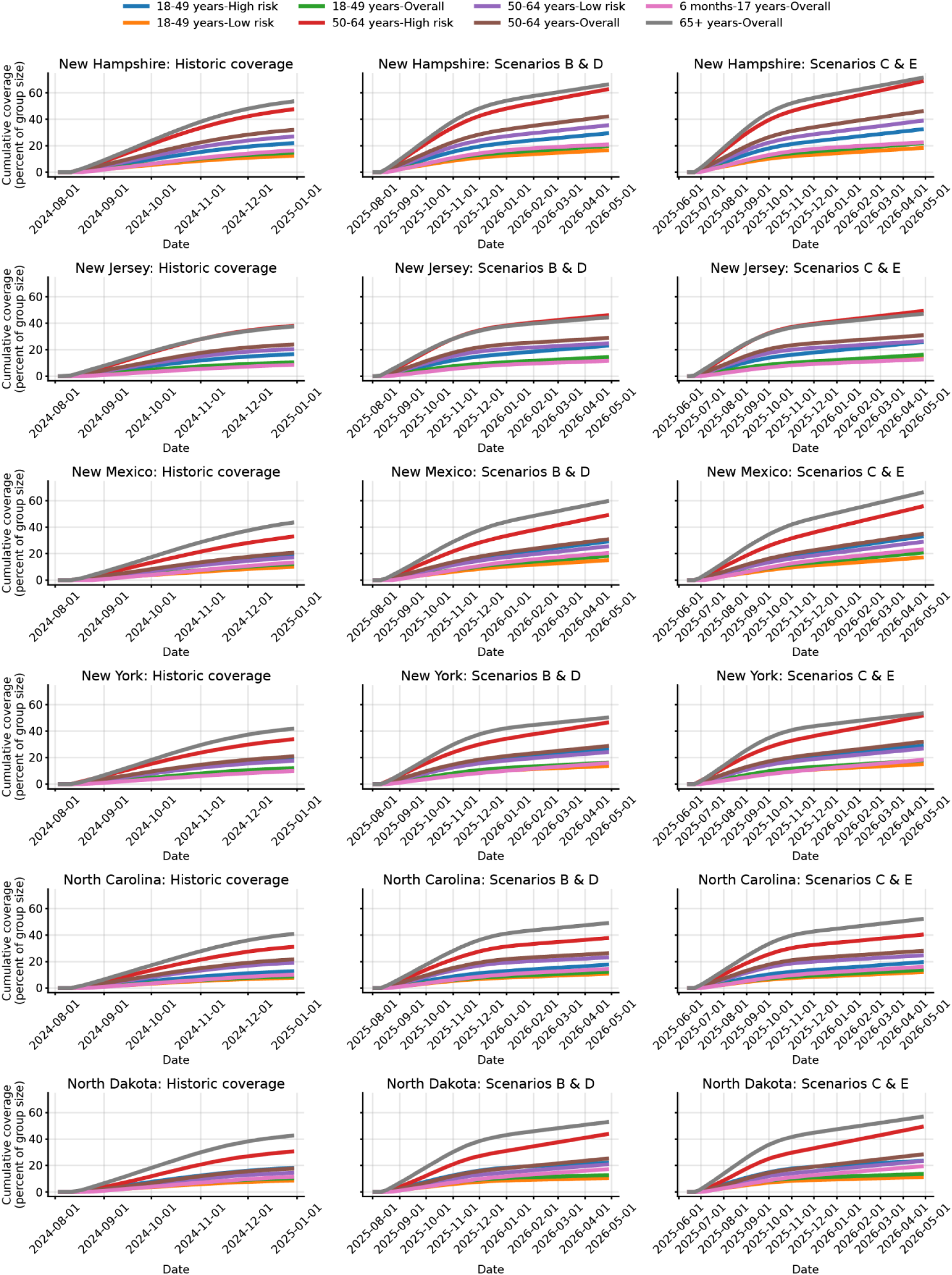
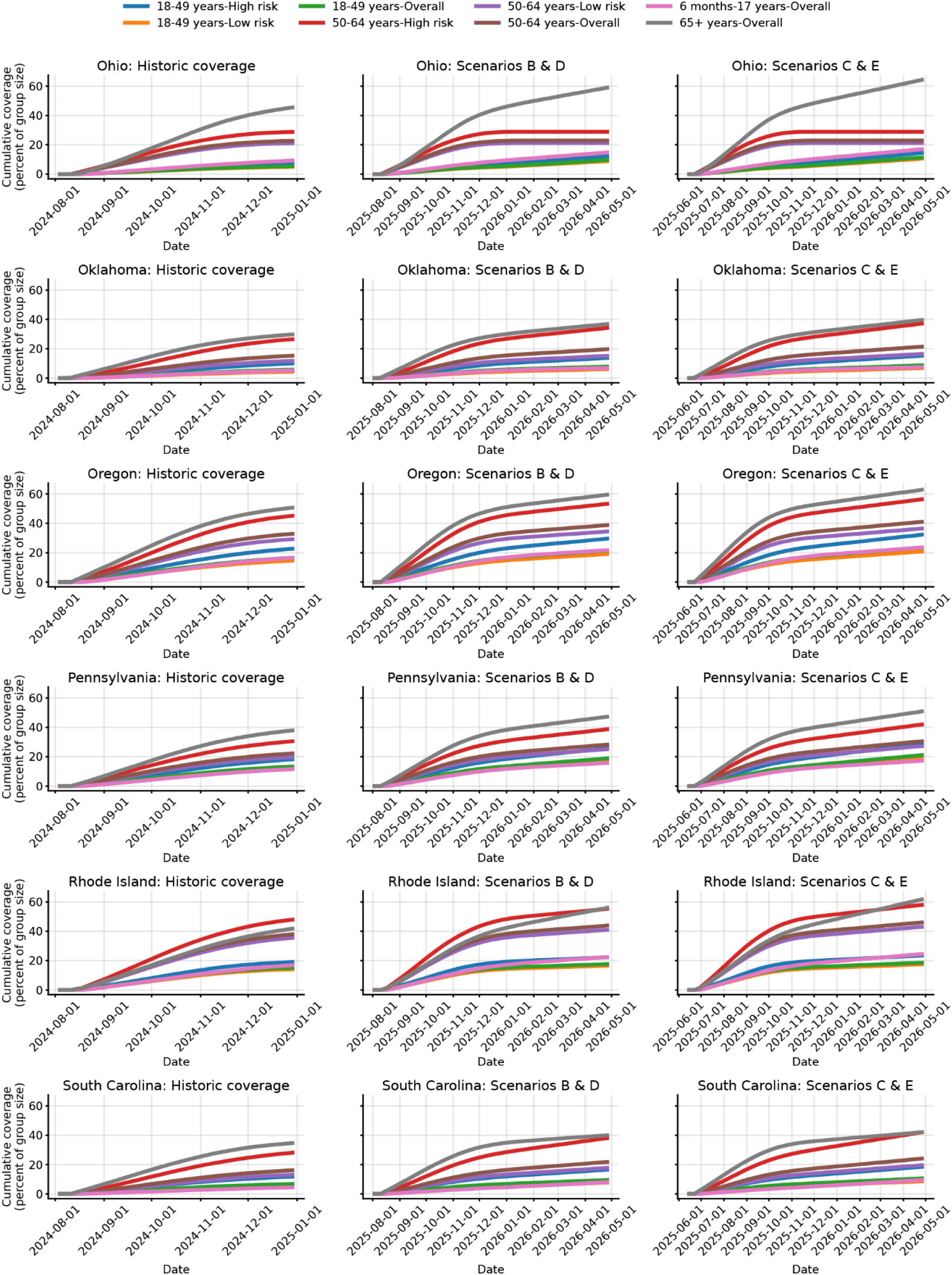
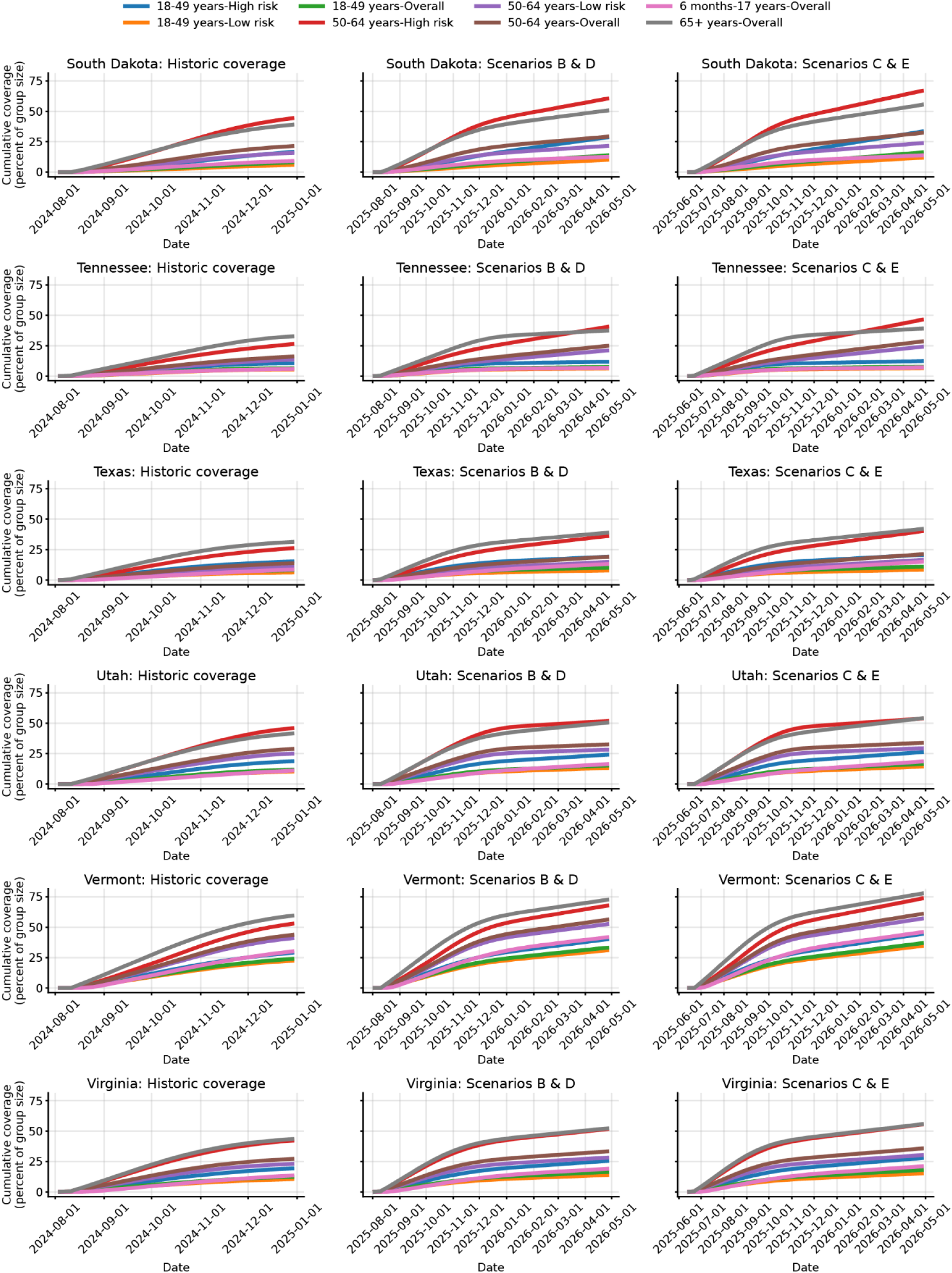
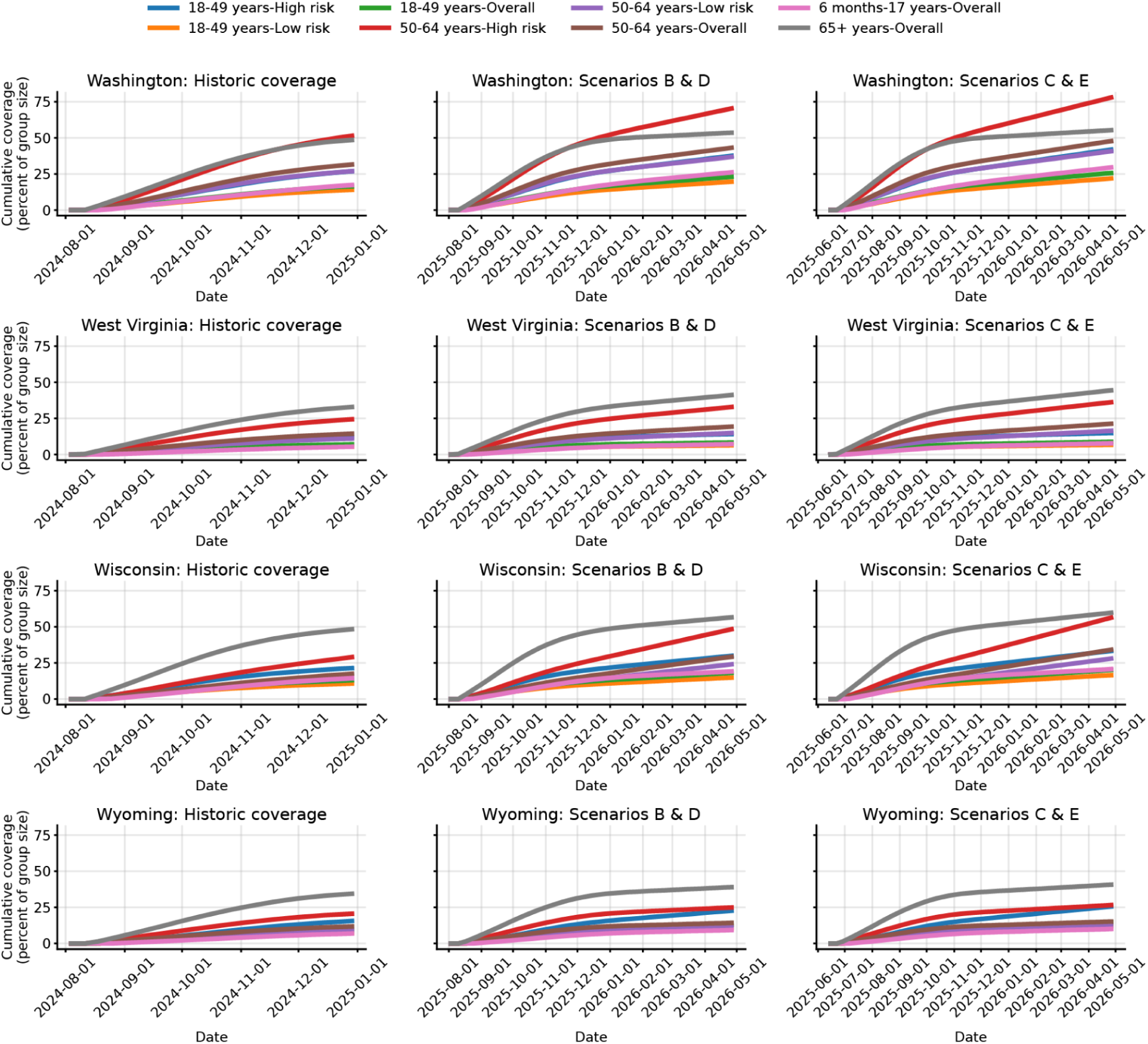
State and age specific vaccine coverage used for different vaccination scenarios from 1 Aug 2024 to 1 May 2026.

**Figure S2.**
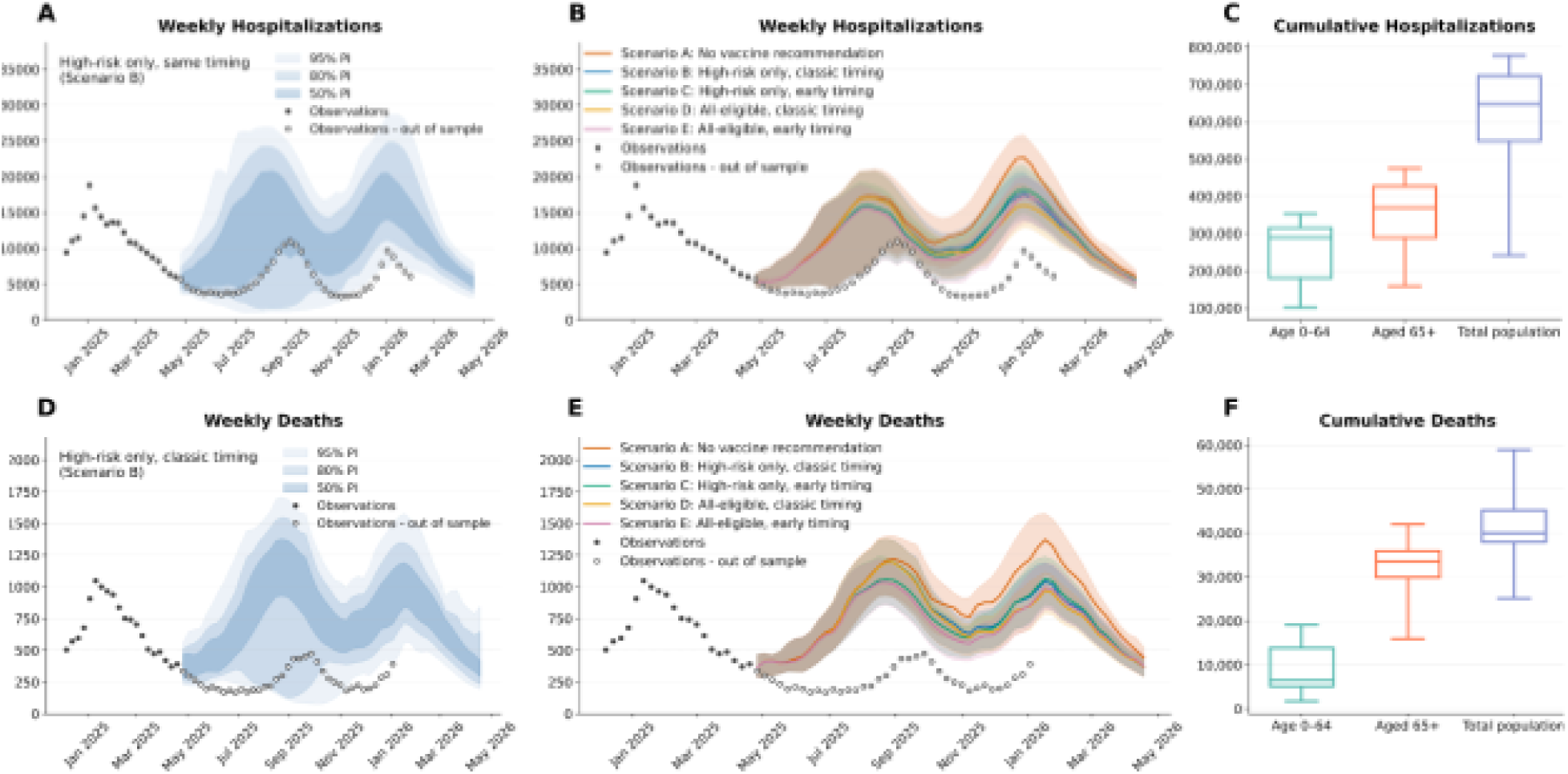
National-level ensemble predictions. A) Ensemble projections of weekly hospital admissions of Scenario B (High risk and classic timing). B) Combined hospital incidence projections across all scenarios. Shaded regions are the 50% projection interval and lines are median values. C) Cumulative hospitalizations across age groups, outer boxes represent the 50% projection interval and the whiskers represent the 95% PI. D) Ensemble projections of weekly deaths of Scenario B (High risk and classic timing). B) Combined weekly death projections across all scenarios. Shaded regions are the 50% projection interval and lines are median values. C) Cumulative deaths across age groups, outer boxes represent the 50% projection interval and the whiskers represent the 95% PI.

**Figure S3.**
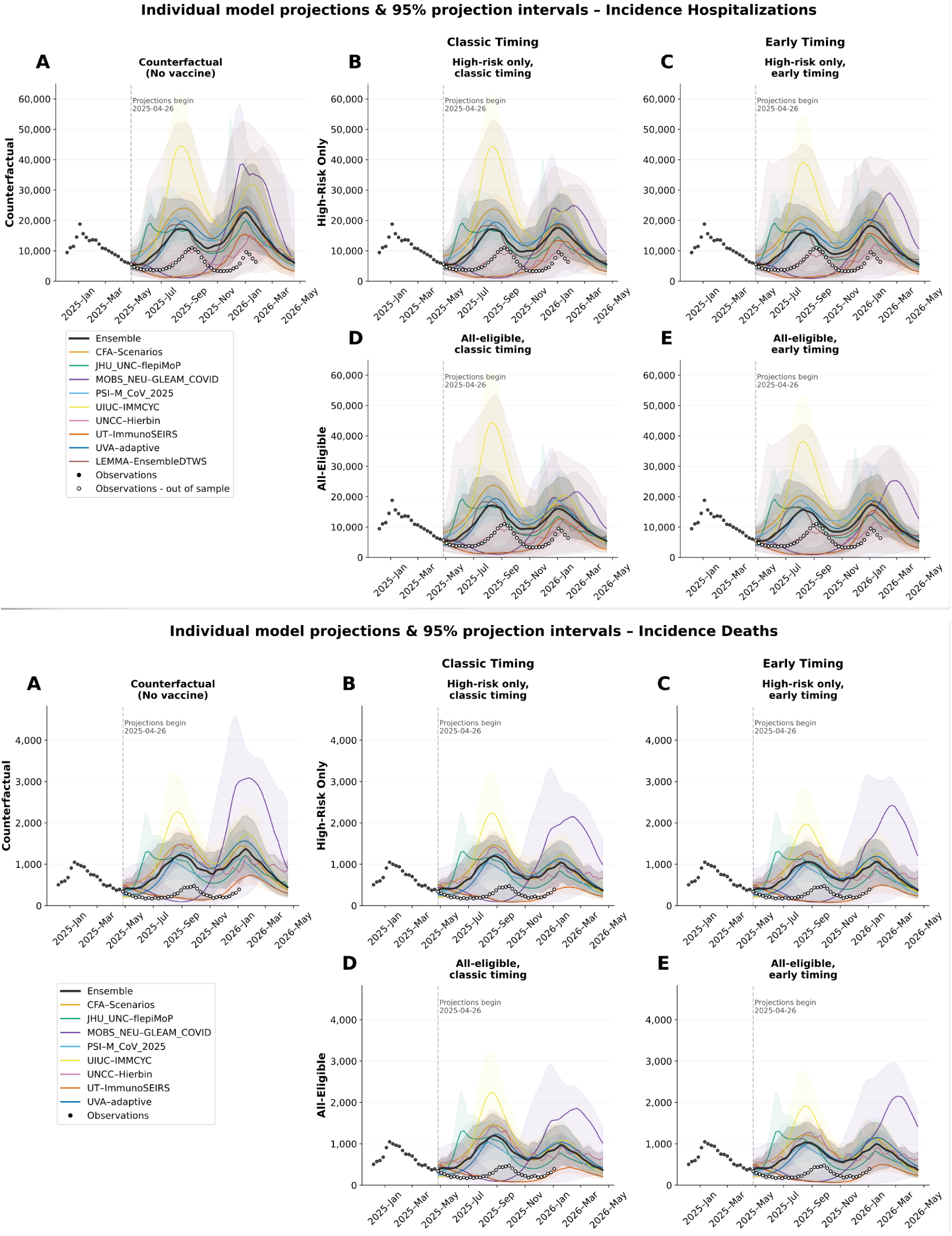
National-level individual-model projections of A) incident hospitalizations and B) incident deaths across scenarios.

**Figure S4.**
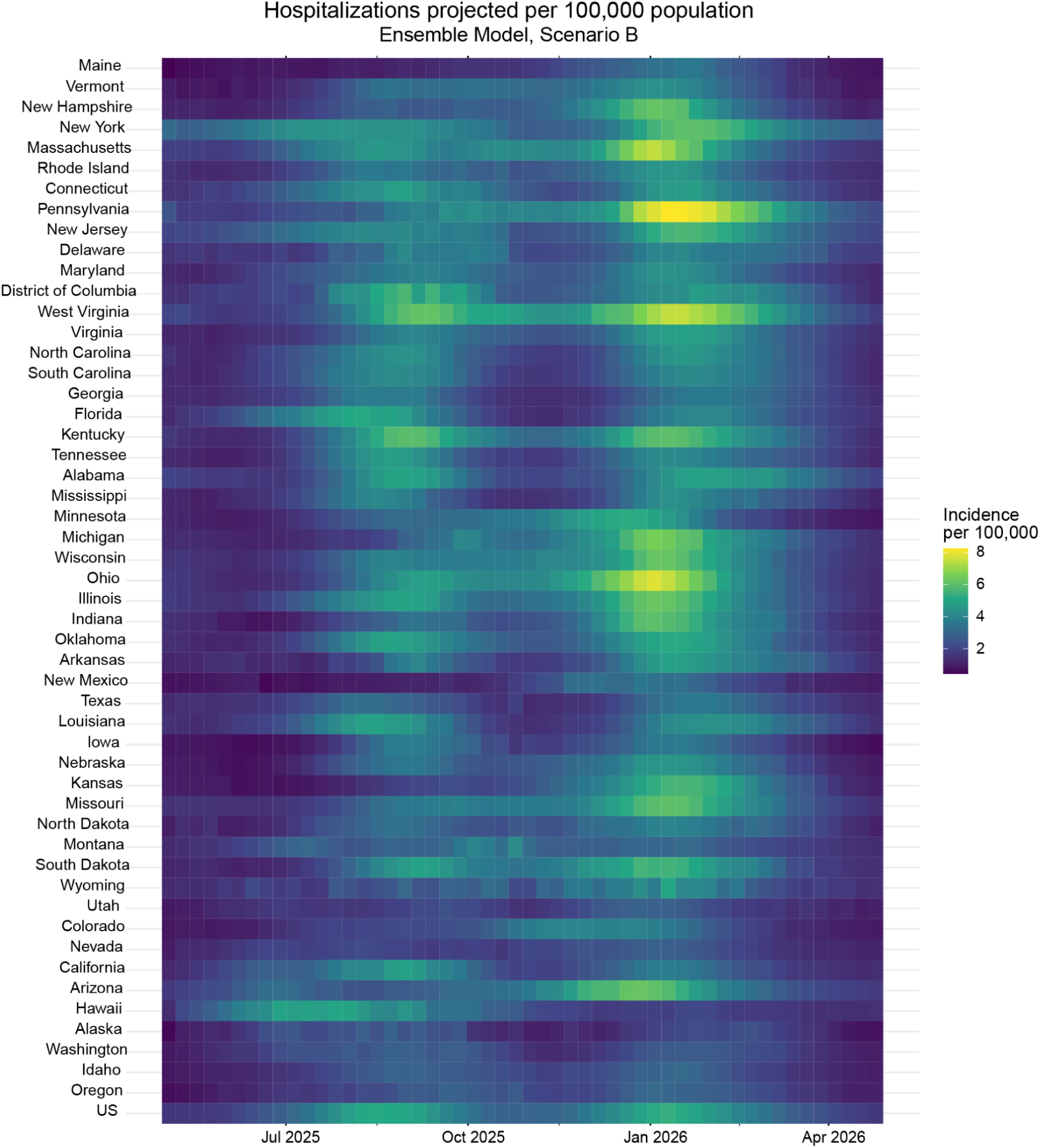
Median projected incident hospitalizations per 100,000 population for Scenario B, by US states in the ensemble model over the projection period 27 April 2025 to 25 April 2026.

**Figure S5.**
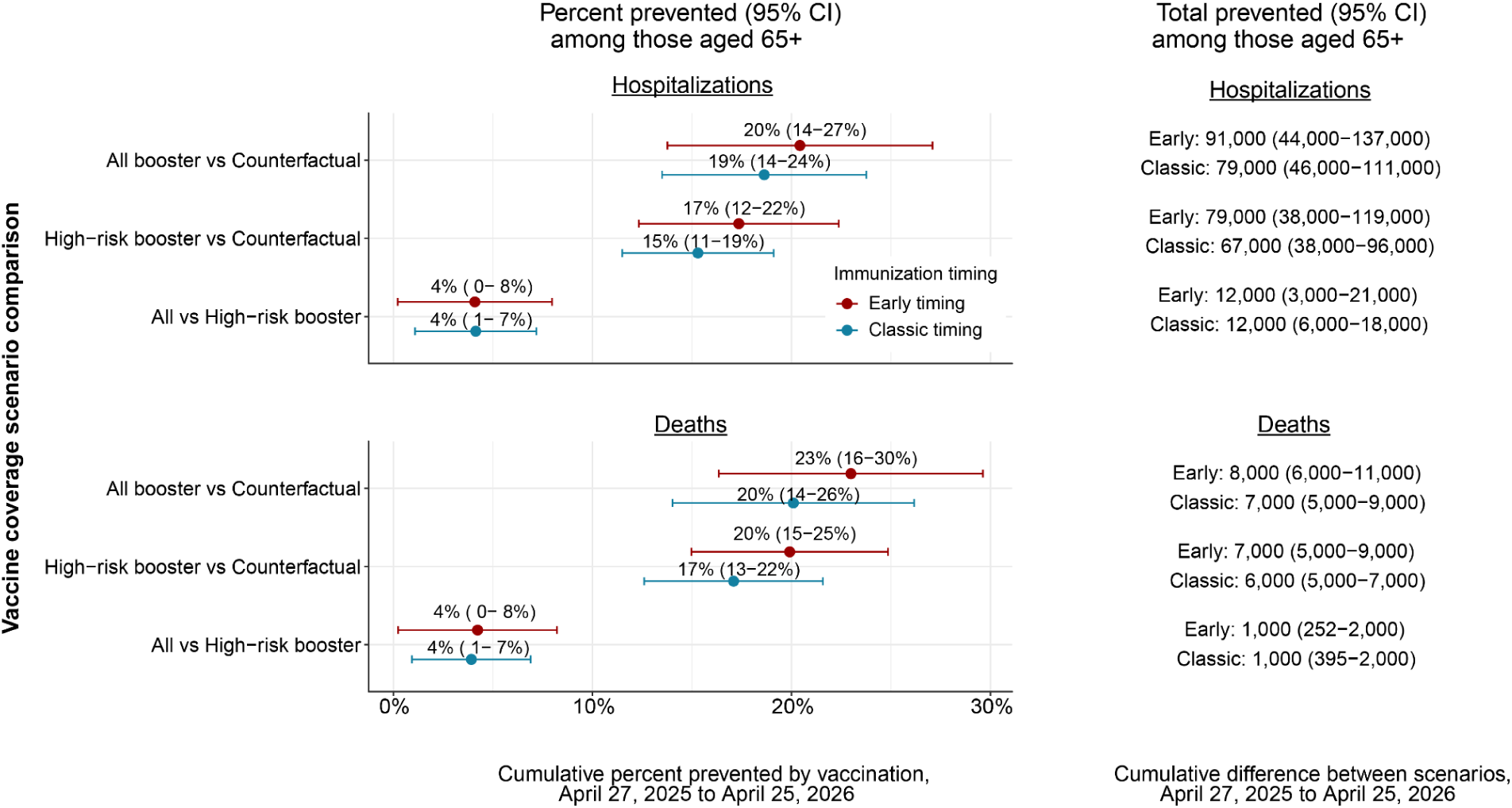
National-level impacts of vaccination coverage on hospitalizations and deaths across the projection period April 27, 2025 to April 25, 2026 for individuals aged 65 and over.

**Figure S6.**
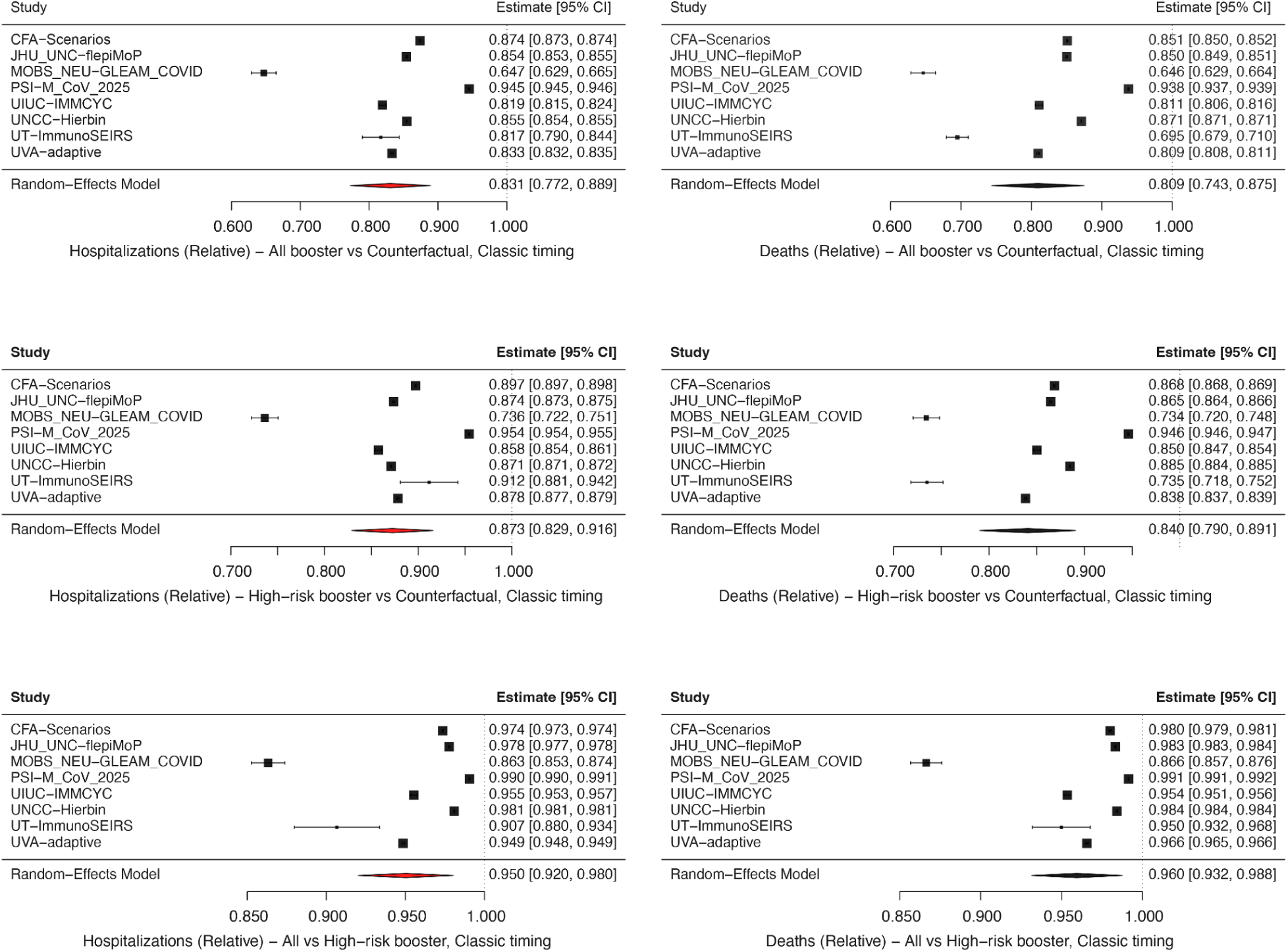
Relative differences between vaccination coverage scenarios by individual model, classic vaccination timing, for all age-groups, compared to the counterfactual (no vaccine recommendation) scenario across the projection period April 27, 2025 to April 25, 2026, United States.

**Figure S7.**
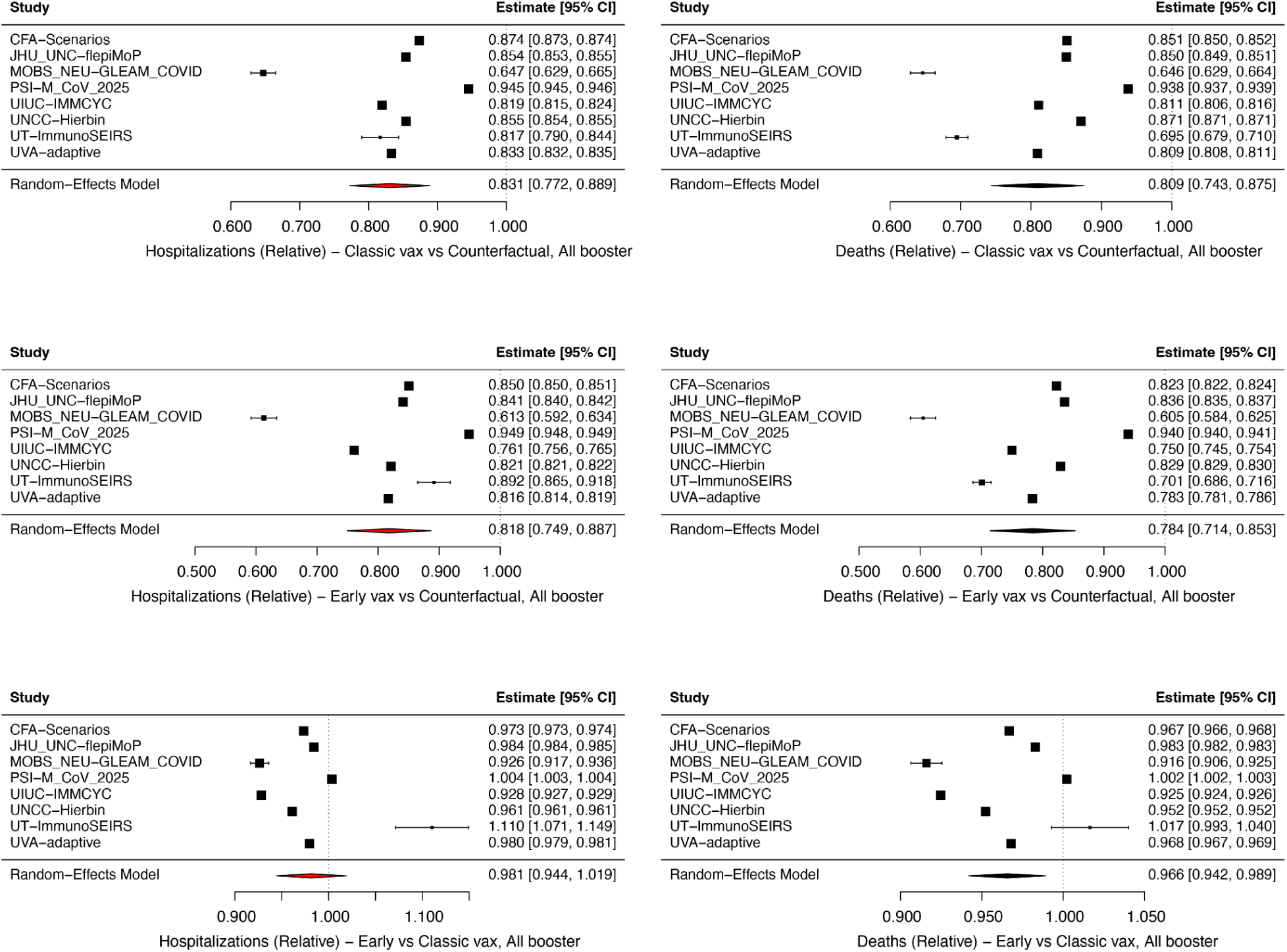
Relative differences between vaccination timing scenarios by individual model, all booster coverage, for all age-groups, compared to the counterfactual (no vaccine recommendation) scenario across the projection period April 27, 2025 to April 25, 2026, United States.

**Figure S8.**
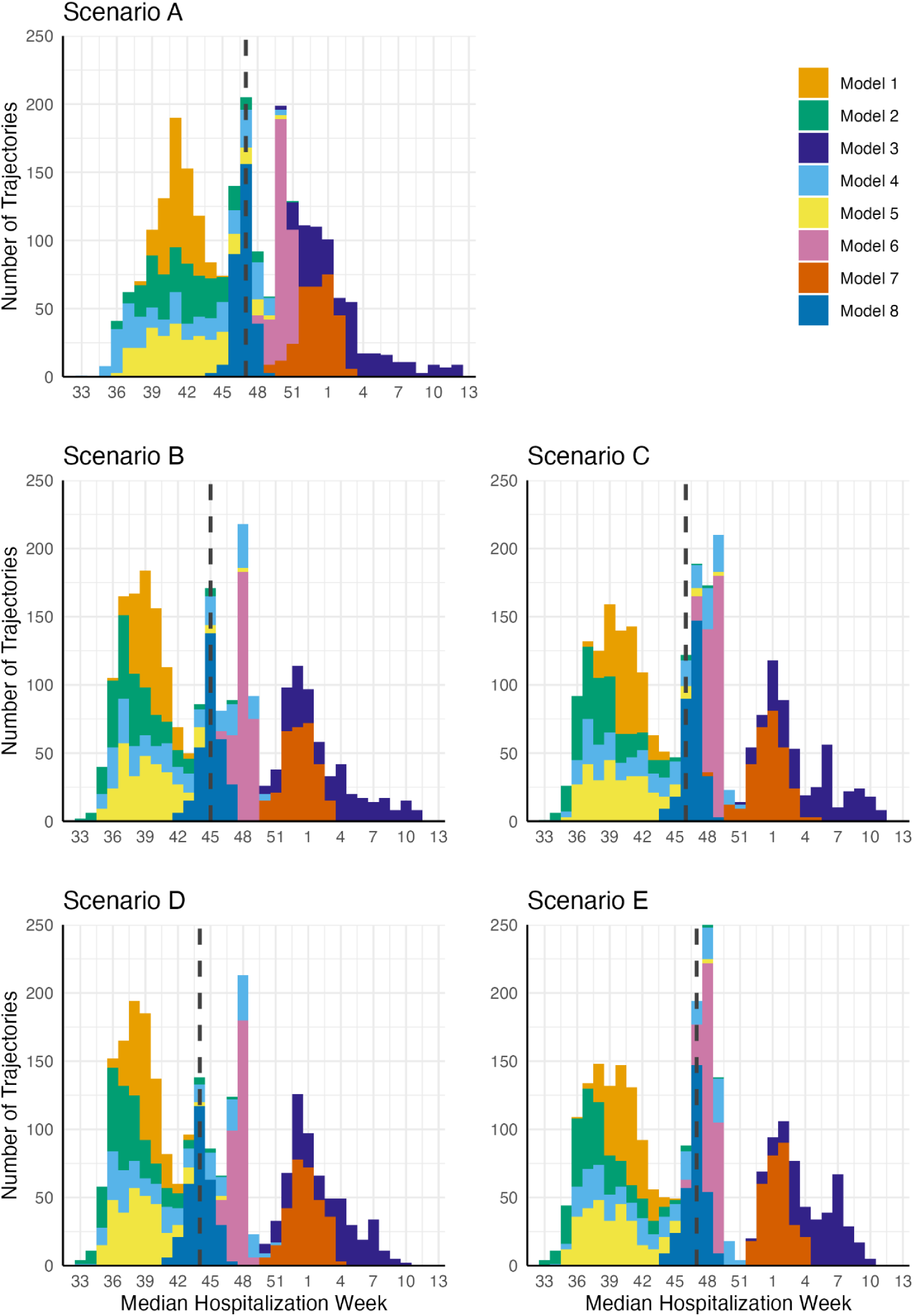
National trajectories distributed by median hospitalization week and colored by model. Vertical dashed lines indicate the median MHW across all models.

**Figure S9.**
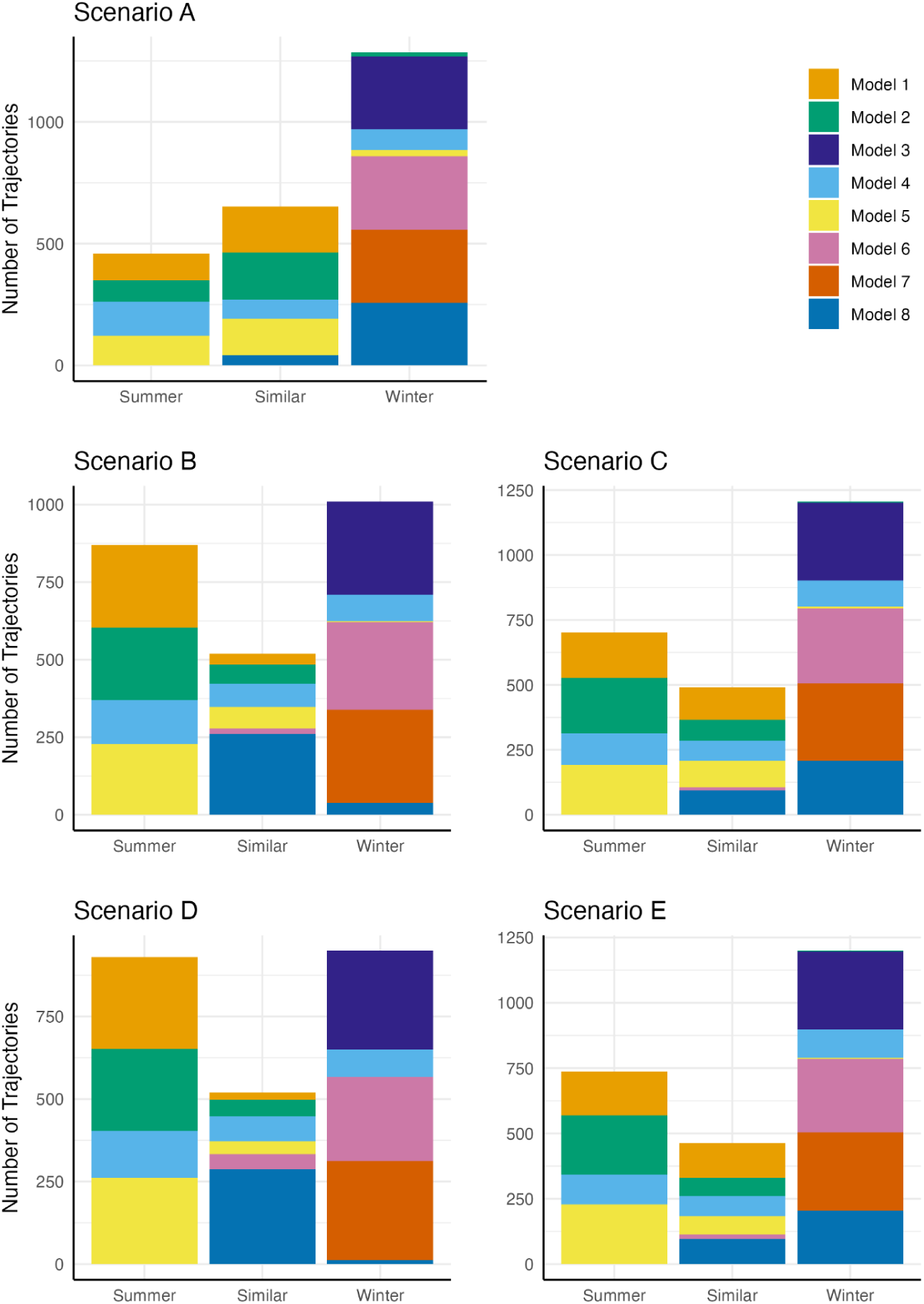
National trajectories characterized by larger summer/winter waves and colored by model. The categories are defined by trajectory cumulative proportion at Epidemic Week 43 (see methods for detail).

**Figure S10.**
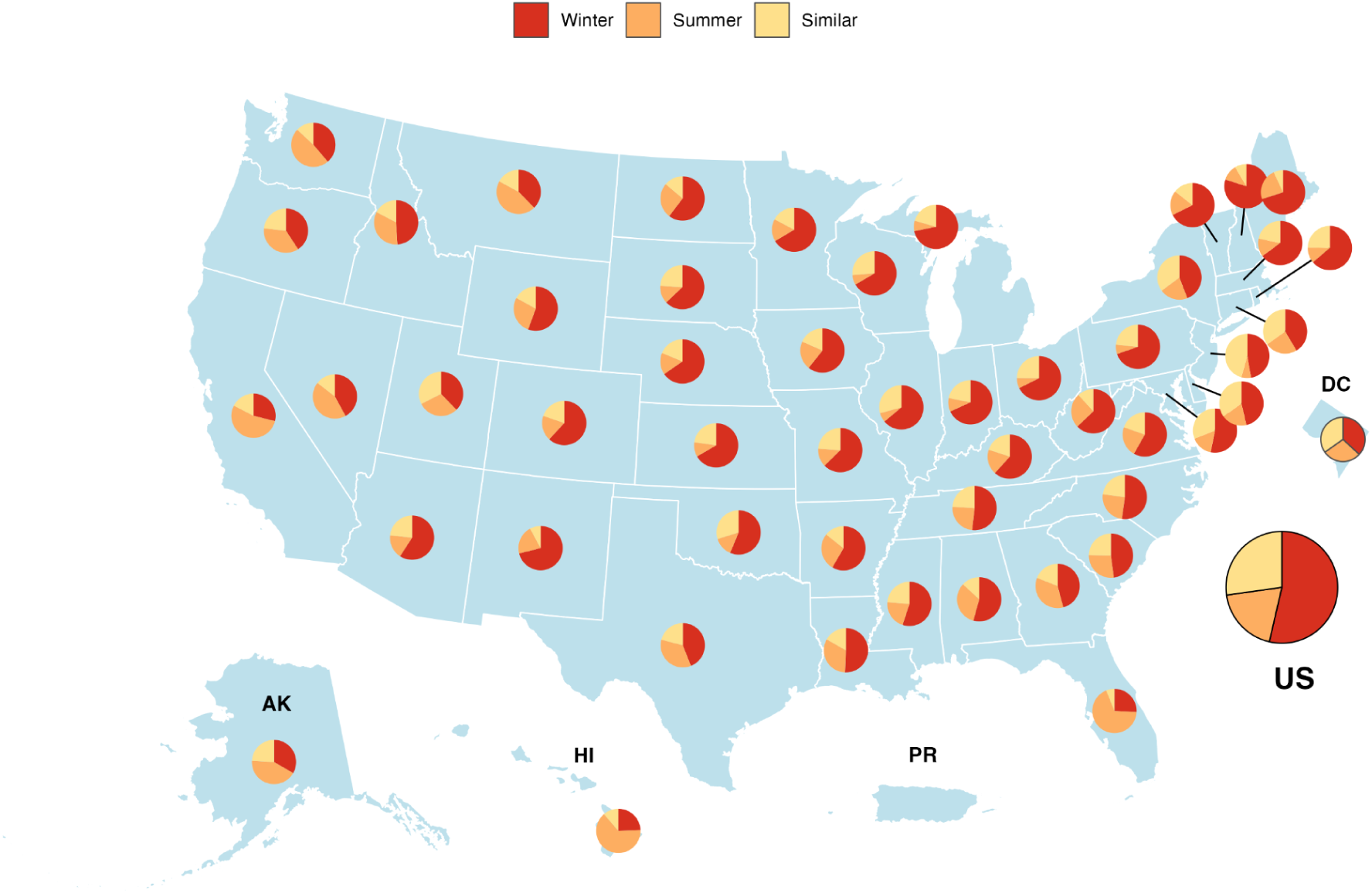
National and location-stratified trajectories for Scenario A, characterized by larger summer/winter waves. The categories are defined by trajectory cumulative proportion at Epidemic Week 43 (see methods for detail).

**Figure S11.**
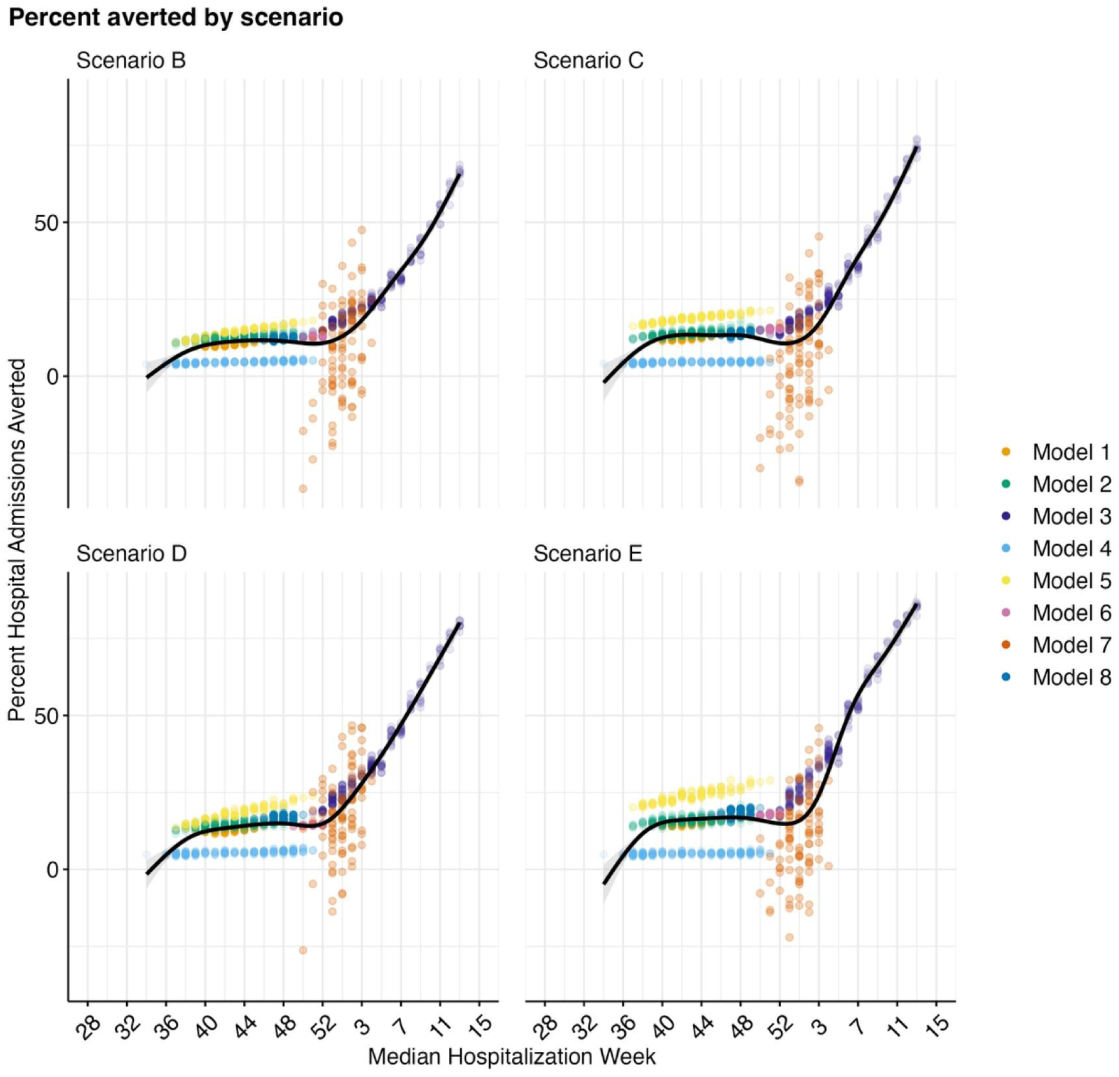
Hospital admissions averted by median hospitalization week at the national level. Points denote individual model trajectories; ensemble regressions (black lines) are generated with a Generalized Additive Model. Median hospitalization week is the epidemic week by which at least 50% of the total hospitalizations have been observed in the paired counterfactual Scenario A.

**Figure S12.**
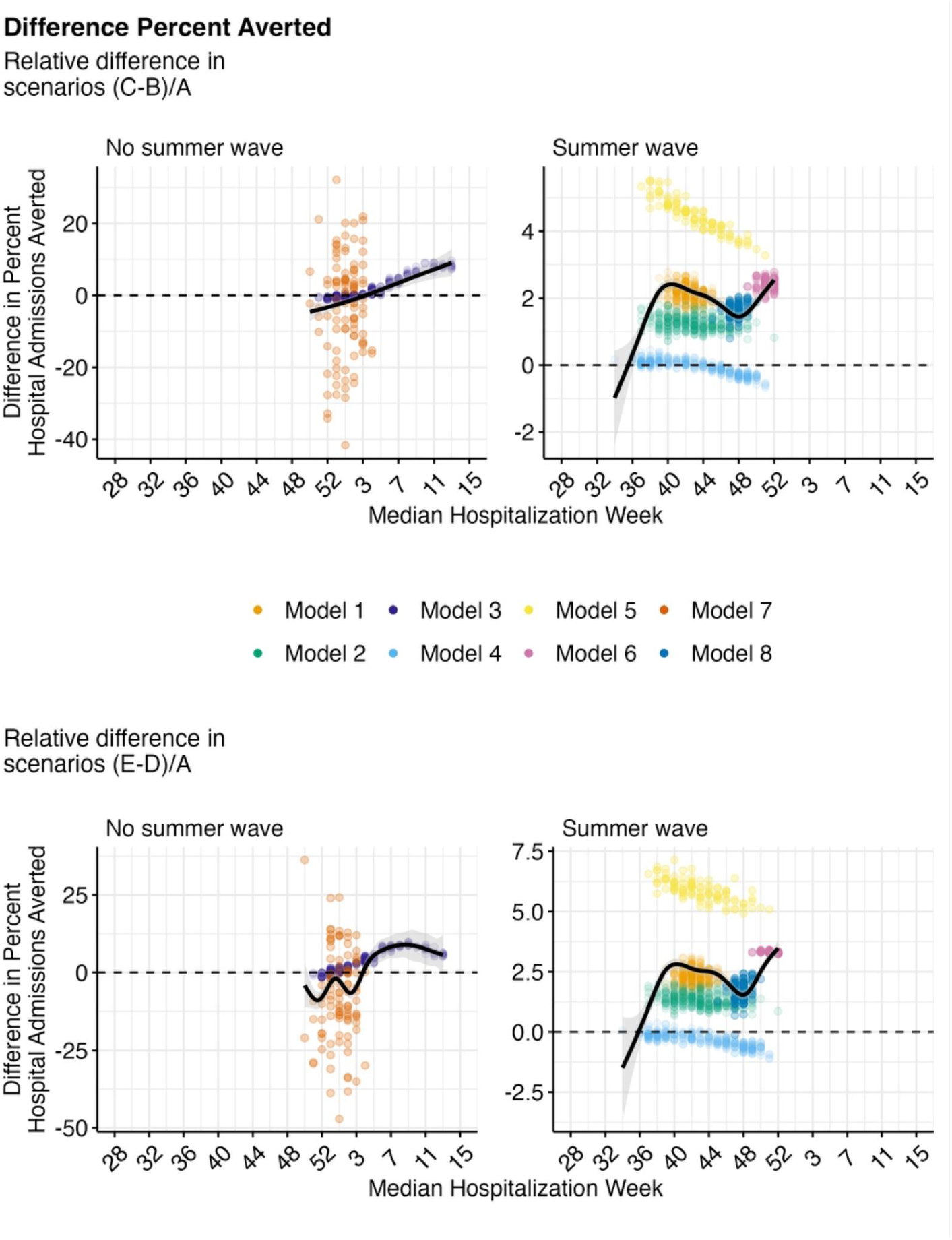
Difference in hospital admissions averted by median hospitalization week at the national level. Points denote individual model trajectories; ensemble regressions (black lines) are generated with a Generalized Additive Model. Dashed lines denote the threshold above which early timing is better and below which classic timing is better. Median hospitalization week is the epidemic week by which at least 50% of the total hospitalizations have been observed in the paired counterfactual Scenario A.

**Figure S13.**
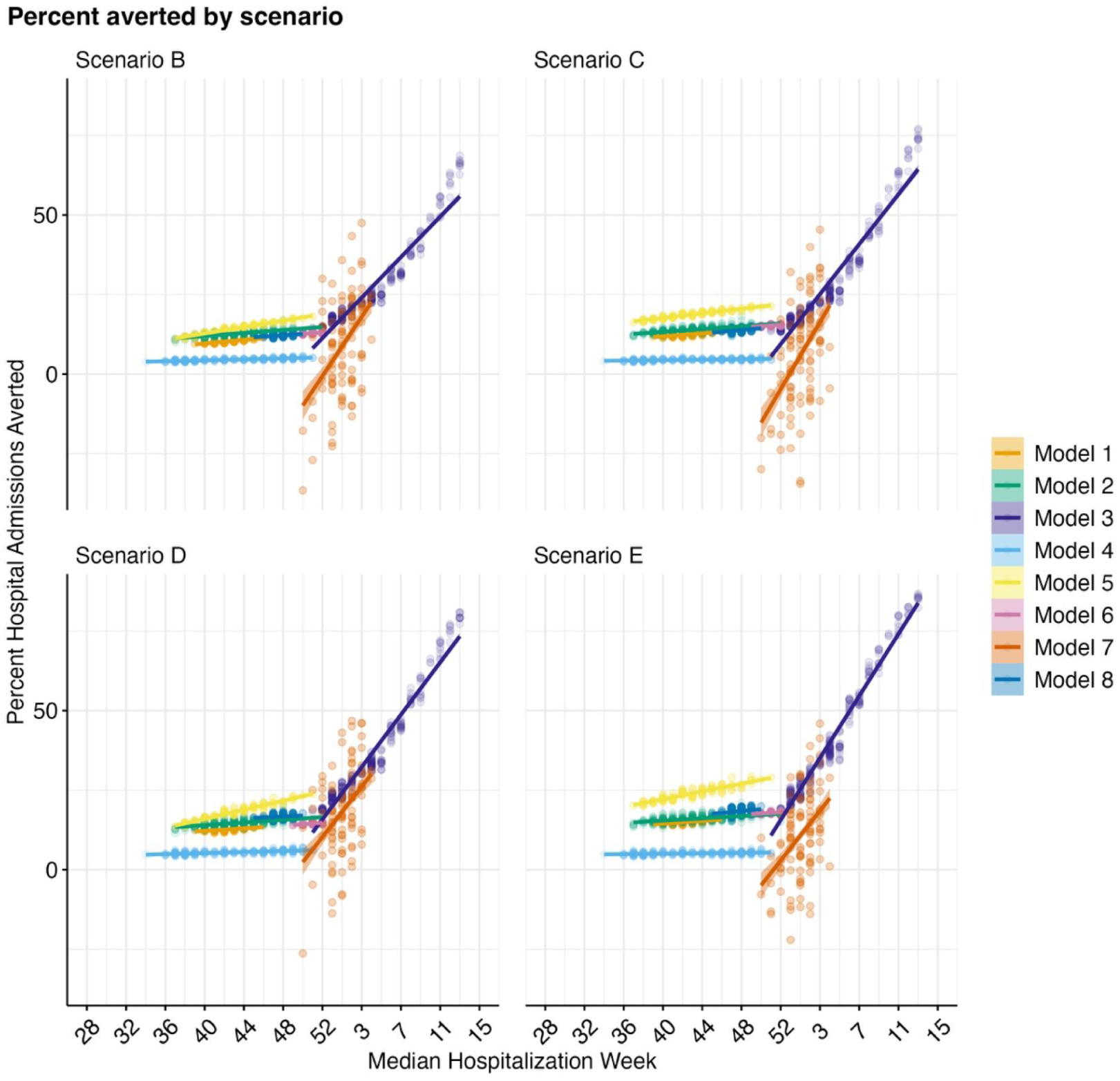
Hospital admissions averted by median hospitalization week at the national level, colored by model. Points denote individual model trajectories. Median hospitalization week is the epidemic week by which at least 50% of the total hospitalizations have been observed in the paired counterfactual Scenario A.

**Figure S14.**
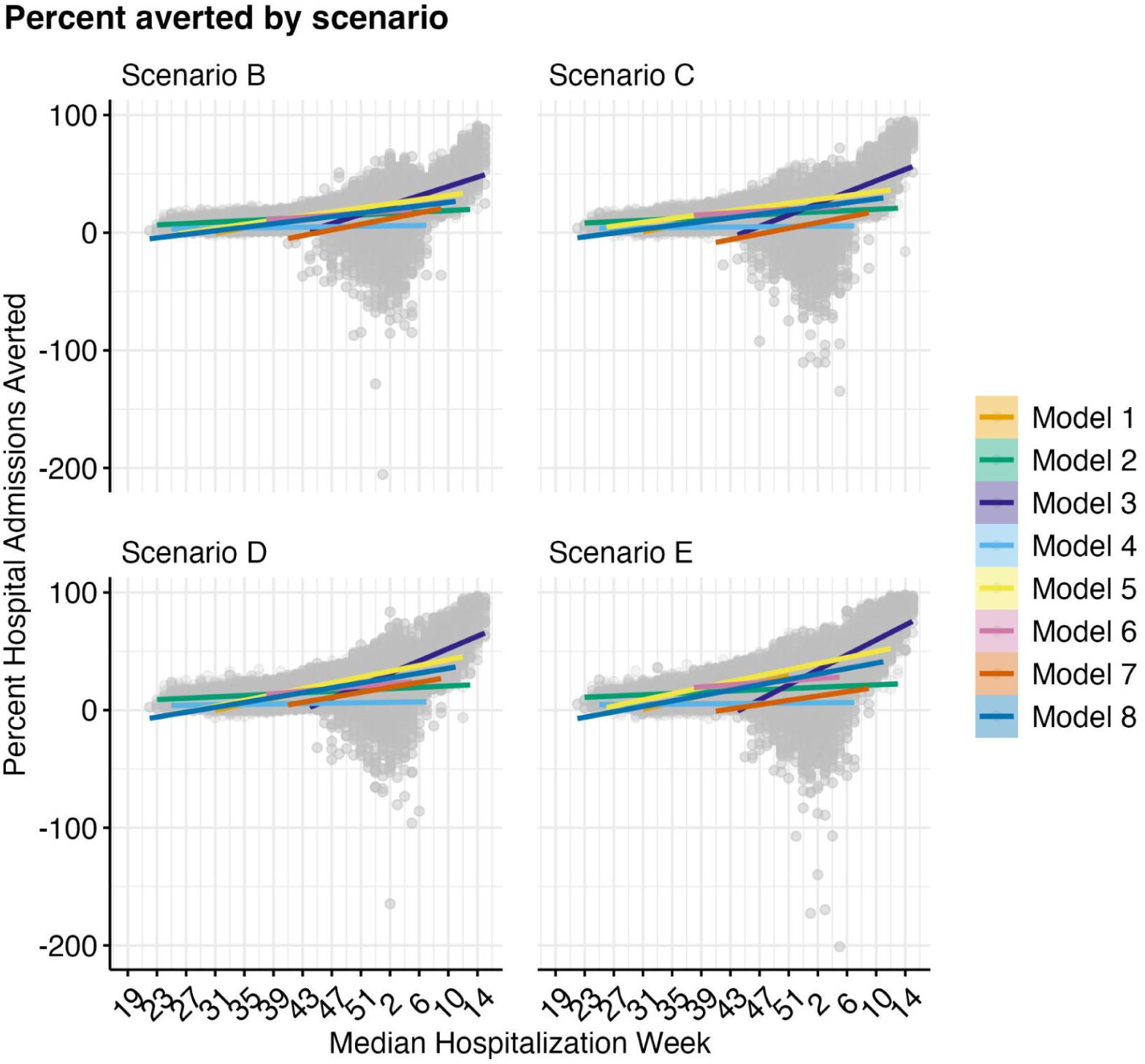
Hospital admissions averted by median hospitalization week across all subnational locations, colored by model. Points denote individual model trajectories. Median hospitalization week is the epidemic week by which at least 50% of the total hospitalizations have been observed in the paired counterfactual Scenario A.

**Figure S15.**
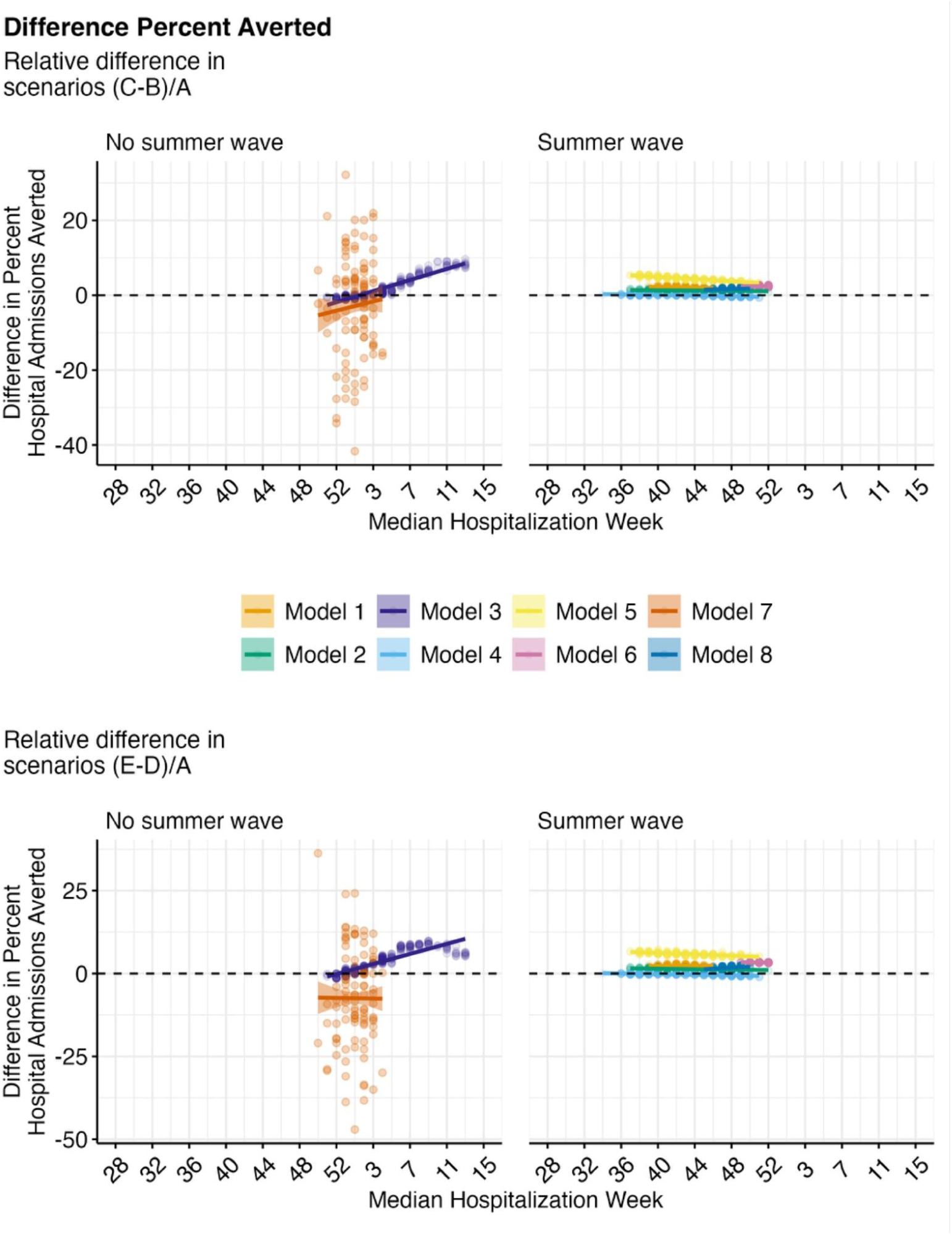
Hospital admissions averted by median hospitalization week, comparing scenarios C/B and E/D at the national level, colored by model. Points denote individual model trajectories. Dashed lines denote the threshold above which early timing is better and below which classic timing is better. Models are grouped by whether they project a summer wave. Median hospitalization week is the epidemic week by which at least 50% of the total hospitalizations have been observed in the paired counterfactual scenario (A).

**Figure S16.**
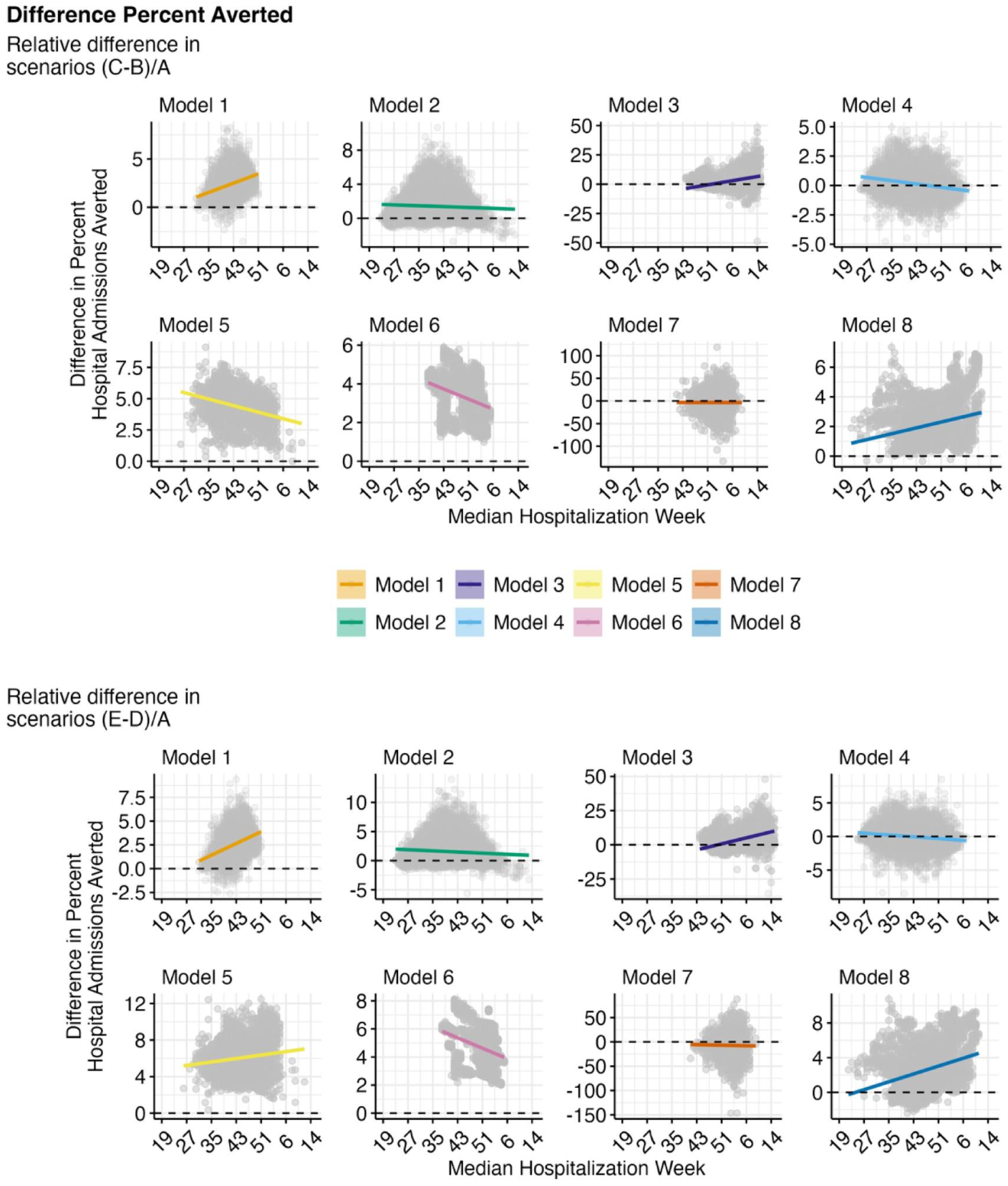
Hospital admissions averted by median hospitalization week across all states, as the difference between scenarios C/B and E/D. Points denote individual model trajectories. Dashed lines denote the threshold above which early timing is better and below which classic timing is better. Models are grouped by whether they project a summer wave. Median hospitalization week is the epidemic week by which at least 50% of the total hospitalizations have been observed in the paired counterfactual scenario (A).

**Figure S17.**
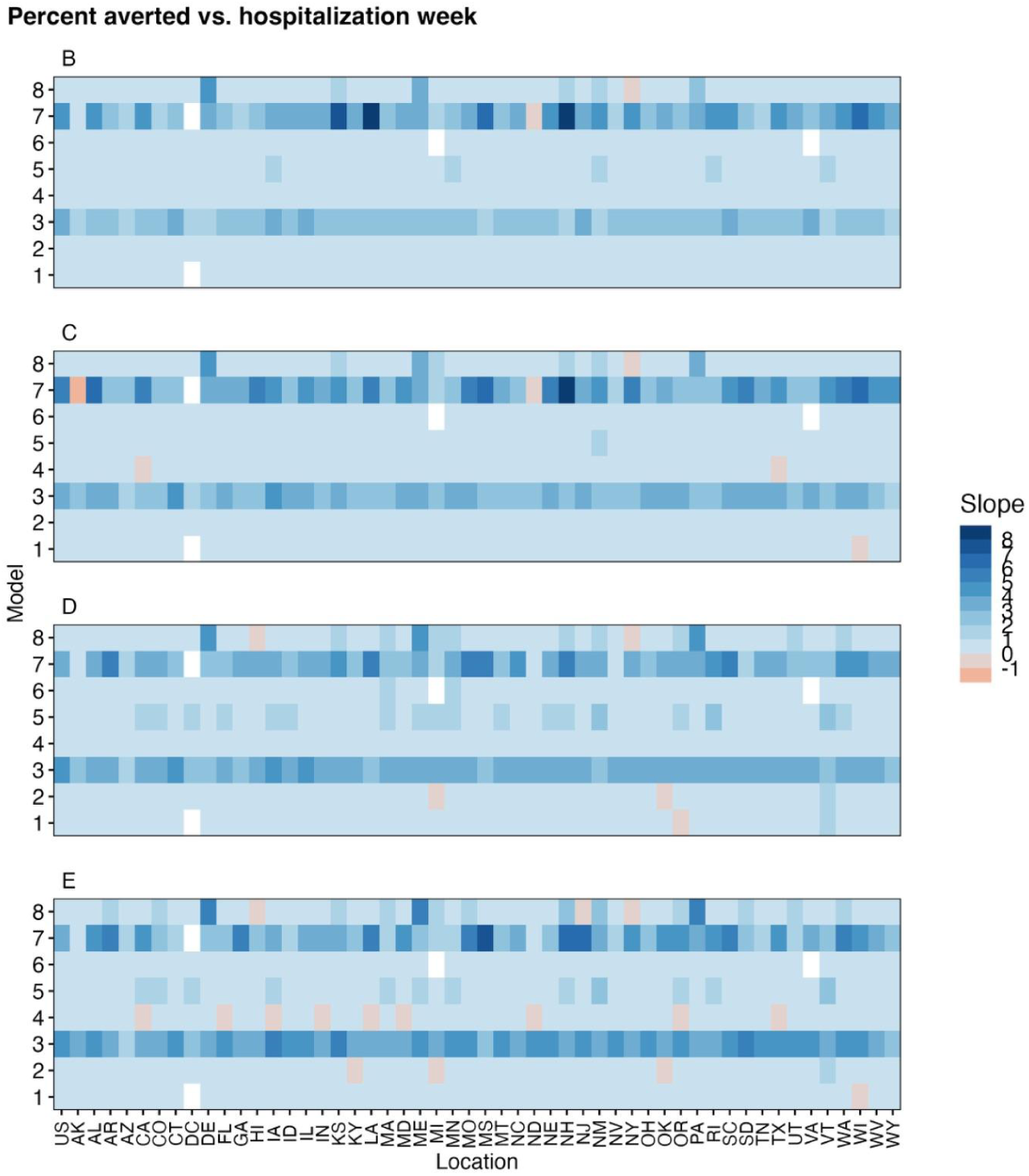
Slope of linear regression comparing percent averted to median hospitalization week, across models and locations. Median hospitalization week is the epidemic week by which at least 50% of the total hospitalizations have been observed in the paired counterfactual scenario (A).

**Figure S18.**
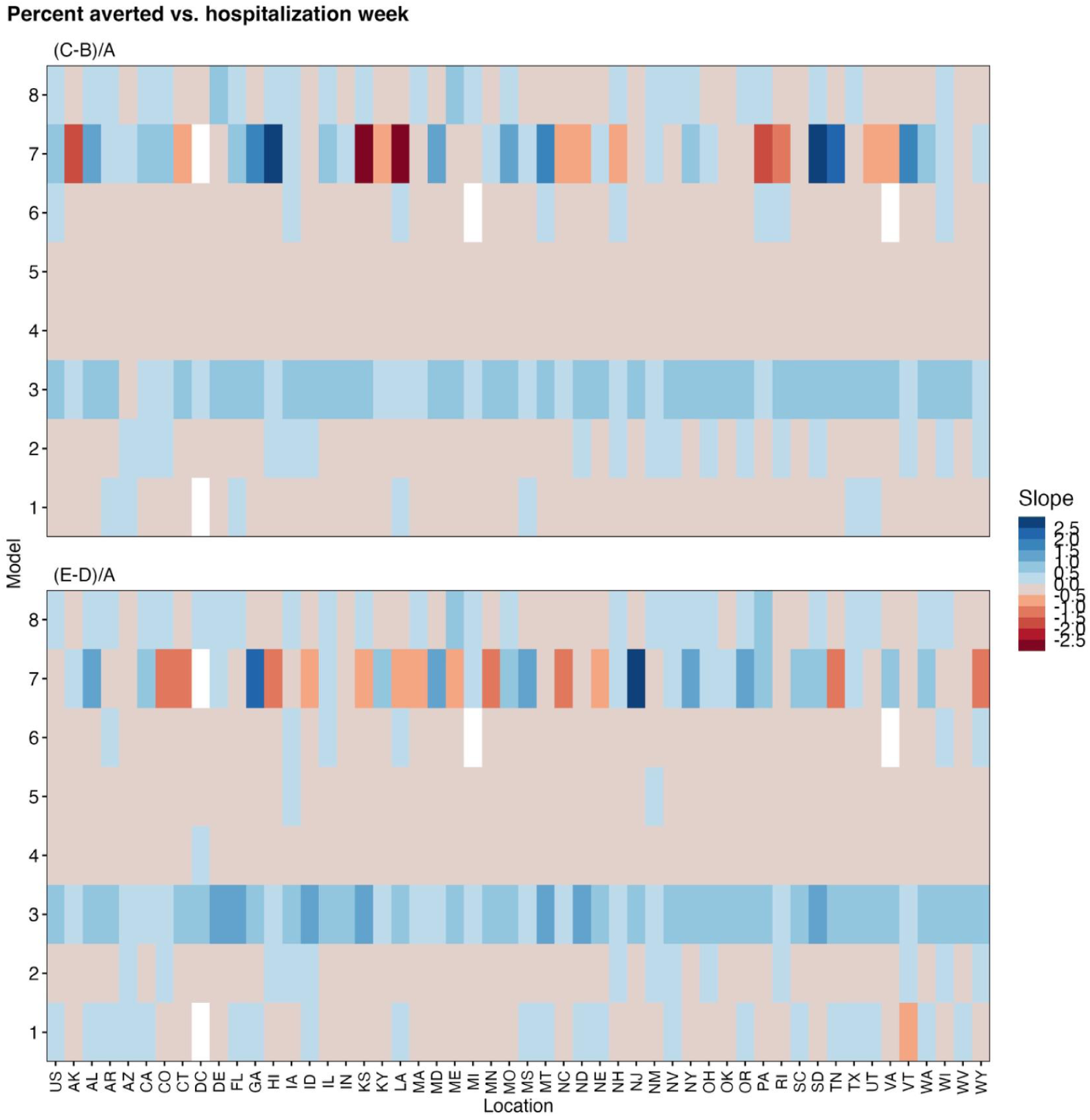
Slope of linear regression comparing difference in percent averted for scenarios C/B and D/E to median hospitalization week across models and locations. Median hospitalization week is the epidemic week by which at least 50% of the total hospitalizations have been observed in the paired counterfactual scenario (A).

**Figure S19.**
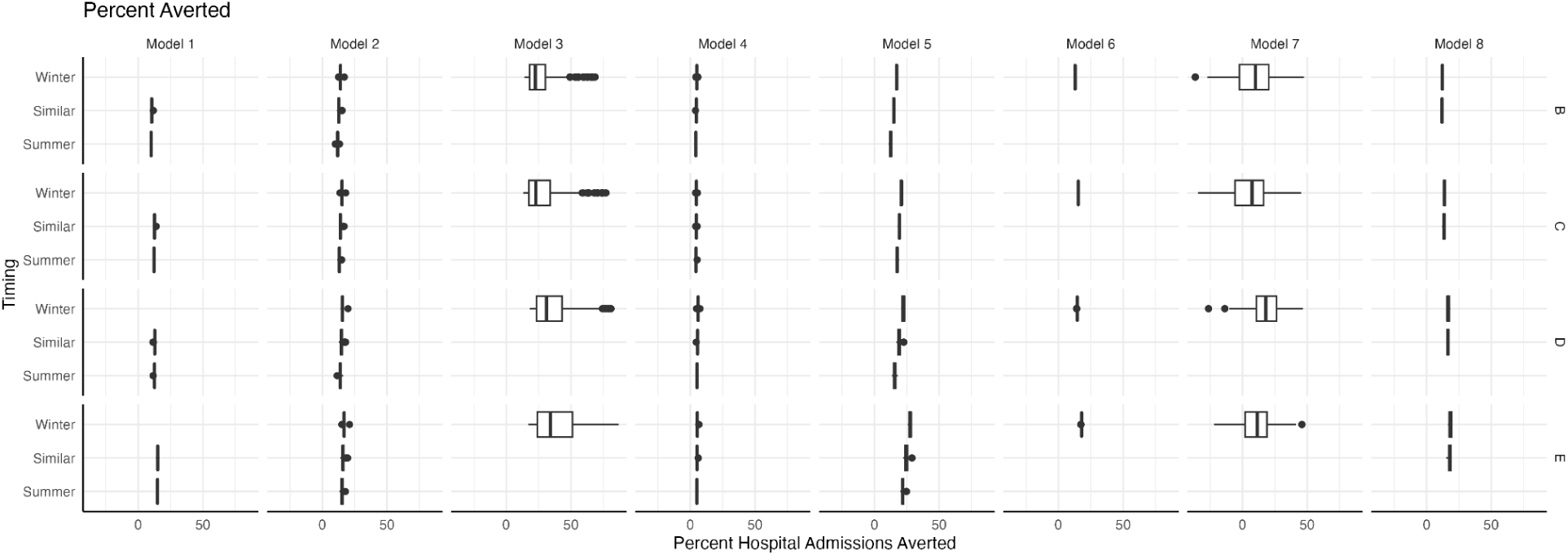
Percent averted by scenario and model, grouped by larger summer/winter wave. The categories are defined by trajectory cumulative proportion at Epidemic Week 43 (see methods for detail). Categories with fewer than 10 unique trajectories are removed.

**Figure S20.**
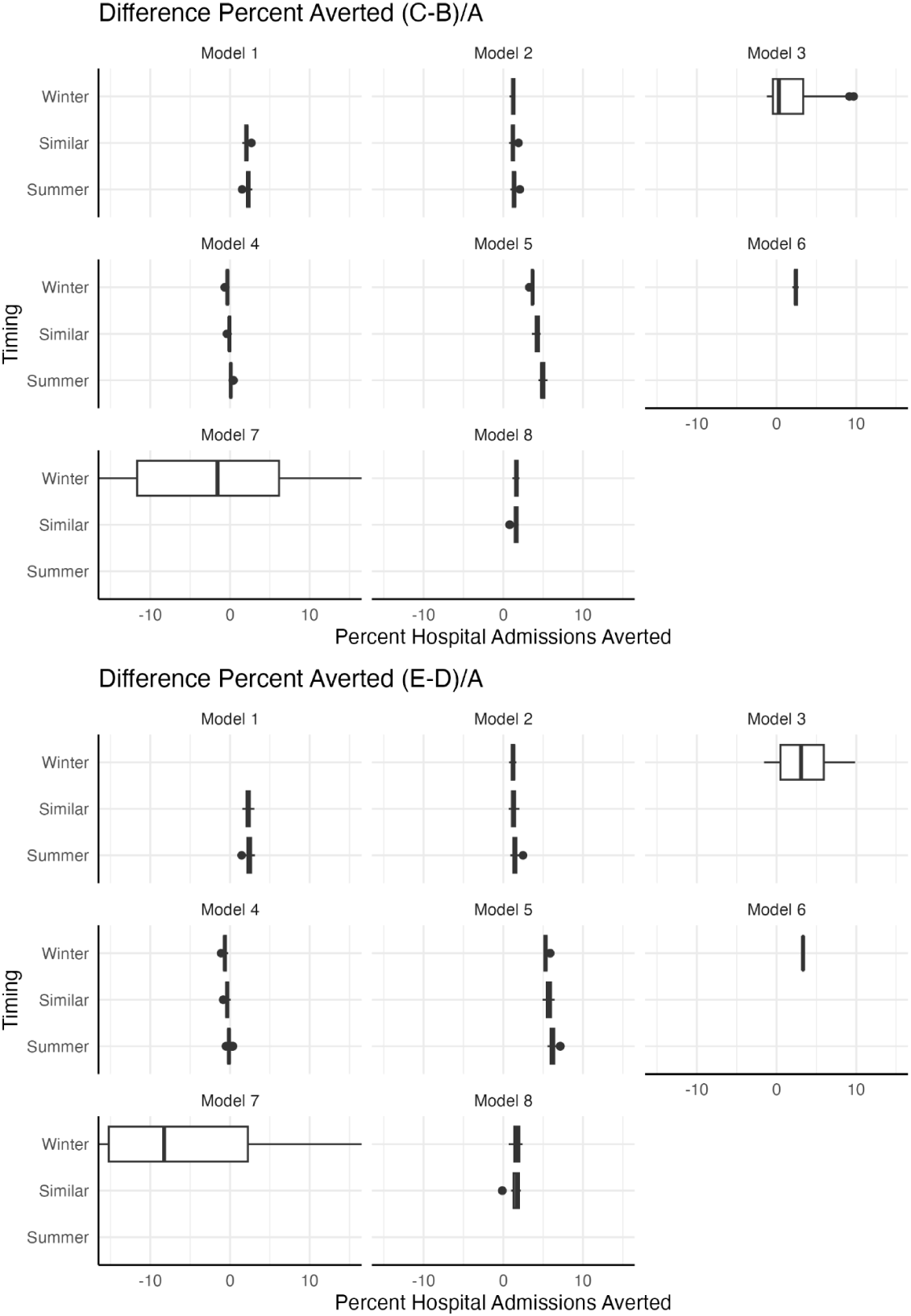
Percent averted comparing C/B and E/D by model, grouped by larger summer/winter wave. The categories are defined by trajectory cumulative proportion at Epidemic Week 43 (see methods for detail). Categories with fewer than 10 unique trajectories are removed.

**Figure S21.**
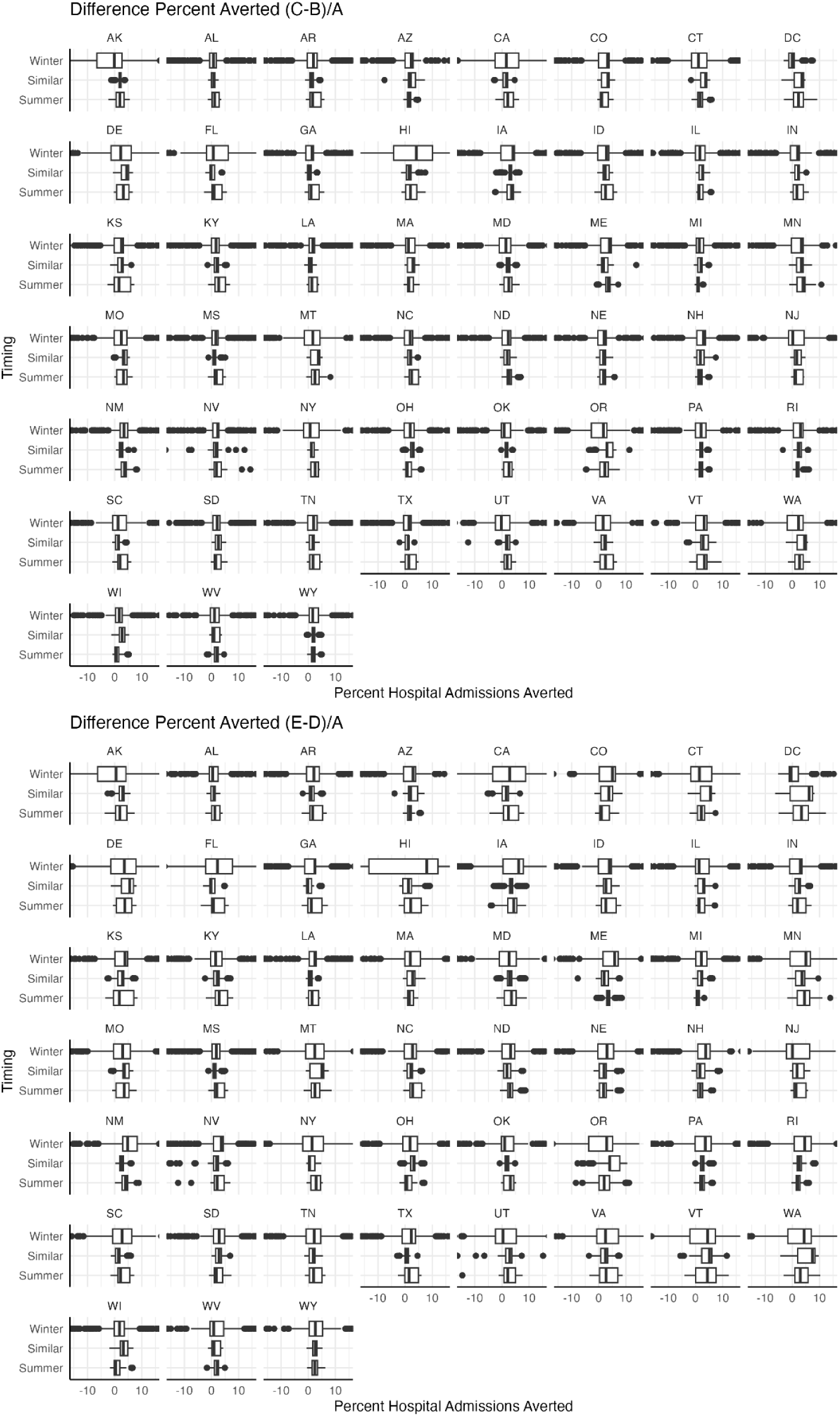
Percent averted comparing C/B and E/D by location, grouped by larger summer/winter wave. The categories are defined by trajectory cumulative proportion at Epidemic Week 43 (see methods for detail).

**Figure S22.**
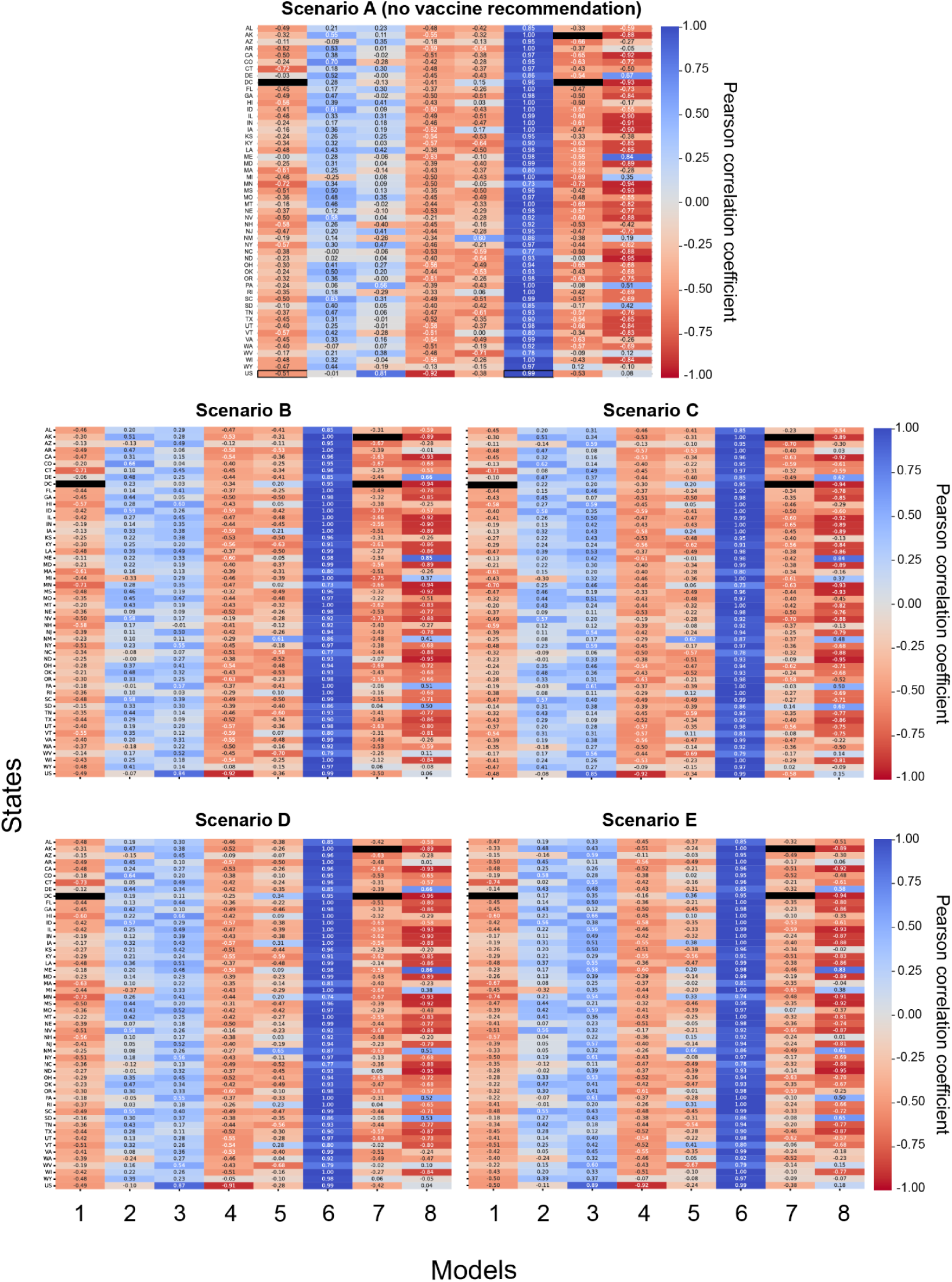
Pearson correlation coefficient *r* between summer and winter wave outbreak sizes for each team and location (state and national-level) for the no vaccine recommendation Scenario A and all vaccine recommendation scenarios B-E over the projection period April 27, 2025 to April 25, 2026. Here wave outbreak size corresponds to the number of hospitalizations per 100,000 individuals. Negative correlation (red) indicates a larger winter wave is associated with a smaller summer wave (example: Model 1), while positive correlation (blue) indicates a larger winter wave is associated with a larger summer wave (example: Model 6). Variation across models implies the relative sizes of the waves are influenced by different mechanisms within the models. Blacked out cells indicate insufficient data for this analysis.

**Figure S23.**
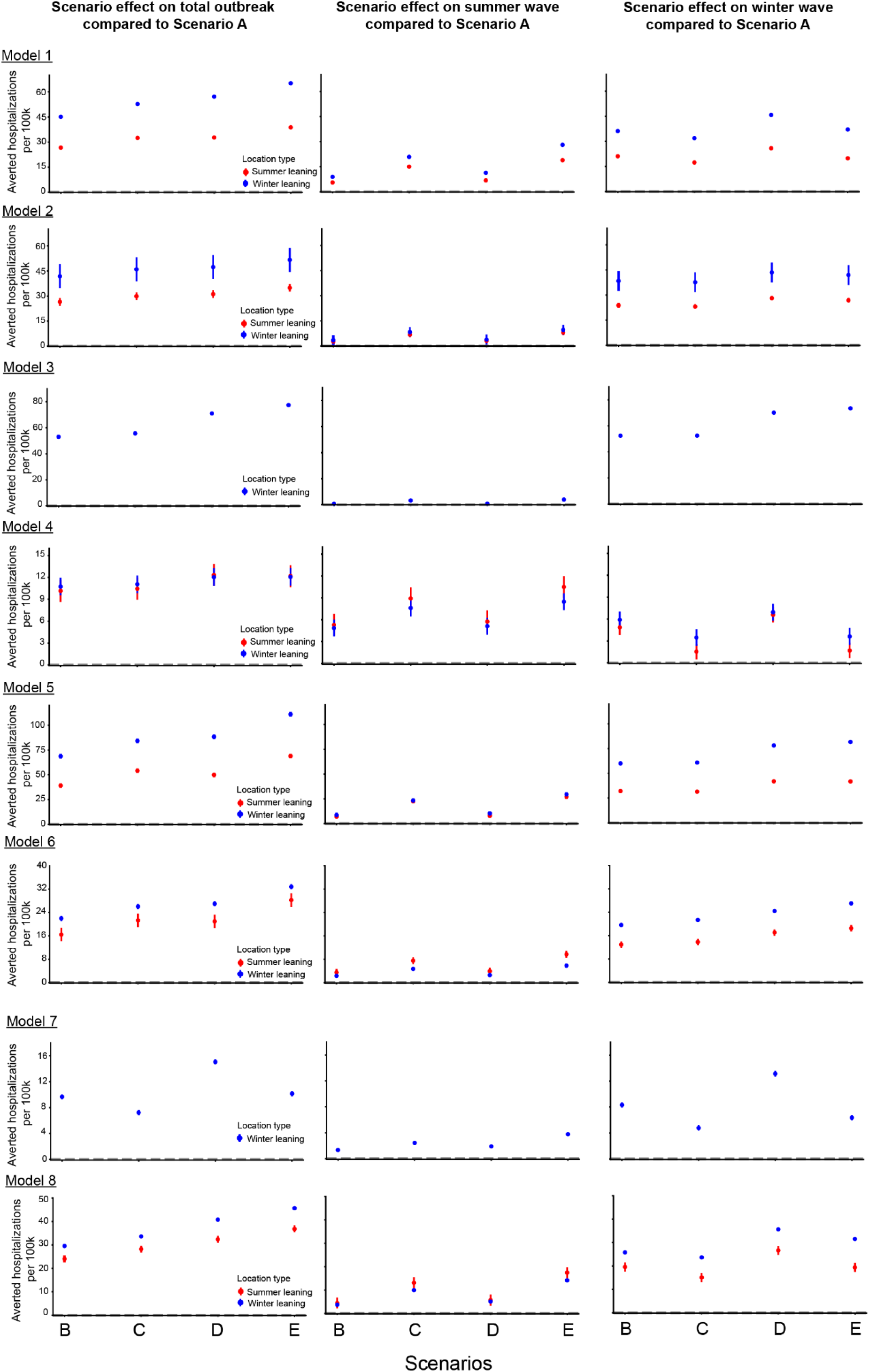
Vaccination scenario effects on the total outbreak, and summer and winter waves per team with scenarios as the fixed effect and location (states) as a random effect over the projection period April 27, 2025 to April 25, 2026. For each model, states are split into summer (red) or winter (blue) leaning based on which wave outbreak size was larger on average across Scenario A trajectories. Scenario effect is shown as the number of hospitalizations averted per 100,000. Error bars correspond to 95% confidence interval.

**Figure S24.**
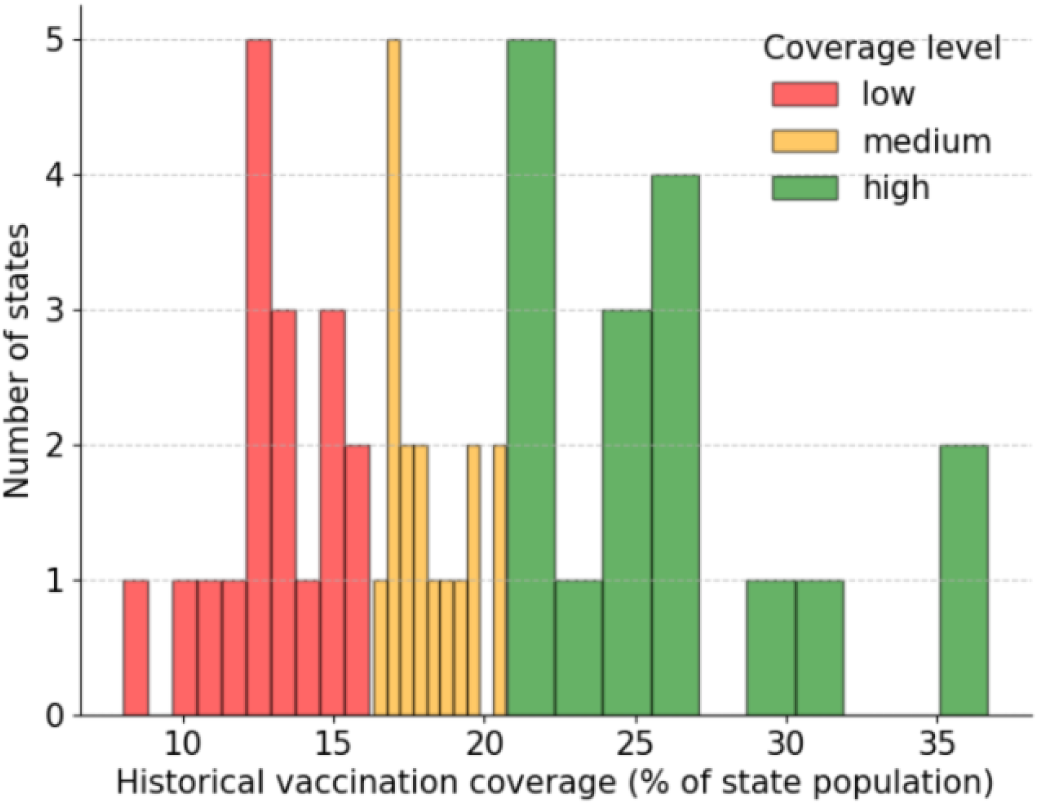
Distribution of states based on their vaccination coverage levels during the 2024-2025 campaign. States were divided into low, medium and high coverage-levels based on tertiles.

**Figure S25.**
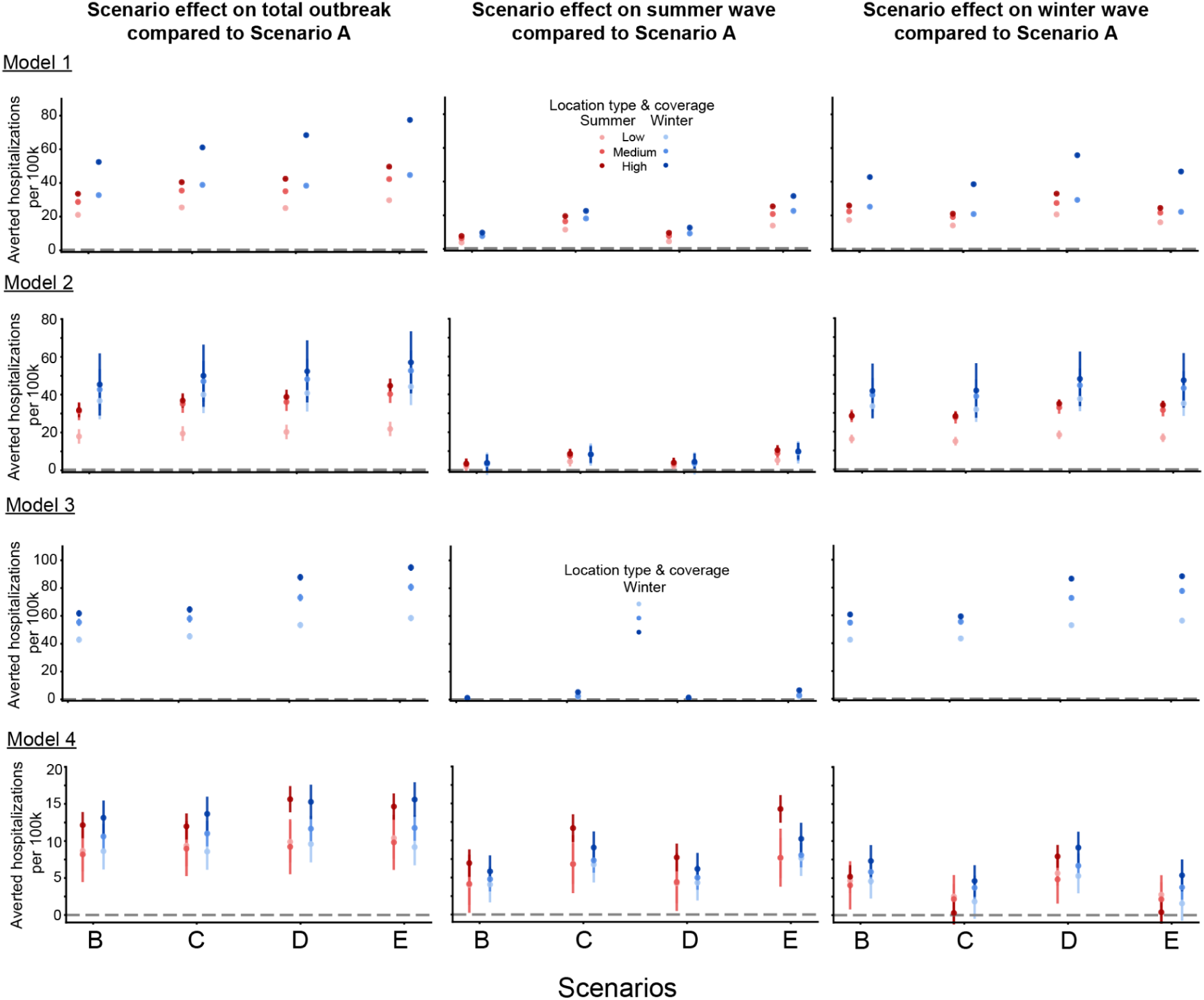
Vaccination scenario effects on the summer and winter wave outbreak sizes for Models 1-4 over the projection period April 27, 2025 to April 25, 2026. States are divided into low, medium and high vaccination coverage levels based on historical coverage levels. For each model, states are further split into summer (red) or winter (blue) leaning based on which wave outbreak size was larger on average across Scenario A trajectories. Scenario effect is expressed as the number of hospitalizations averted per 100,000. Error bars correspond to 95% CI.

**Figure S26.**
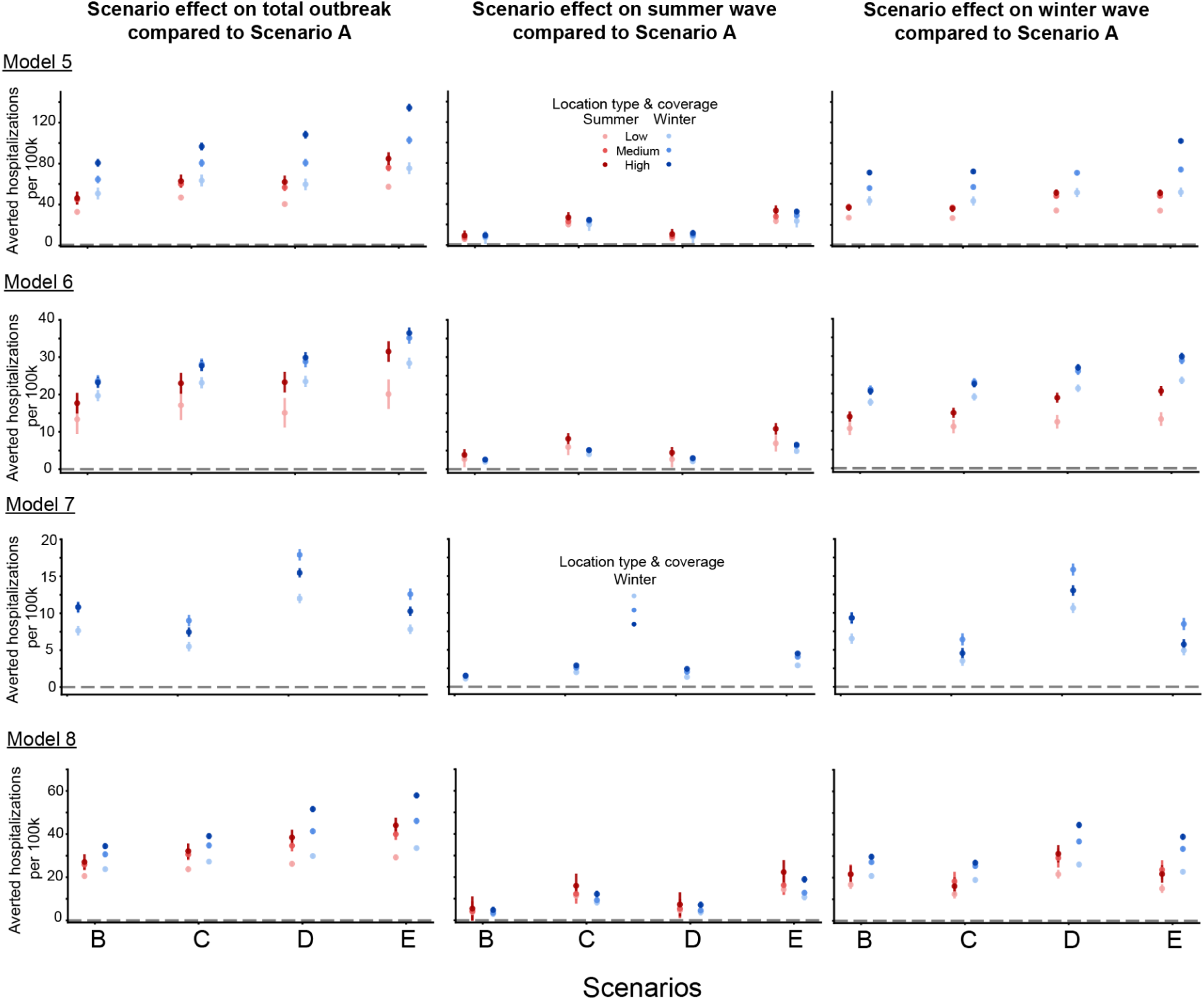
Vaccination scenario effects on the summer and winter wave outbreak sizes for Models 5-8 over the projection period April 27, 2025 to April 25, 2026. States are divided into low, medium and high vaccination coverage levels based on historical coverage levels. For each model, states are further split into summer (red) or winter (blue) leaning based on which wave outbreak size was larger on average across Scenario A trajectories. Scenario effect is expressed as the number of hospitalizations averted per 100,000. Error bars correspond to 95% CI.

### 4. Supplementary Tables

**Table S5.** Linear regression results for relative percent averted in Scenario B using the median horizon week of Scenario A as the independent variable.

| Model ID | Term | Estimate | Std. Error | t value | Pr(>\|t\|) | [0.025 | 0.975] |
| --- | --- | --- | --- | --- | --- | --- | --- |
| Model 1 | **Intercept** | 3.505 | 0.252 | 13.900 | 0.000 | 3.009 | 4.001 |
| Model 1 | **Slope** | 0.278 | 0.010 | 26.854 | 0.000 | 0.258 | 0.298 |
| Model 2 | **Intercept** | 6.789 | 0.283 | 23.981 | 0.000 | 6.232 | 7.346 |
| Model 2 | **Slope** | 0.236 | 0.011 | 20.778 | 0.000 | 0.213 | 0.258 |
| Model 3 | **Intercept** | -96.896 | 2.286 | -42.392 | 0.000 | -101.394 | -92.398 |
| Model 3 | **Slope** | 3.182 | 0.059 | 54.181 | 0.000 | 3.067 | 3.298 |
| Model 4 | **Intercept** | 2.695 | 0.089 | 30.380 | 0.000 | 2.520 | 2.869 |
| Model 4 | **Slope** | 0.075 | 0.003 | 21.394 | 0.000 | 0.068 | 0.082 |
| Model 5 | **Intercept** | 1.651 | 0.156 | 10.552 | 0.000 | 1.343 | 1.959 |
| Model 5 | **Slope** | 0.505 | 0.006 | 81.095 | 0.000 | 0.493 | 0.518 |
| Model 6 | **Intercept** | 6.520 | 0.485 | 13.440 | 0.000 | 5.565 | 7.475 |
| Model 6 | **Slope** | 0.192 | 0.015 | 13.111 | 0.000 | 0.163 | 0.221 |
| Model 7 | **Intercept** | -158.483 | 19.378 | -8.178 | 0.000 | -196.619 | -120.348 |
| Model 7 | **Slope** | 4.644 | 0.537 | 8.641 | 0.000 | 3.586 | 5.701 |
| Model 8 | **Intercept** | 6.358 | 1.032 | 6.161 | 0.000 | 4.327 | 8.389 |
| Model 8 | **Slope** | 0.195 | 0.035 | 5.621 | 0.000 | 0.127 | 0.263 |

**Table S6.** Linear regression results for relative percent averted in Scenario C using the median horizon week of Scenario A as the independent variable.

| Model ID | Term | Estimate | Std. Error | t value | Pr(>\|t\|) | [0.025 | 0.975] |
| --- | --- | --- | --- | --- | --- | --- | --- |
| Model 1 | **Intercept** | 6.632 | 0.399 | 16.628 | 0.000 | 5.847 | 7.417 |
| Model 1 | **Slope** | 0.237 | 0.016 | 14.495 | 0.000 | 0.205 | 0.270 |
| Model 2 | **Intercept** | 8.468 | 0.314 | 26.991 | 0.000 | 7.851 | 9.086 |
| Model 2 | **Slope** | 0.219 | 0.013 | 17.400 | 0.000 | 0.194 | 0.243 |
| Model 3 | **Intercept** | -123.950 | 2.617 | -47.358 | 0.000 | -129.101 | -118.799 |
| Model 3 | **Slope** | 3.923 | 0.067 | 58.336 | 0.000 | 3.791 | 4.056 |
| Model 4 | **Intercept** | 3.638 | 0.090 | 40.479 | 0.000 | 3.461 | 3.815 |
| Model 4 | **Slope** | 0.035 | 0.004 | 9.792 | 0.000 | 0.028 | 0.042 |
| Model 5 | **Intercept** | 9.785 | 0.184 | 53.172 | 0.000 | 9.423 | 10.147 |
| Model 5 | **Slope** | 0.358 | 0.007 | 48.922 | 0.000 | 0.344 | 0.373 |
| Model 6 | **Intercept** | 8.853 | 0.528 | 16.762 | 0.000 | 7.814 | 9.893 |
| Model 6 | **Slope** | 0.195 | 0.016 | 12.205 | 0.000 | 0.163 | 0.226 |
| Model 7 | **Intercept** | -183.548 | 19.910 | -9.219 | 0.000 | -222.729 | -144.366 |
| Model 7 | **Slope** | 5.260 | 0.552 | 9.526 | 0.000 | 4.173 | 6.346 |
| Model 8 | **Intercept** | 6.007 | 1.377 | 4.361 | 0.000 | 3.296 | 8.717 |
| Model 8 | **Slope** | 0.262 | 0.046 | 5.657 | 0.000 | 0.171 | 0.353 |

**Table S7.** Linear regression results for relative percent averted in Scenario D using the median horizon week of Scenario A as the independent variable.

| **Model ID** | **Term** | **Estimate** | **Std. Error** | **t value** | **Pr(>\|t\|)** | **[0.025** | **0.975]** |
| --- | --- | --- | --- | --- | --- | --- | --- |
| Model 1 | **Intercept** | 7.774 | 0.389 | 19.989 | 0.000 | 7.008 | 8.539 |
| Model 1 | **Slope** | 0.200 | 0.016 | 12.522 | 0.000 | 0.169 | 0.231 |
| Model 2 | **Intercept** | 9.391 | 0.424 | 22.142 | 0.000 | 8.556 | 10.226 |
| Model 2 | **Slope** | 0.209 | 0.017 | 12.329 | 0.000 | 0.176 | 0.243 |
| Model 3 | **Intercept** | -123.961 | 2.082 | -59.543 | 0.000 | -128.058 | -119.864 |
| Model 3 | **Slope** | 4.111 | 0.053 | 76.853 | 0.000 | 4.006 | 4.216 |
| Model 4 | **Intercept** | 3.411 | 0.147 | 23.206 | 0.000 | 3.122 | 3.700 |
| Model 4 | **Slope** | 0.082 | 0.006 | 14.238 | 0.000 | 0.071 | 0.094 |
| Model 5 | **Intercept** | 0.343 | 0.311 | 1.105 | 0.270 | -0.268 | 0.955 |
| Model 5 | **Slope** | 0.711 | 0.012 | 57.479 | 0.000 | 0.687 | 0.735 |
| Model 6 | **Intercept** | 7.083 | 0.427 | 16.587 | 0.000 | 6.243 | 7.924 |
| Model 6 | **Slope** | 0.225 | 0.013 | 17.483 | 0.000 | 0.200 | 0.251 |
| Model 7 | **Intercept** | -124.03 | 17.054 | -7.273 | 0.000 | -157.592 | -90.469 |
| Model 7 | **Slope** | 3.951 | 0.473 | 8.354 | 0.000 | 3.020 | 4.881 |
| Model 8 | **Intercept** | 11.906 | 1.703 | 6.993 | 0.000 | 8.555 | 15.256 |
| Model 8 | **Slope** | 0.160 | 0.057 | 2.799 | 0.005 | 0.048 | 0.273 |

**Table S8.** Linear regression results for relative percent averted in Scenario E using the median horizon week of Scenario A as the independent variable.

| **Model ID** | **Term** | **Estimate** | **Std. Error** | **t value** | **Pr(>\|t\|)** | **[0.025** | **0.975** |
| --- | --- | --- | --- | --- | --- | --- | --- |
| Model 1 | **Intercept** | 9.927 | 0.451 | 22.025 | 0.000 | 9.040 | 10.814 |
| Model 1 | **Slope** | 0.207 | 0.019 | 11.193 | 0.000 | 0.171 | 0.244 |
| Model 2 | **Intercept** | 11.496 | 0.462 | 24.872 | 0.000 | 10.587 | 12.406 |
| Model 2 | **Slope** | 0.178 | 0.019 | 9.642 | 0.000 | 0.142 | 0.215 |
| Model 3 | **Intercept** | -149.950 | 1.932 | -77.633 | 0.000 | -153.751 | -146.149 |
| Model 3 | **Slope** | 4.870 | 0.050 | 98.134 | 0.000 | 4.773 | 4.968 |
| Model 4 | **Intercept** | 4.271 | 0.144 | 29.685 | 0.000 | 3.988 | 4.555 |
| Model 4 | **Slope** | 0.035 | 0.006 | 6.134 | 0.000 | 0.024 | 0.046 |
| Model 5 | **Intercept** | 8.799 | 0.407 | 21.642 | 0.000 | 7.999 | 9.599 |
| Model 5 | **Slope** | 0.607 | 0.016 | 37.527 | 0.000 | 0.576 | 0.639 |
| Model 6 | **Intercept** | 11.207 | 0.462 | 24.235 | 0.000 | 10.297 | 12.117 |
| Model 6 | **Slope** | 0.201 | 0.014 | 14.412 | 0.000 | 0.174 | 0.229 |
| Model 7 | **Intercept** | -129.989 | 17.061 | -7.619 | 0.000 | -163.564 | -96.414 |
| Model 7 | **Slope** | 3.909 | 0.473 | 8.262 | 0.000 | 2.978 | 4.840 |
| Model 8 | **Intercept** | 9.420 | 2.355 | 4.001 | 0.000 | 4.787 | 14.054 |
| Model 8 | **Slope** | 0.300 | 0.079 | 3.797 | 0.000 | 0.145 | 0.456 |

**Table S9.** Linear regression results for difference in relative percent averted in Scenario C vs. B, using the median horizon week of Scenario A as the independent variable.

| Model ID | Term | Estimate | Std. Error | t value | Pr(>\|t\|) | [0.025 | 0.975 |
| --- | --- | --- | --- | --- | --- | --- | --- |
| Model 1 | **Intercept** | 3.127 | 0.256 | 12.206 | 0.000 | 2.623 | 3.631 |
| Model 1 | **Slope** | -0.041 | 0.011 | -3.865 | 0.000 | -0.061 | -0.020 |
| Model 2 | **Intercept** | 1.680 | 0.110 | 15.243 | 0.000 | 1.463 | 1.896 |
| Model 2 | **Slope** | -0.017 | 0.004 | -3.836 | 0.000 | -0.026 | -0.008 |
| Model 3 | **Intercept** | -27.054 | 0.563 | -48.026 | 0.000 | -28.163 | -25.946 |
| Model 3 | **Slope** | 0.741 | 0.014 | 51.199 | 0.000 | 0.713 | 0.770 |
| Model 4 | **Intercept** | 0.944 | 0.037 | 25.376 | 0.000 | 0.870 | 1.017 |
| Model 4 | **Slope** | -0.040 | 0.001 | -27.367 | 0.000 | -0.043 | -0.037 |
| Model 5 | **Intercept** | 8.134 | 0.084 | 96.384 | 0.000 | 7.968 | 8.300 |
| Model 5 | **Slope** | -0.147 | 0.003 | -43.686 | 0.000 | -0.153 | -0.140 |
| Model 6 | **Intercept** | 2.333 | 0.468 | 4.981 | 0.000 | 1.411 | 3.255 |
| Model 6 | **Slope** | 0.003 | 0.014 | 0.183 | 0.855 | -0.025 | 0.030 |
| Model 7 | **Intercept** | -25.064 | 19.672 | -1.274 | 0.204 | -63.779 | 13.650 |
| Model 7 | **Slope** | 0.616 | 0.546 | 1.129 | 0.26 | -0.458 | 1.690 |
| Model 8 | **Intercept** | -0.351 | 0.465 | -0.755 | 0.451 | -1.266 | 0.564 |
| Model 8 | **Slope** | 0.067 | 0.016 | 4.280 | 0.000 | 0.036 | 0.098 |

**Table S10.** Linear regression results for difference in relative percent averted in Scenario E vs. D, using the median horizon week of Scenario A as the independent variable.

| Model ID | Term | Estimate | Std. Error | t value | Pr(>\|t\|) | [0.025 | 0.975 |
| --- | --- | --- | --- | --- | --- | --- | --- |
| Model 1 | **Intercept** | 2.153 | 0.336 | 6.417 | 0.000 | 1.493 | 2.813 |
| Model 1 | **Slope** | 0.007 | 0.014 | 0.522 | 0.602 | -0.020 | 0.034 |
| Model 2 | **Intercept** | 2.105 | 0.138 | 15.239 | 0.000 | 1.833 | 2.377 |
| Model 2 | **Slope** | -0.031 | 0.006 | -5.592 | 0.000 | -0.042 | -0.020 |
| Model 3 | **Intercept** | -25.990 | 0.963 | -26.977 | 0.000 | -27.886 | -24.094 |
| Model 3 | **Slope** | 0.759 | 0.025 | 30.674 | 0.000 | 0.711 | 0.808 |
| Model 4 | **Intercept** | 0.861 | 0.053 | 16.148 | 0.000 | 0.756 | 0.966 |
| Model 4 | **Slope** | -0.048 | 0.002 | -22.705 | 0.000 | -0.052 | -0.044 |
| Model 5 | **Intercept** | 8.456 | 0.146 | 57.856 | 0.000 | 8.168 | 8.743 |
| Model 5 | **Slope** | -0.103 | 0.006 | -17.766 | 0.000 | -0.115 | -0.092 |
| Model 6 | **Intercept** | 4.124 | 0.095 | 43.380 | 0.000 | 3.937 | 4.311 |
| Model 6 | **Slope** | -0.024 | 0.003 | -8.431 | 0.000 | -0.030 | -0.019 |
| Model 7 | **Intercept** | -5.958 | 20.886 | -0.285 | 0.776 | -47.061 | 35.144 |
| Model 7 | **Slope** | -0.042 | 0.579 | -0.072 | 0.943 | -1.182 | 1.098 |
| Model 8 | **Intercept** | -2.485 | 0.906 | -2.742 | 0.006 | -4.269 | -0.701 |
| Model 8 | **Slope** | 0.140 | 0.030 | 4.607 | 0.000 | 0.080 | 0.200 |

**Table S11.** Mixed effects analysis results for the ensemble model over the projection period (April 27, 2025 to April 25, 2026) using Scenario A as baseline, rest of the scenarios (B-E) as fixed effects and location (states) and teams (models) as random effects. The three dependent variables considered are: total, summer, and winter wave sizes per 100k population. Absolute values of the entries in the Coef. column for the scenario variables corresponds to the average number of hospitalizations averted per 100k population compared to Scenario A.

| Variable | Coef. | Std.Err. | z | P>\|z\| | [0.025 | 0.975] |
| --- | --- | --- | --- | --- | --- | --- |
| **Intercept** | 211.601 | 5.113 | 41.388 | 0.000 | 201.581 | 221.622 |
| **scenario_id[T.B-2025-04-01]** | -30.091 | 0.288 | -104.483 | 0.000 | -30.655 | -29.526 |
| **scenario_id[T.C-2025-04-01]** | -33.750 | 0.288 | -117.189 | 0.000 | -34.314 | -33.185 |
| scenario_id[T.D-2025-04-01] | -38.244 | 0.288 | -132.793 | 0.000 | -38.808 | -37.679 |
| scenario_id[T.E-2025-04-01] | -43.045 | 0.288 | -149.465 | 0.000 | -43.610 | -42.481 |
| Group Var | 585.783 | 6.695 |  |  |  |  |
| Team Var | 6135.740 | 7.629 |  |  |  |  |

| Variable | Coef. | Std.Err. | z | P>\|z\| | [0.025 | 0.975] |
| --- | --- | --- | --- | --- | --- | --- |
| Intercept | 89.363 | 3.012 | 29.664 | 0.000 | 83.458 | 95.267 |
| scenario_id[T.B-2025-04-01] | -3.730 | 0.178 | -20.998 | 0.000 | -4.078 | -3.382 |
| scenario_id[T.C-2025-04-01] | -9.029 | 0.178 | -50.824 | 0.000 | -9.377 | -8.680 |
| scenario_id[T.D-2025-04-01] | -4.365 | 0.178 | -24.572 | 0.000 | -4.713 | -4.017 |
| scenario_id[T.E-2025-04-01] | -11.212 | 0.178 | -63.113 | 0.000 | -11.560 | -10.864 |
| Group Var | 51.146 |  |  |  |  |  |
| Team Var | 3343.503 | 6.459 |  |  |  |  |

**Table S11.** Mixed effects analysis results for the ensemble model over the projection period (April 27, 2025 to April 25, 2026) using Scenario A as baseline, rest of the scenarios (B-E) as fixed effects and location (states) and teams (models) as random effects.
| Variable | Coef. | Std.Err. | z | P>\|z\| | [0.025 | 0.975] |
| --- | --- | --- | --- | --- | --- | --- |
| Intercept | 122.222 | 2.941 | 41.555 | 0.000 | 116.458 | 127.987 |
| scenario_id[T.B-2025-04-01] | -26.360 | 0.217 | -121.202 | 0.000 | -26.787 | -25.934 |
| scenario_id[T.C-2025-04-01] | -24.721 | 0.217 | -113.665 | 0.000 | -25.147 | -24.295 |
| scenario_id[T.D-2025-04-01] | -33.879 | 0.217 | -155.770 | 0.000 | -34.305 | -33.452 |
| scenario_id[T.E-2025-04-01] | -31.833 | 0.217 | -146.366 | 0.000 | -32.260 | -31.407 |
| **Group Var** | 279.346 | 1.765 |  |  |  |  |
| **Team Var** | 1345.835 | 2.287 |  |  |  |  |

**Table S12.** Mean difference across teams in scenario effects on the total, summer, and winter wave sizes between winter and summer leaning states over the projection period (April 27, 2025 to April 25, 2026). The difference is the number of excess hospitalizations averted per 100k individuals in the winter leaning states compared to the summer ones.

| **Scenario** | **Difference**<br>**in effect on total epidemic size**<br>**(mean±std)** | **Difference**<br>**in effect on summer wave size**<br>**(mean±std)** | **Difference**<br>**in effect on winter wave size**<br>**(mean±std)** |
| --- | --- | --- | --- |
| B | 12.49±10.71 | 1.35±1.07 | 11.91±9.47 |
| C | 12.87±11.27 | 2.58±1.83 | 12.74±9.41 |
| D | 15.64±13.99 | 1.69±1.53 | 14.70±12.50 |
| E | 16.46±15.67 | 3.72±2.79 | 15.78±12.95 |

**Table S13.** Mean difference across teams in the scenario effects on the summer wave size between scenarios C and B (left) and D and B (right) for both summer and winter leaning states over the projection period (April 27, 2025 to April 25, 2026). The difference is the number of excess hospitalizations averted per 100k individuals.

| **Scenario C - Scenario B**<br>**(mean±std)** |  | **Scenario D - Scenario B**<br>**(mean±std)** |  |
| --- | --- | --- | --- |
| **Summer leaning** | **Winter leaning** | **Summer leaning** | **Winter leaning** |
| 7.57±4.67 | 5.80±4.94 | 0.78±0.43 | 0.87±0.83 |

**Table S14.** Mean difference across teams in the scenario effects on the winter wave size between scenarios C and B (left) and D and B (right) for both summer and winter leaning locations over the projection period (April 27, 2025 to April 25, 2026). The difference is the number of excess hospitalizations averted per 100k individuals.

| **Scenario C - Scenario B**<br>**(mean±std)** |  | **Scenario D - Scenario B**<br>**(mean±std)** |  |
| --- | --- | --- | --- |
| **Summer leaning** | **Winter leaning** | **Summer leaning** | **Winter leaning** |
| -1.99±2.14 | -1.31±2.14 | 5.30±2.77 | 8.89±6.27 |

**Table S15.**
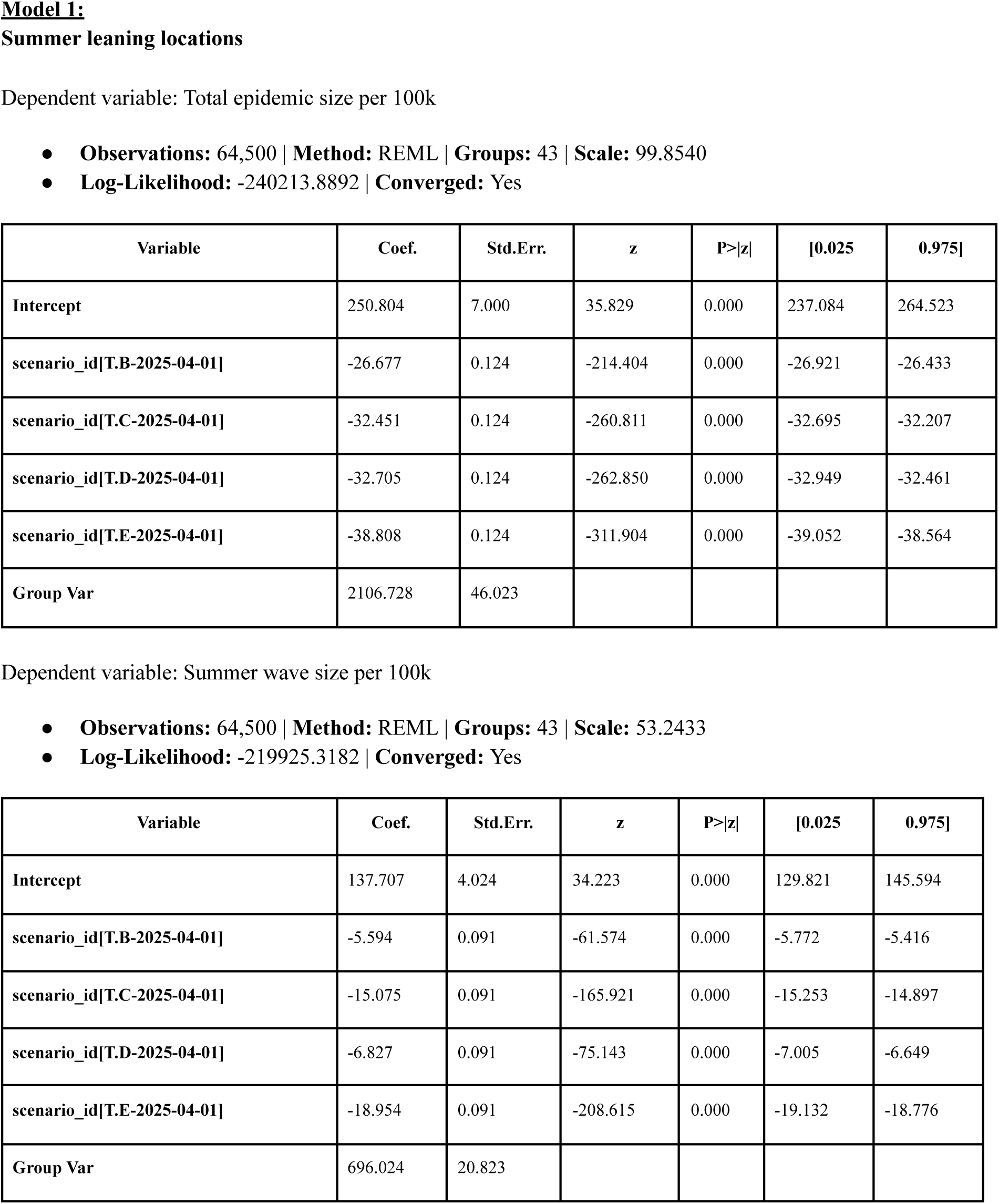
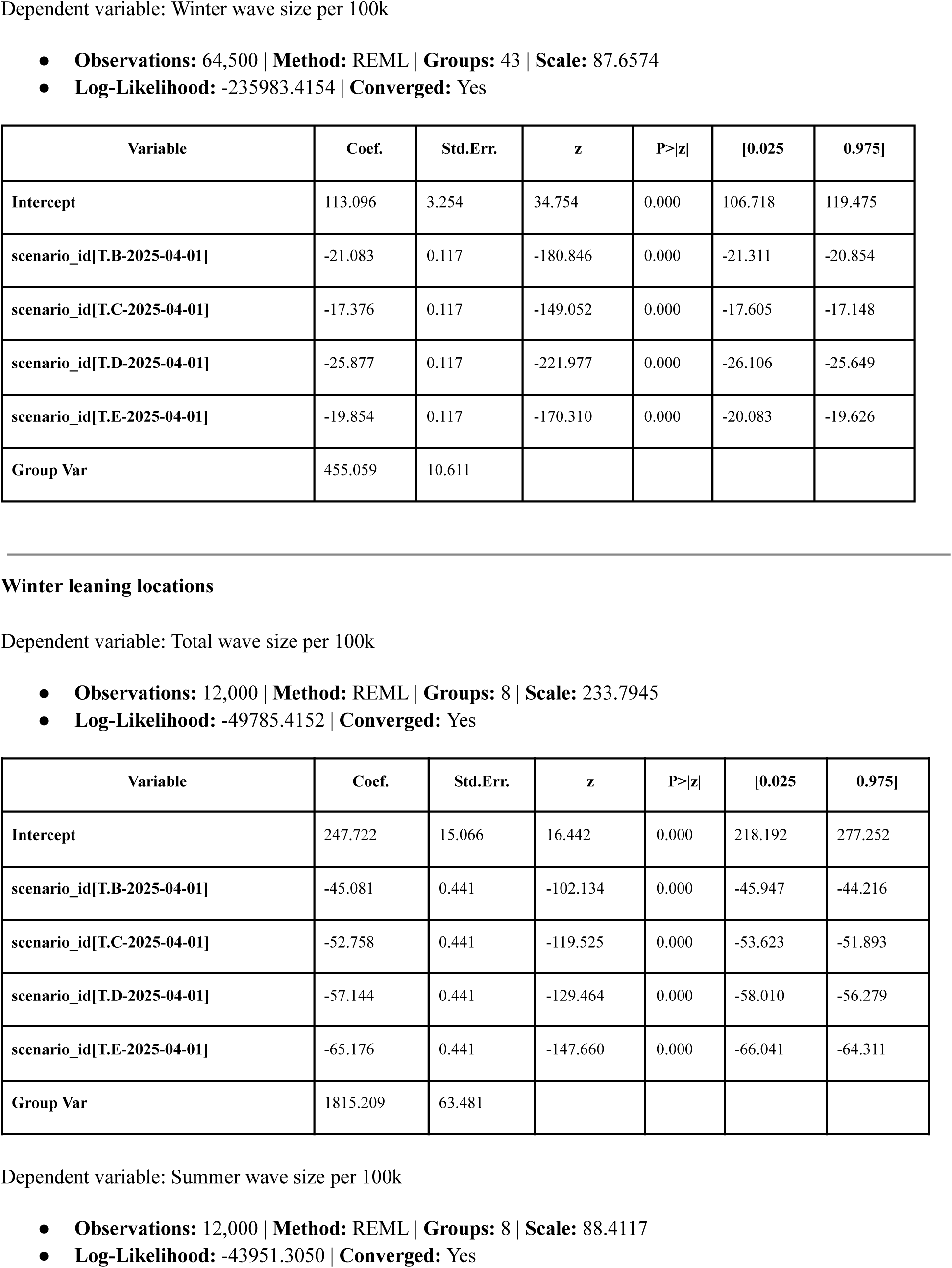
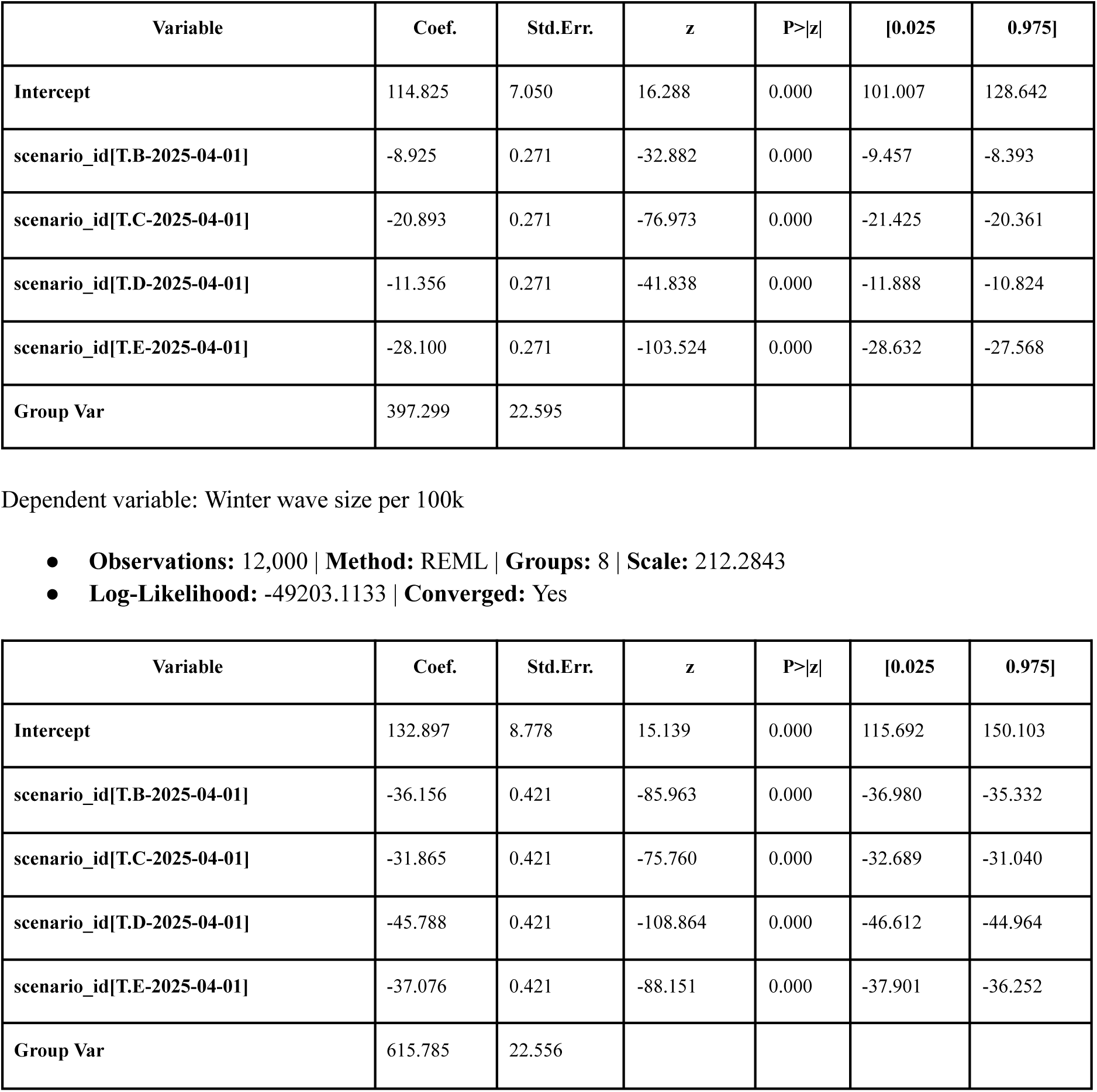
Mixed effects analysis results for Model 1 over the projection period (April 27, 2025 to April 25, 2026) using Scenario A as baseline, rest of the scenarios (B-E) as fixed effects and location (states) as random effects for both summer and winter leaning locations. The three dependent variables considered are: total, summer, and winter wave sizes per 100k population. Absolute values of the entries in the Coef. column for the scenario variables corresponds to the average number of hospitalizations averted per 100k population compared to Scenario A.

| Variable | Coef. | Std.Err. | z | P>\|z\| | [0.025 | 0.975] |
| --- | --- | --- | --- | --- | --- | --- |
| Intercept | 250.804 | 7.000 | 35.829 | 0.000 | 237.084 | 264.523 |
| scenario_id[T.B-2025-04-01] | -26.677 | 0.124 | -214.404 | 0.000 | -26.921 | -26.433 |
| scenario_id[T.C-2025-04-01] | -32.451 | 0.124 | -260.811 | 0.000 | -32.695 | -32.207 |
| scenario_id[T.D-2025-04-01] | -32.705 | 0.124 | -262.850 | 0.000 | -32.949 | -32.461 |
| scenario_id[T.E-2025-04-01] | -38.808 | 0.124 | -311.904 | 0.000 | -39.052 | -38.564 |
| Group Var | 2106.728 | 46.023 |  |  |  |  |

| Variable | Coef. | Std.Err. | z | P>\|z\| | [0.025 | 0.975] |
| --- | --- | --- | --- | --- | --- | --- |
| Intercept | 137.707 | 4.024 | 34.223 | 0.000 | 129.821 | 145.594 |
| scenario_id[T.B-2025-04-01] | -5.594 | 0.091 | -61.574 | 0.000 | -5.772 | -5.416 |
| scenario_id[T.C-2025-04-01] | -15.075 | 0.091 | -165.921 | 0.000 | -15.253 | -14.897 |
| scenario_id[T.D-2025-04-01] | -6.827 | 0.091 | -75.143 | 0.000 | -7.005 | -6.649 |
| scenario_id[T.E-2025-04-01] | -18.954 | 0.091 | -208.615 | 0.000 | -19.132 | -18.776 |
| Group Var | 696.024 | 20.823 |  |  |  |  |
| Intercept | 113.096 | 3.254 | 34.754 | 0.000 | 106.718 | 119.475 |
| scenario_id[T.B-2025-04-01] | -21.083 | 0.117 | -180.846 | 0.000 | -21.311 | -20.854 |
| scenario_id[T.C-2025-04-01] | -17.376 | 0.117 | -149.052 | 0.000 | -17.605 | -17.148 |
| scenario_id[T.D-2025-04-01] | -25.877 | 0.117 | -221.977 | 0.000 | -26.106 | -25.649 |
| scenario_id[T.E-2025-04-01] | -19.854 | 0.117 | -170.310 | 0.000 | -20.083 | -19.626 |
| Group Var | 455.059 | 10.611 |  |  |  |  |

| Variable | Coef. | Std.Err. | z | P>\|z\| | [0.025 | 0.975] |
| --- | --- | --- | --- | --- | --- | --- |
| Intercept | 247.722 | 15.066 | 16.442 | 0.000 | 218.192 | 277.252 |
| scenario_id[T.B-2025-04-01] | -45.081 | 0.441 | -102.134 | 0.000 | -45.947 | -44.216 |
| scenario_id[T.C-2025-04-01] | -52.758 | 0.441 | -119.525 | 0.000 | -53.623 | -51.893 |
| scenario_id[T.D-2025-04-01] | -57.144 | 0.441 | -129.464 | 0.000 | -58.010 | -56.279 |
| scenario_id[T.E-2025-04-01] | -65.176 | 0.441 | -147.660 | 0.000 | -66.041 | -64.311 |
| Group Var | 1815.209 | 63.481 |  |  |  |  |
| Intercept | 114.825 | 7.050 | 16.288 | 0.000 | 101.007 | 128.642 |
| scenario_id[T.B-2025-04-01] | -8.925 | 0.271 | -32.882 | 0.000 | -9.457 | -8.393 |
| scenario_id[T.C-2025-04-01] | -20.893 | 0.271 | -76.973 | 0.000 | -21.425 | -20.361 |
| scenario_id[T.D-2025-04-01] | -11.356 | 0.271 | -41.838 | 0.000 | -11.888 | -10.824 |
| scenario_id[T.E-2025-04-01] | -28.100 | 0.271 | -103.524 | 0.000 | -28.632 | -27.568 |
| Group Var | 397.299 | 22.595 |  |  |  |  |

**Table S15.** Mixed effects analysis results for Model 1 over the projection period (April 27, 2025 to April 25, 2026) using Scenario A as baseline, rest of the scenarios (B-E) as fixed effects and location (states) as random effects for both summer and winter leaning locations.
| Variable | Coef. | Std.Err. | z | P>\|z\| | [0.025 | 0.975] |
| --- | --- | --- | --- | --- | --- | --- |
| Intercept | 132.897 | 8.778 | 15.139 | 0.000 | 115.692 | 150.103 |
| scenario_id[T.B-2025-04-01] | -36.156 | 0.421 | -85.963 | 0.000 | -36.980 | -35.332 |
| scenario_id[T.C-2025-04-01] | -31.865 | 0.421 | -75.760 | 0.000 | -32.689 | -31.040 |
| scenario_id[T.D-2025-04-01] | -45.788 | 0.421 | -108.864 | 0.000 | -46.612 | -44.964 |
| scenario_id[T.E-2025-04-01] | -37.076 | 0.421 | -88.151 | 0.000 | -37.901 | -36.252 |
| Group Var | 615.785 | 22.556 |  |  |  |  |

**Table S16.**
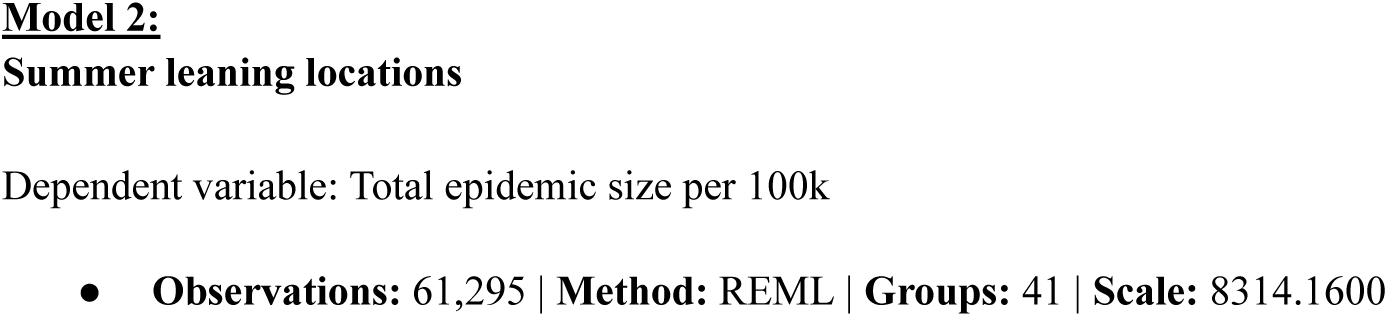
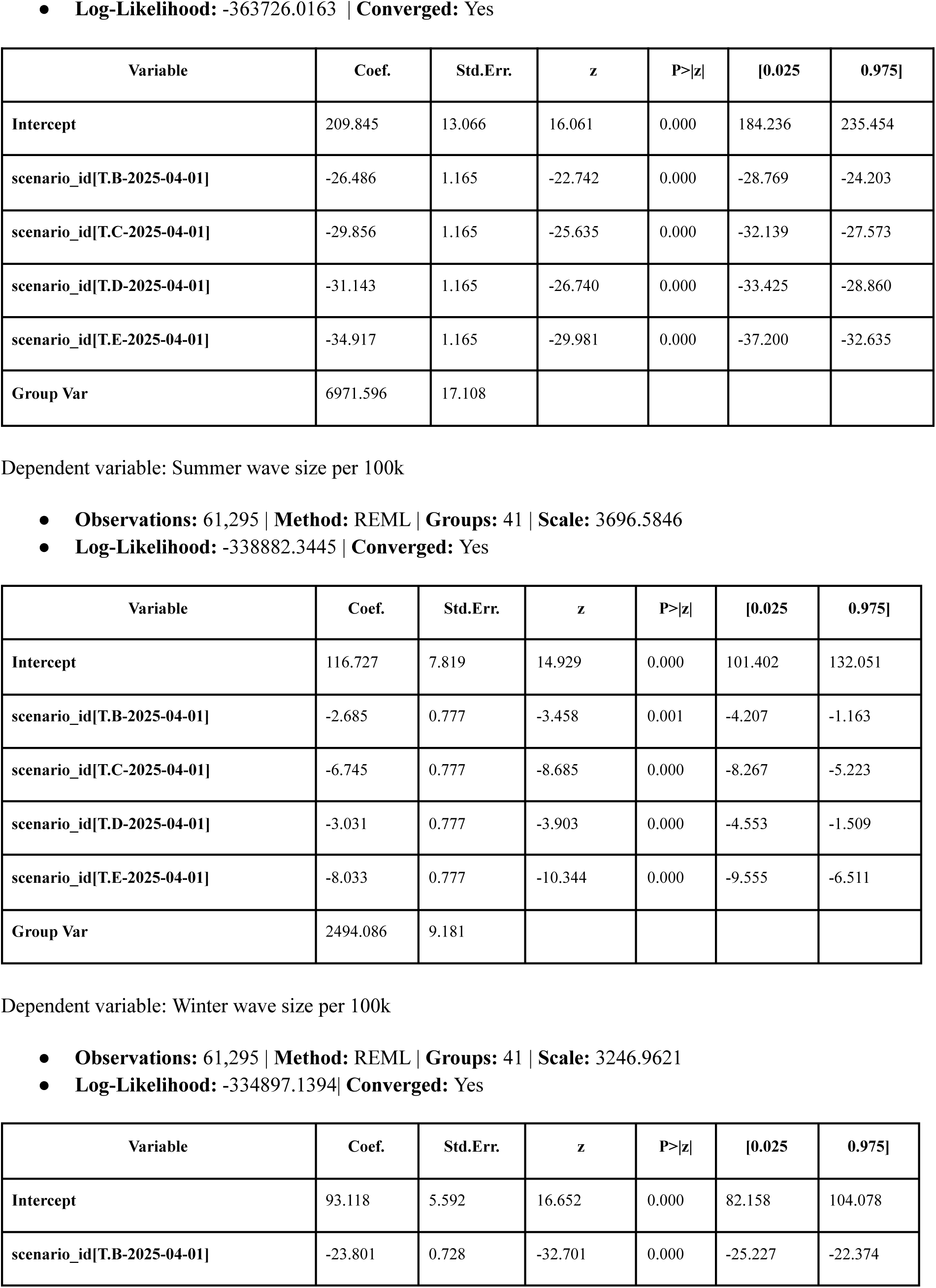
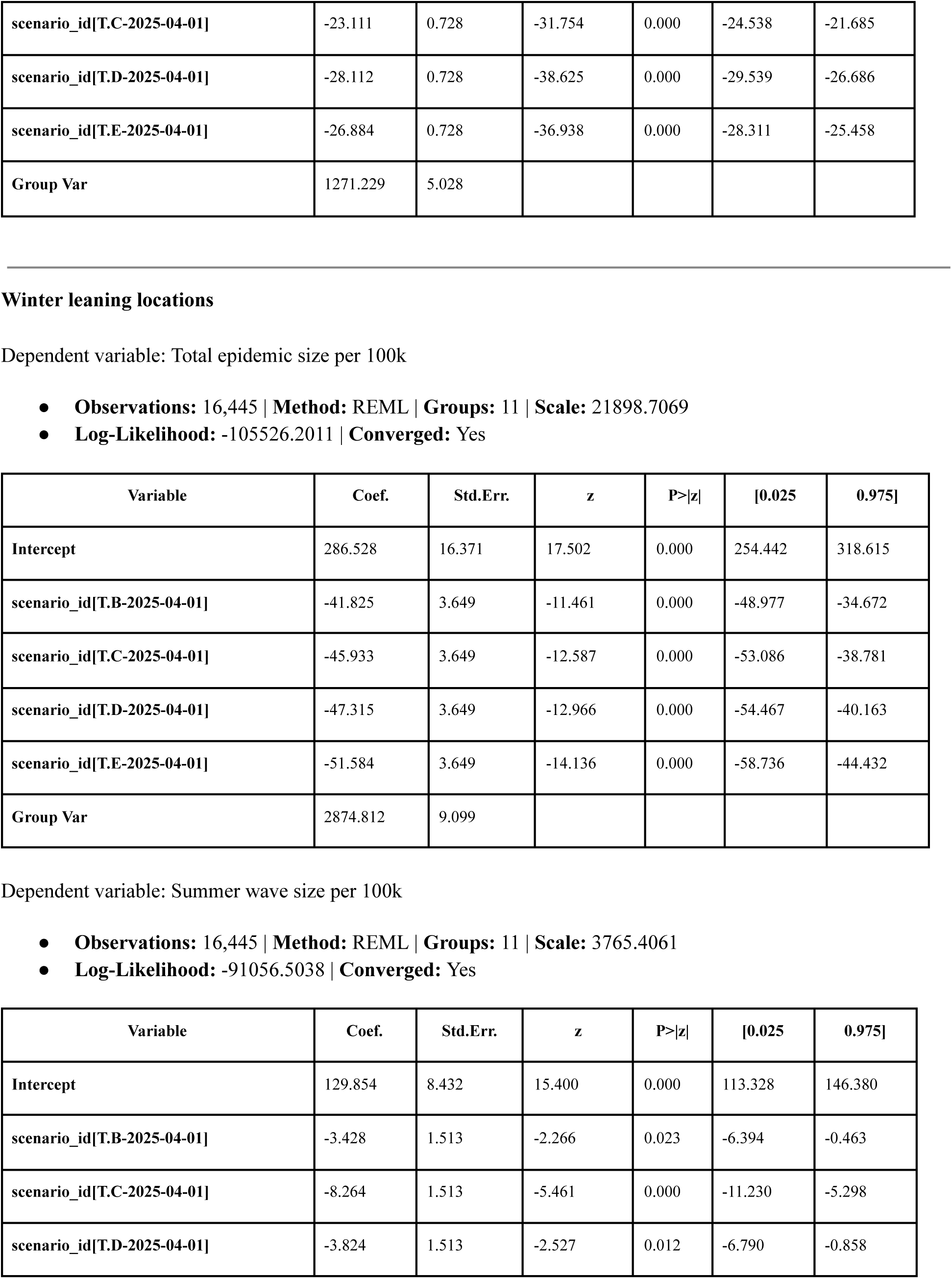
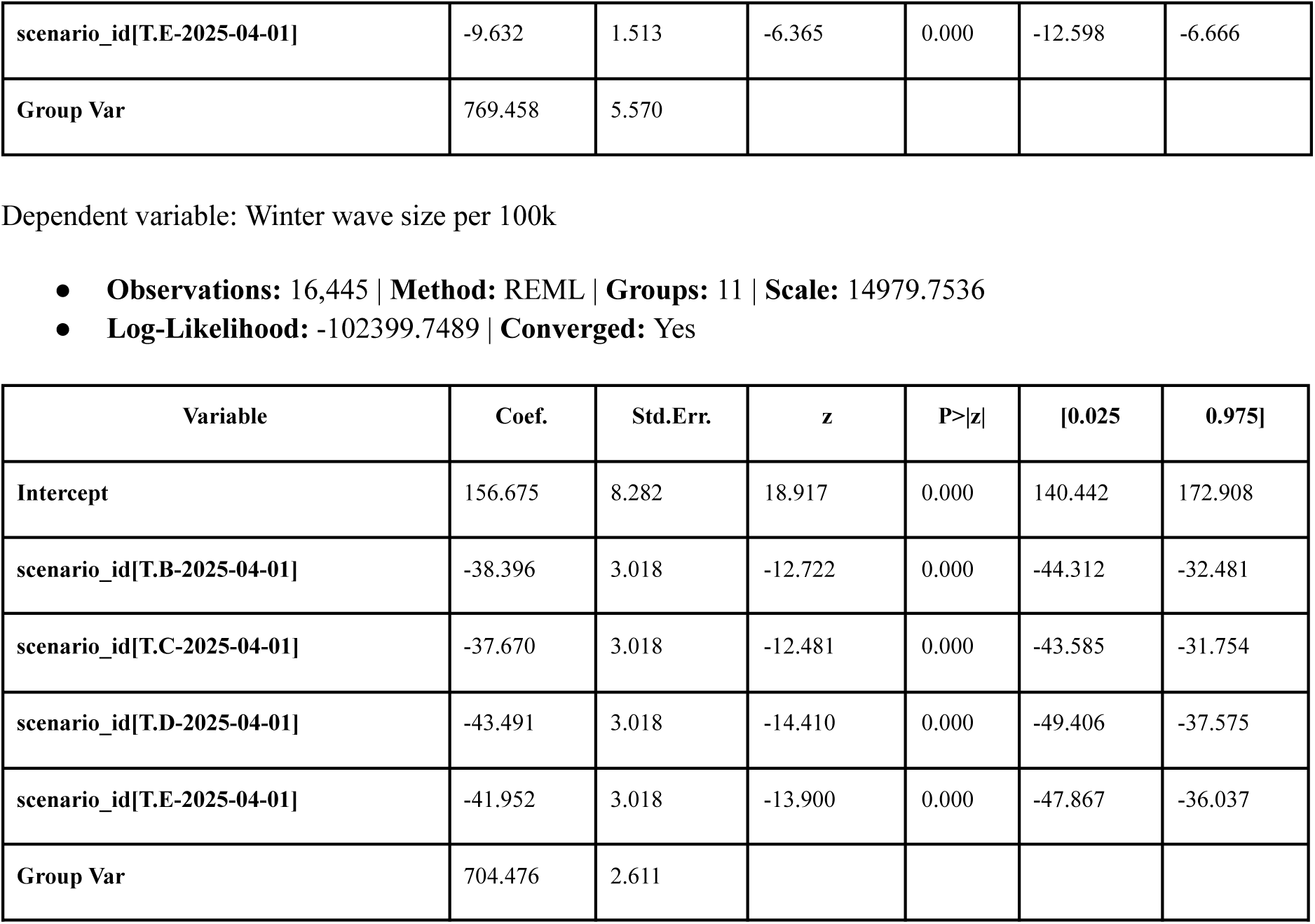
Mixed effects analysis results for Model 2 over the projection period (April 27, 2025 to April 25, 2026) using Scenario A as baseline, rest of the scenarios (B-E) as fixed effects and location (states) as random effects for both summer and winter leaning locations. The three dependent variables considered are: total, summer, and winter wave sizes per 100k population. Absolute values of the entries in the Coef. column for the scenario variables corresponds to the average number of hospitalizations averted per 100k population compared to Scenario A.

**Table S17.** Mixed effects analysis results for Model 3 over the projection period (April 27, 2025 to April 25, 2026) using Scenario A as baseline, rest of the scenarios (B-E) as fixed effects and location (states) as random effects for winter leaning locations (model has no summer leaning locations). The three dependent variables considered are: total, summer, and winter wave sizes per 100k population. Absolute values of the entries in the Coef. column for the scenario variables corresponds to the average number of hospitalizations averted per 100k population compared to Scenario A.

| Variable | Coef. | Std.Err. | z | P>\|z\| | [0.025 | 0.975] |
| --- | --- | --- | --- | --- | --- | --- |
| Intercept | 223.509 | 5.967 | 37.455 | 0.000 | 211.813 | 235.204 |
| scenario_id[T.B-2025-04-01] | -53.063 | 0.695 | -76.332 | 0.000 | -54.425 | -51.700 |
| scenario_id[T.C-2025-04-01] | -55.654 | 0.695 | -80.060 | 0.000 | -57.017 | -54.292 |
| scenario_id[T.D-2025-04-01] | -71.003 | 0.695 | -102.140 | 0.000 | -72.366 | -69.641 |
| scenario_id[T.E-2025-04-01] | -77.439 | 0.695 | -111.397 | 0.000 | -78.801 | -76.076 |
| Group Var | 1839.142 | 5.921 |  |  |  |  |

| Variable | Coef. | Std.Err. | z | P>\|z\| | [0.025 | 0.975] |
| --- | --- | --- | --- | --- | --- | --- |
| Intercept | 28.889 | 2.198 | 13.143 | 0.000 | 24.581 | 33.197 |
| scenario_id[T.B-2025-04-01] | -0.696 | 0.293 | -2.374 | 0.018 | -1.271 | -0.121 |
| scenario_id[T.C-2025-04-01] | -3.233 | 0.293 | -11.027 | 0.000 | -3.807 | -2.658 |
| scenario_id[T.D-2025-04-01] | -0.831 | 0.293 | -2.833 | 0.005 | -1.405 | -0.256 |
| scenario_id[T.E-2025-04-01] | -3.970 | 0.293 | -13.541 | 0.000 | -4.544 | -3.395 |
| Group Var | 248.999 | 1.902 |  |  |  |  |

**Table S17.** Mixed effects analysis results for Model 3 over the projection period (April 27, 2025 to April 25, 2026) using Scenario A as baseline, rest of the scenarios (B-E) as fixed effects and location (states) as random effects for winter leaning locations (model has no summer leaning locations).
| Variable | Coef. | Std.Err. | z | P>\|z\| | [0.025 | 0.975] |
| --- | --- | --- | --- | --- | --- | --- |
| Intercept | 194.620 | 4.531 | 42.957 | 0.000 | 185.740 | 203.500 |
| scenario_id[T.B-2025-04-01] | -52.367 | 0.557 | -93.937 | 0.000 | -53.459 | -51.274 |
| scenario_id[T.C-2025-04-01] | -52.422 | 0.557 | -94.036 | 0.000 | -53.514 | -51.329 |
| scenario_id[T.D-2025-04-01] | -70.173 | 0.557 | -125.879 | 0.000 | -71.265 | -69.080 |
| scenario_id[T.E-2025-04-01] | -73.469 | 0.557 | -131.792 | 0.000 | -74.562 | -72.376 |
| Group Var | 1059.298 | 4.270 |  |  |  |  |

**Table S18.**
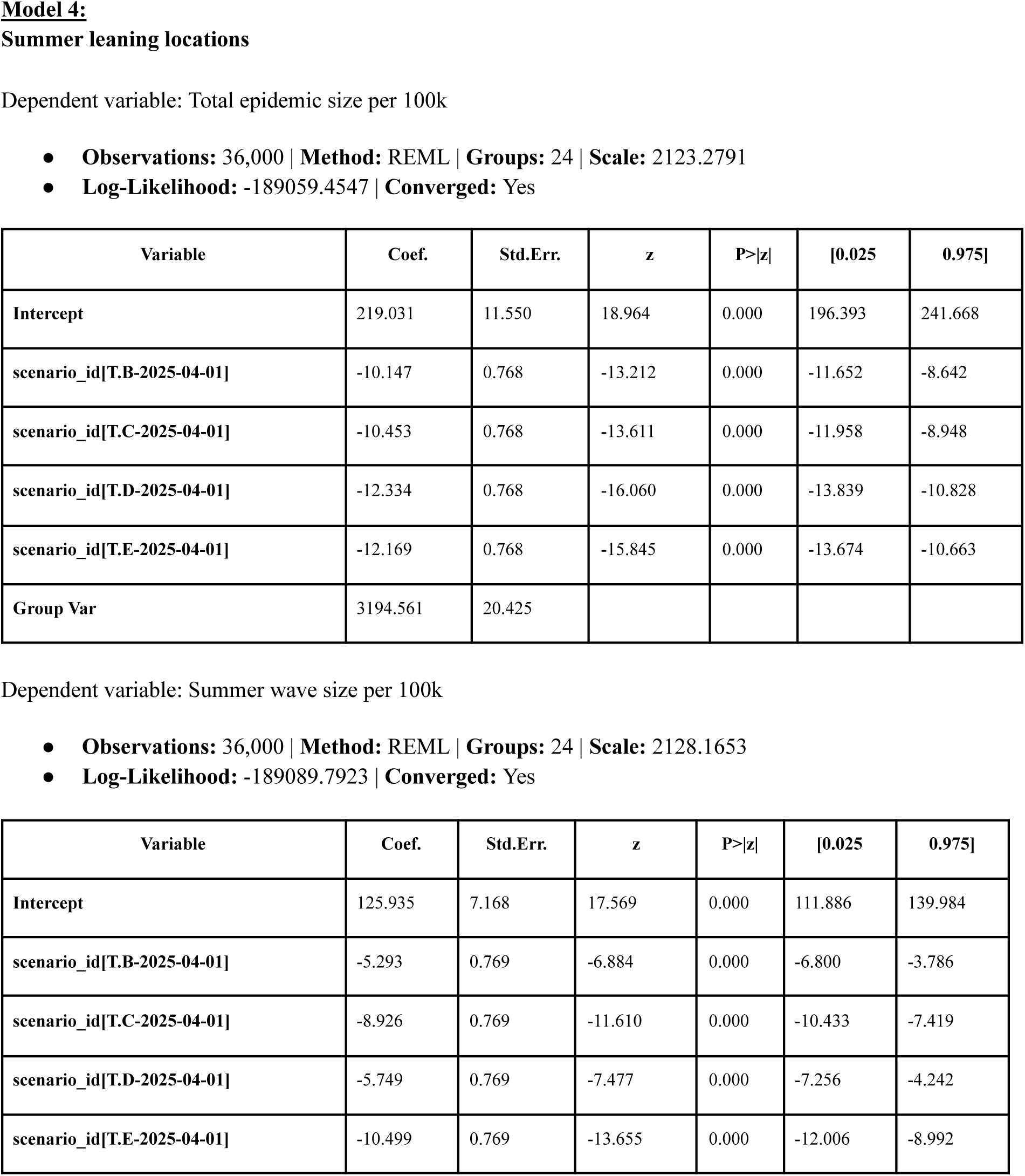
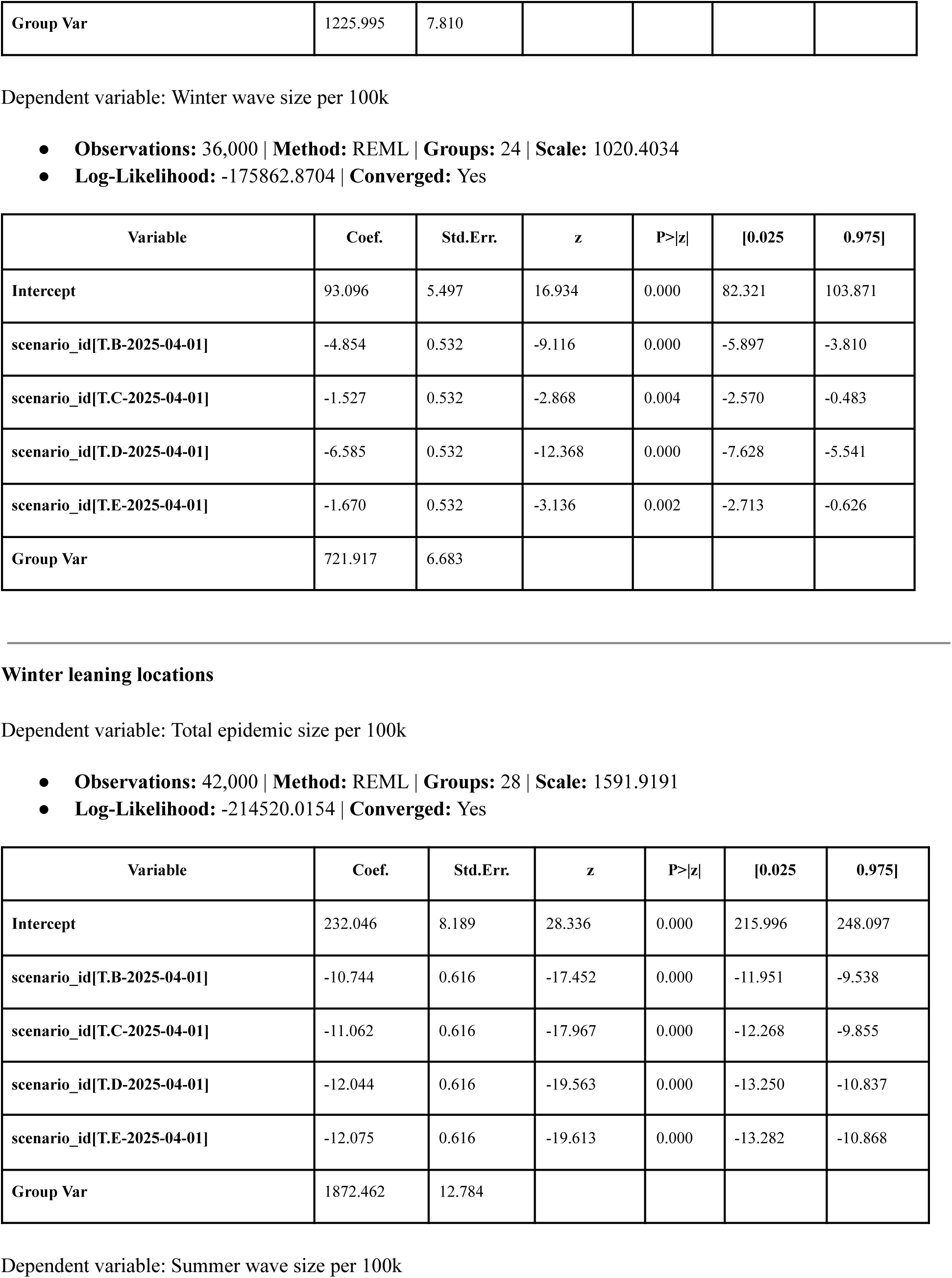
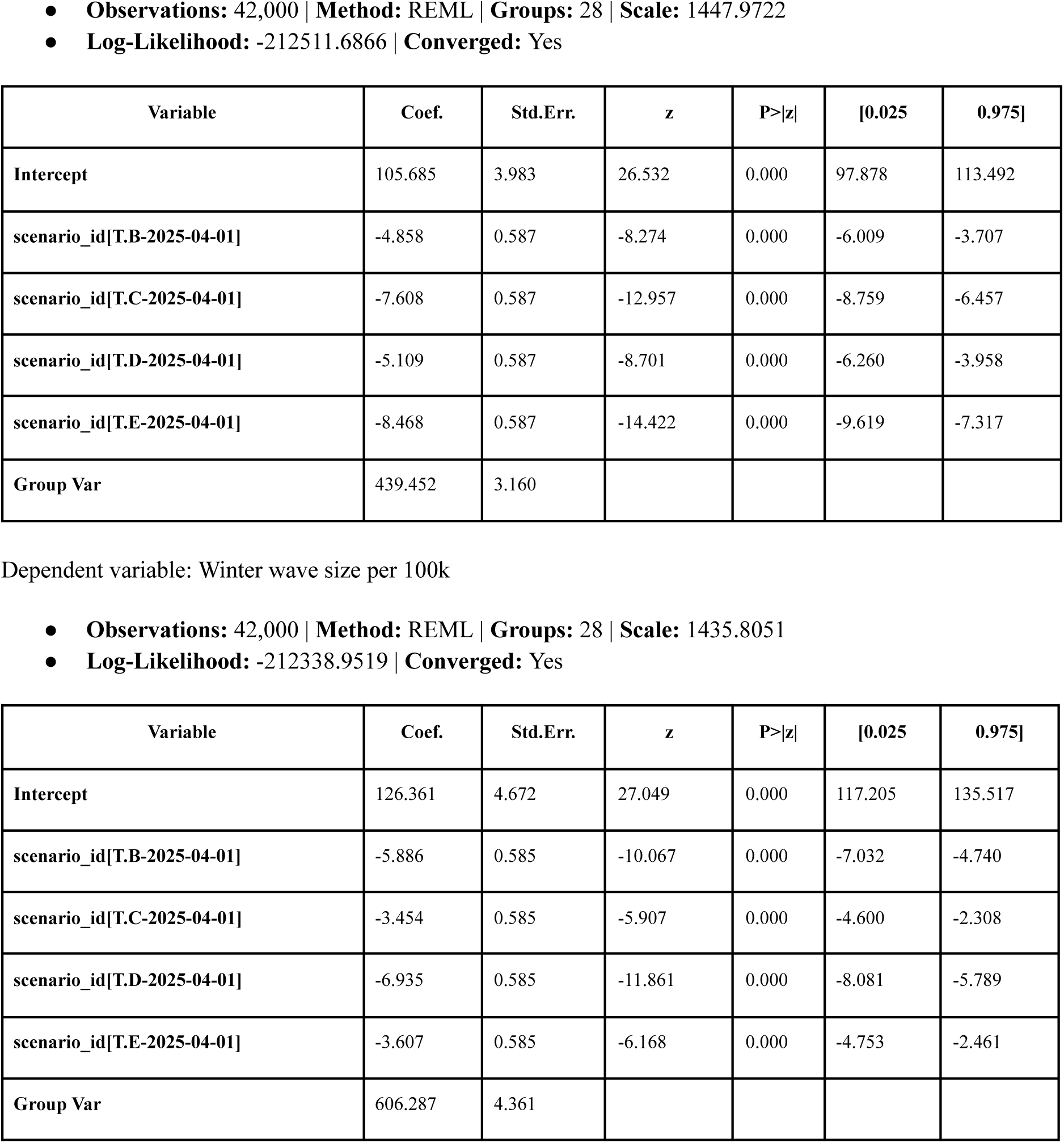
Mixed effects analysis results for Model 4 over the projection period (April 27, 2025 to April 25, 2026) using Scenario A as baseline, rest of the scenarios (B-E) as fixed effects and location (states) as random effects for both summer and winter leaning locations. The three dependent variables considered are: total, summer, and winter wave sizes per 100k population. Absolute values of the entries in the Coef. column for the scenario variables corresponds to the average number of hospitalizations averted per 100k population compared to Scenario A.

| Variable | Coef. | Std.Err. | z | P>\|z\| | [0.025 | 0.975] |
| --- | --- | --- | --- | --- | --- | --- |
| Intercept | 219.031 | 11.550 | 18.964 | 0.000 | 196.393 | 241.668 |
| scenario_id[T.B-2025-04-01] | -10.147 | 0.768 | -13.212 | 0.000 | -11.652 | -8.642 |
| scenario_id[T.C-2025-04-01] | -10.453 | 0.768 | -13.611 | 0.000 | -11.958 | -8.948 |
| scenario_id[T.D-2025-04-01] | -12.334 | 0.768 | -16.060 | 0.000 | -13.839 | -10.828 |
| scenario_id[T.E-2025-04-01] | -12.169 | 0.768 | -15.845 | 0.000 | -13.674 | -10.663 |
| Group Var | 3194.561 | 20.425 |  |  |  |  |

| Variable | Coef. | Std.Err. | z | P>\|z\| | [0.025 | 0.975] |
| --- | --- | --- | --- | --- | --- | --- |
| Intercept | 125.935 | 7.168 | 17.569 | 0.000 | 111.886 | 139.984 |
| scenario_id[T.B-2025-04-01] | -5.293 | 0.769 | -6.884 | 0.000 | -6.800 | -3.786 |
| scenario_id[T.C-2025-04-01] | -8.926 | 0.769 | -11.610 | 0.000 | -10.433 | -7.419 |
| scenario_id[T.D-2025-04-01] | -5.749 | 0.769 | -7.477 | 0.000 | -7.256 | -4.242 |
| scenario_id[T.E-2025-04-01] | -10.499 | 0.769 | -13.655 | 0.000 | -12.006 | -8.992 |
| Group Var | 1225.995 | 7.810 |  |  |  |  |

| Variable | Coef. | Std.Err. | z | P>\|z\| | [0.025 | 0.975] |
| --- | --- | --- | --- | --- | --- | --- |
| Intercept | 93.096 | 5.497 | 16.934 | 0.000 | 82.321 | 103.871 |
| scenario_id[T.B-2025-04-01] | -4.854 | 0.532 | -9.116 | 0.000 | -5.897 | -3.810 |
| scenario_id[T.C-2025-04-01] | -1.527 | 0.532 | -2.868 | 0.004 | -2.570 | -0.483 |
| scenario_id[T.D-2025-04-01] | -6.585 | 0.532 | -12.368 | 0.000 | -7.628 | -5.541 |
| scenario_id[T.E-2025-04-01] | -1.670 | 0.532 | -3.136 | 0.002 | -2.713 | -0.626 |
| Group Var | 721.917 | 6.683 |  |  |  |  |

| Variable | Coef. | Std.Err. | z | P>\|z\| | [0.025 | 0.975] |
| --- | --- | --- | --- | --- | --- | --- |
| Intercept | 232.046 | 8.189 | 28.336 | 0.000 | 215.996 | 248.097 |
| scenario_id[T.B-2025-04-01] | -10.744 | 0.616 | -17.452 | 0.000 | -11.951 | -9.538 |
| scenario_id[T.C-2025-04-01] | -11.062 | 0.616 | -17.967 | 0.000 | -12.268 | -9.855 |
| scenario_id[T.D-2025-04-01] | -12.044 | 0.616 | -19.563 | 0.000 | -13.250 | -10.837 |
| scenario_id[T.E-2025-04-01] | -12.075 | 0.616 | -19.613 | 0.000 | -13.282 | -10.868 |
| Group Var | 1872.462 | 12.784 |  |  |  |  |
| Intercept | 105.685 | 3.983 | 26.532 | 0.000 | 97.878 | 113.492 |
| scenario_id[T.B-2025-04-01] | -4.858 | 0.587 | -8.274 | 0.000 | -6.009 | -3.707 |
| scenario_id[T.C-2025-04-01] | -7.608 | 0.587 | -12.957 | 0.000 | -8.759 | -6.457 |
| scenario_id[T.D-2025-04-01] | -5.109 | 0.587 | -8.701 | 0.000 | -6.260 | -3.958 |
| scenario_id[T.E-2025-04-01] | -8.468 | 0.587 | -14.422 | 0.000 | -9.619 | -7.317 |
| Group Var | 439.452 | 3.160 |  |  |  |  |

**Table S18.** Mixed effects analysis results for Model 4 over the projection period (April 27, 2025 to April 25, 2026) using Scenario A as baseline, rest of the scenarios (B-E) as fixed effects and location (states) as random effects for both summer and winter leaning locations.
| Variable | Coef. | Std.Err. | z | P>\|z\| | [0.025 | 0.975] |
| --- | --- | --- | --- | --- | --- | --- |
| Intercept | 126.361 | 4.672 | 27.049 | 0.000 | 117.205 | 135.517 |
| scenario_id[T.B-2025-04-01] | -5.886 | 0.585 | -10.067 | 0.000 | -7.032 | -4.740 |
| scenario_id[T.C-2025-04-01] | -3.454 | 0.585 | -5.907 | 0.000 | -4.600 | -2.308 |
| scenario_id[T.D-2025-04-01] | -6.935 | 0.585 | -11.861 | 0.000 | -8.081 | -5.789 |
| scenario_id[T.E-2025-04-01] | -3.607 | 0.585 | -6.168 | 0.000 | -4.753 | -2.461 |
| Group Var | 606.287 | 4.361 |  |  |  |  |

**Table S19.**
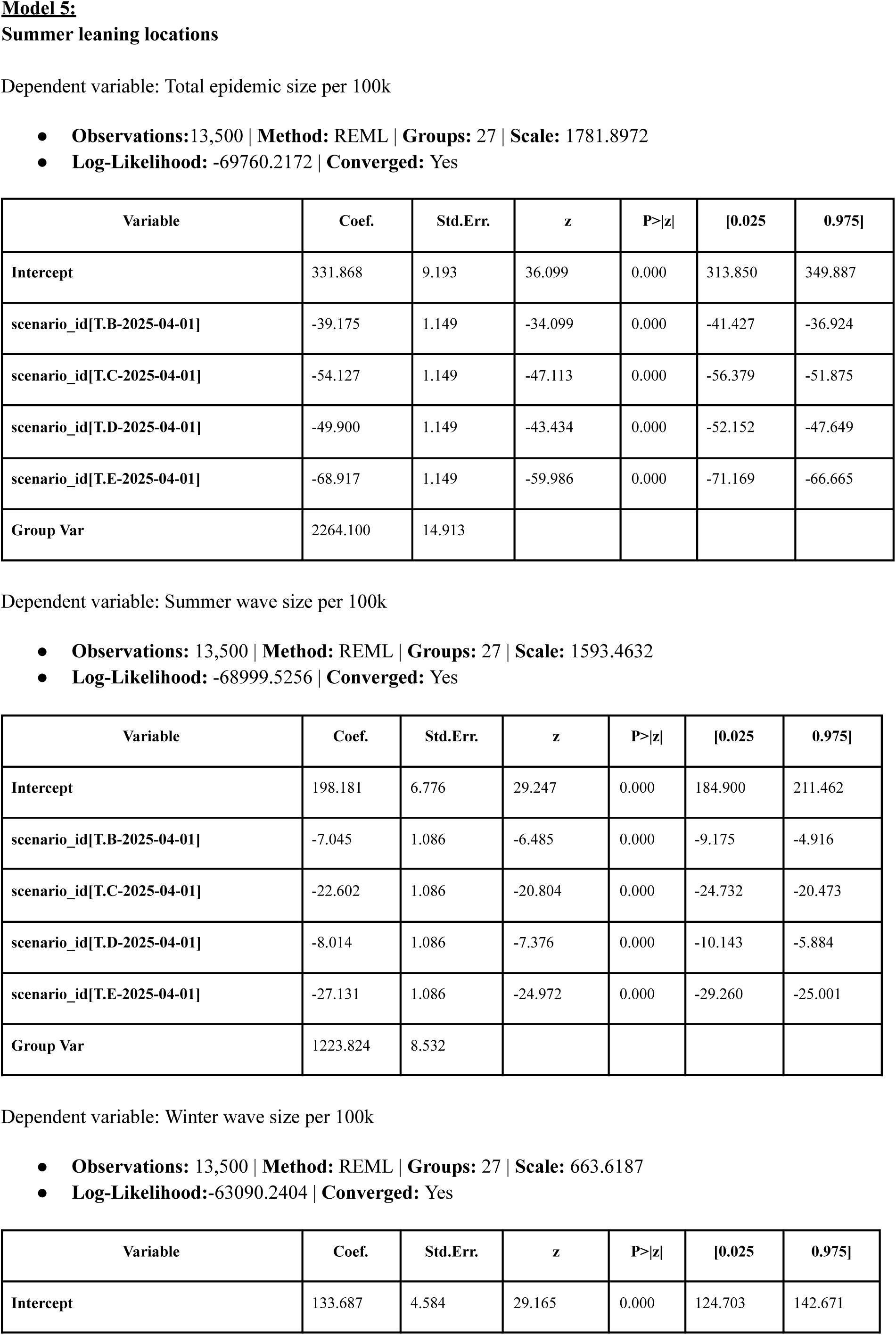
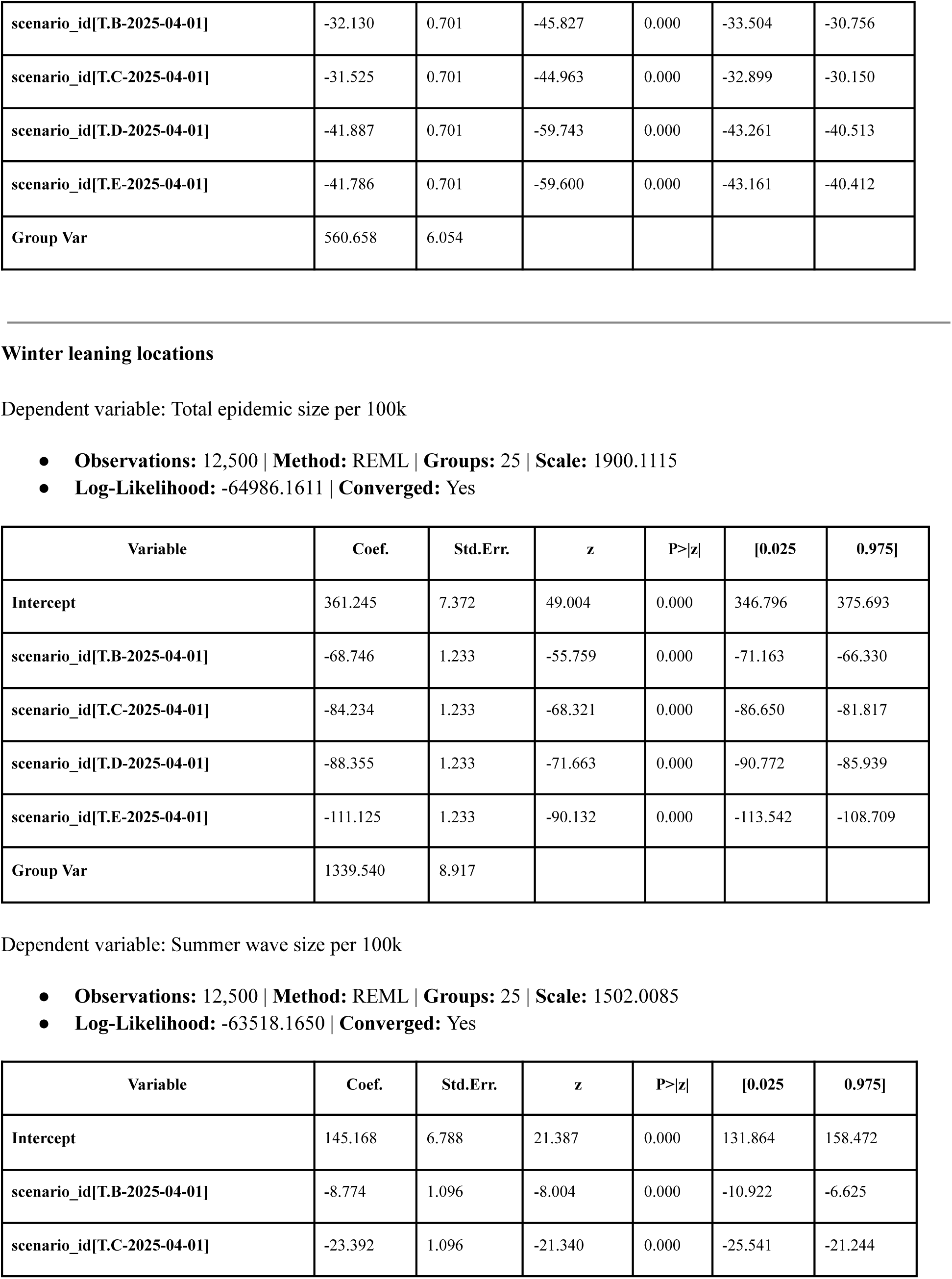
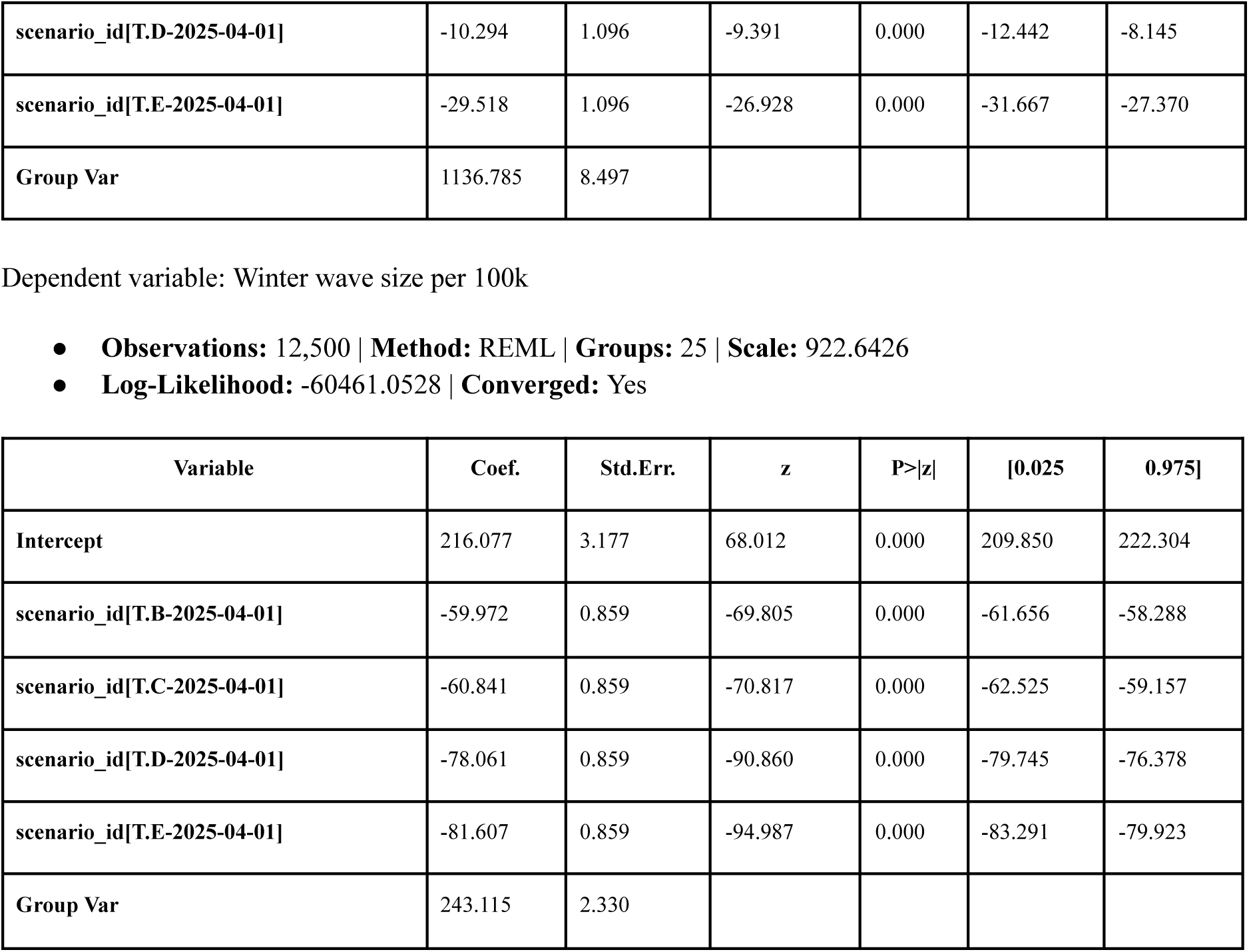
Mixed effects analysis results for Model 5 over the projection period (April 27, 2025 to April 25, 2026) using Scenario A as baseline, rest of the scenarios (B-E) as fixed effects and location (states) as random effects for both summer and winter leaning locations. The three dependent variables considered are: total, summer, and winter wave sizes per 100k population. Absolute values of the entries in the Coef. column for the scenario variables corresponds to the average number of hospitalizations averted per 100k population compared to Scenario A.

| Variable | Coef. | Std.Err. | z | P>\|z\| | [0.025 | 0.975] |
| --- | --- | --- | --- | --- | --- | --- |
| Intercept | 331.868 | 9.193 | 36.099 | 0.000 | 313.850 | 349.887 |
| scenario_id[T.B-2025-04-01] | -39.175 | 1.149 | -34.099 | 0.000 | -41.427 | -36.924 |
| scenario_id[T.C-2025-04-01] | -54.127 | 1.149 | -47.113 | 0.000 | -56.379 | -51.875 |
| scenario_id[T.D-2025-04-01] | -49.900 | 1.149 | -43.434 | 0.000 | -52.152 | -47.649 |
| scenario_id[T.E-2025-04-01] | -68.917 | 1.149 | -59.986 | 0.000 | -71.169 | -66.665 |
| Group Var | 2264.100 | 14.913 |  |  |  |  |

| Variable | Coef. | Std.Err. | z | P>\|z\| | [0.025 | 0.975] |
| --- | --- | --- | --- | --- | --- | --- |
| Intercept | 198.181 | 6.776 | 29.247 | 0.000 | 184.900 | 211.462 |
| scenario_id[T.B-2025-04-01] | -7.045 | 1.086 | -6.485 | 0.000 | -9.175 | -4.916 |
| scenario_id[T.C-2025-04-01] | -22.602 | 1.086 | -20.804 | 0.000 | -24.732 | -20.473 |
| scenario_id[T.D-2025-04-01] | -8.014 | 1.086 | -7.376 | 0.000 | -10.143 | -5.884 |
| scenario_id[T.E-2025-04-01] | -27.131 | 1.086 | -24.972 | 0.000 | -29.260 | -25.001 |
| Group Var | 1223.824 | 8.532 |  |  |  |  |

| Variable | Coef. | Std.Err. | z | P>\|z\| | [0.025 | 0.975] |
| --- | --- | --- | --- | --- | --- | --- |
| Intercept | 133.687 | 4.584 | 29.165 | 0.000 | 124.703 | 142.671 |
| scenario_id[T.B-2025-04-01] | -32.130 | 0.701 | -45.827 | 0.000 | -33.504 | -30.756 |
| scenario_id[T.C-2025-04-01] | -31.525 | 0.701 | -44.963 | 0.000 | -32.899 | -30.150 |
| scenario_id[T.D-2025-04-01] | -41.887 | 0.701 | -59.743 | 0.000 | -43.261 | -40.513 |
| scenario_id[T.E-2025-04-01] | -41.786 | 0.701 | -59.600 | 0.000 | -43.161 | -40.412 |
| Group Var | 560.658 | 6.054 |  |  |  |  |

| Variable | Coef. | Std.Err. | z | P>\|z\| | [0.025 | 0.975] |
| --- | --- | --- | --- | --- | --- | --- |
| Intercept | 361.245 | 7.372 | 49.004 | 0.000 | 346.796 | 375.693 |
| scenario_id[T.B-2025-04-01] | -68.746 | 1.233 | -55.759 | 0.000 | -71.163 | -66.330 |
| scenario_id[T.C-2025-04-01] | -84.234 | 1.233 | -68.321 | 0.000 | -86.650 | -81.817 |
| scenario_id[T.D-2025-04-01] | -88.355 | 1.233 | -71.663 | 0.000 | -90.772 | -85.939 |
| scenario_id[T.E-2025-04-01] | -111.125 | 1.233 | -90.132 | 0.000 | -113.542 | -108.709 |
| Group Var | 1339.540 | 8.917 |  |  |  |  |

| Variable | Coef. | Std.Err. | z | P>\|z\| | [0.025 | 0.975] |
| --- | --- | --- | --- | --- | --- | --- |
| Intercept | 145.168 | 6.788 | 21.387 | 0.000 | 131.864 | 158.472 |
| scenario_id[T.B-2025-04-01] | -8.774 | 1.096 | -8.004 | 0.000 | -10.922 | -6.625 |
| scenario_id[T.C-2025-04-01] | -23.392 | 1.096 | -21.340 | 0.000 | -25.541 | -21.244 |
| scenario_id[T.D-2025-04-01] | -10.294 | 1.096 | -9.391 | 0.000 | -12.442 | -8.145 |
| scenario_id[T.E-2025-04-01] | -29.518 | 1.096 | -26.928 | 0.000 | -31.667 | -27.370 |
| Group Var | 1136.785 | 8.497 |  |  |  |  |

**Table S19.** Mixed effects analysis results for Model 5 over the projection period (April 27, 2025 to April 25, 2026) using Scenario A as baseline, rest of the scenarios (B-E) as fixed effects and location (states) as random effects for both summer and winter leaning locations.
| Variable | Coef. | Std.Err. | z | P>\|z\| | [0.025 | 0.975] |
| --- | --- | --- | --- | --- | --- | --- |
| Intercept | 216.077 | 3.177 | 68.012 | 0.000 | 209.850 | 222.304 |
| scenario_id[T.B-2025-04-01] | -59.972 | 0.859 | -69.805 | 0.000 | -61.656 | -58.288 |
| scenario_id[T.C-2025-04-01] | -60.841 | 0.859 | -70.817 | 0.000 | -62.525 | -59.157 |
| scenario_id[T.D-2025-04-01] | -78.061 | 0.859 | -90.860 | 0.000 | -79.745 | -76.378 |
| scenario_id[T.E-2025-04-01] | -81.607 | 0.859 | -94.987 | 0.000 | -83.291 | -79.923 |
| Group Var | 243.115 | 2.330 |  |  |  |  |

**Table S20.**
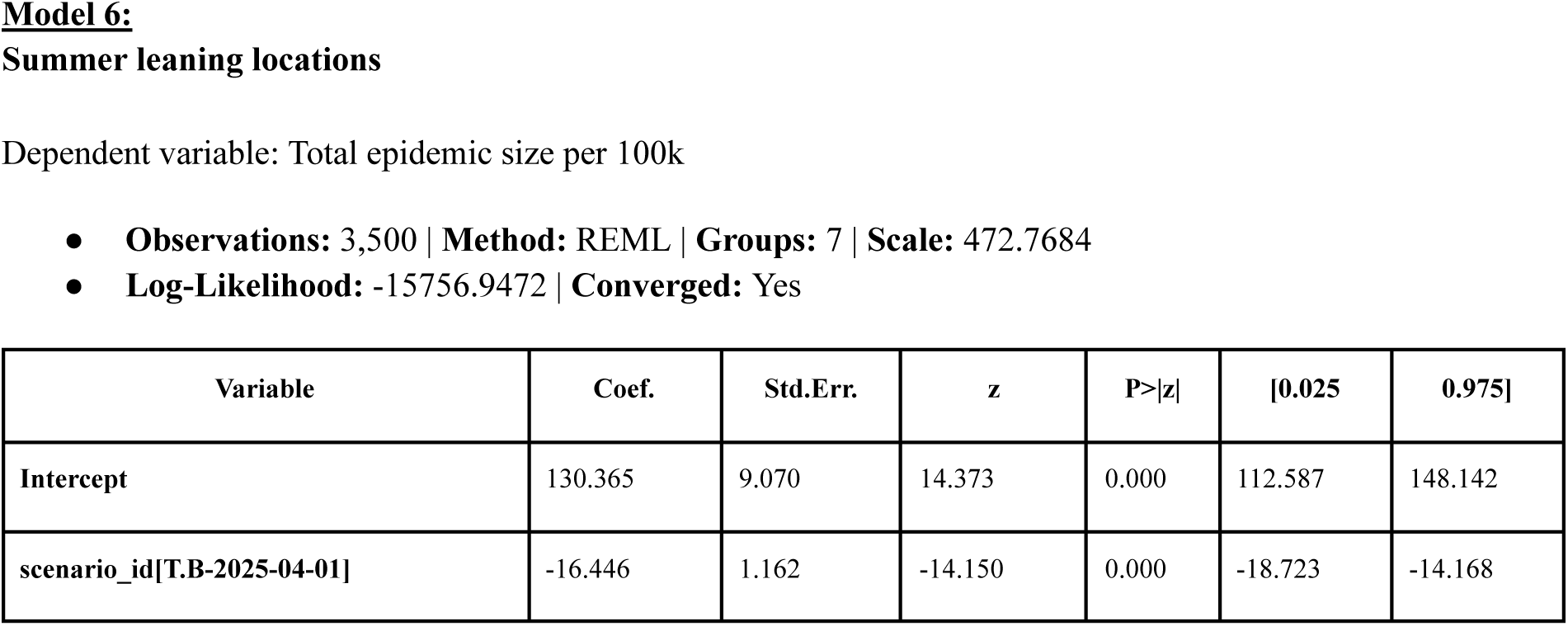
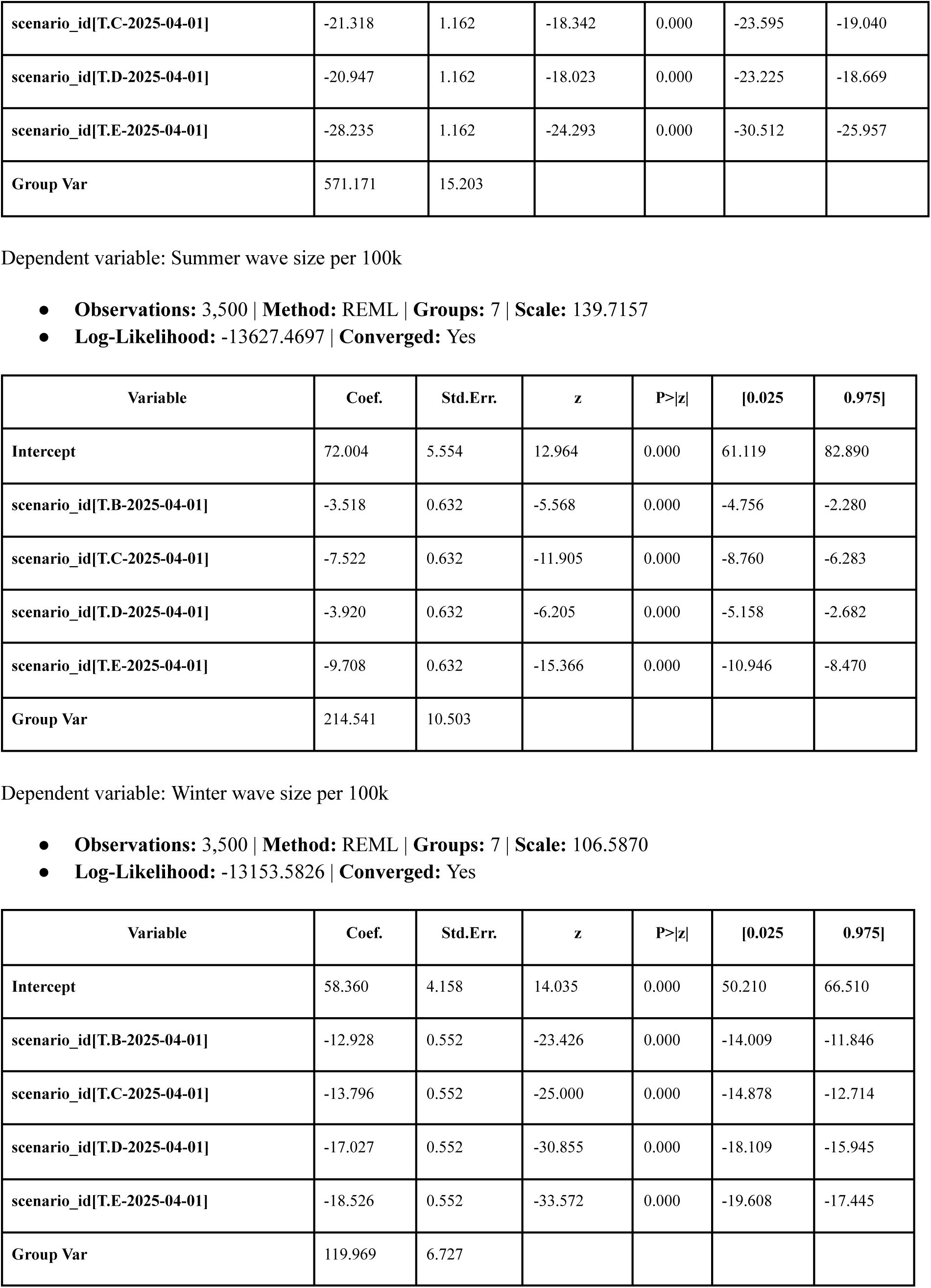
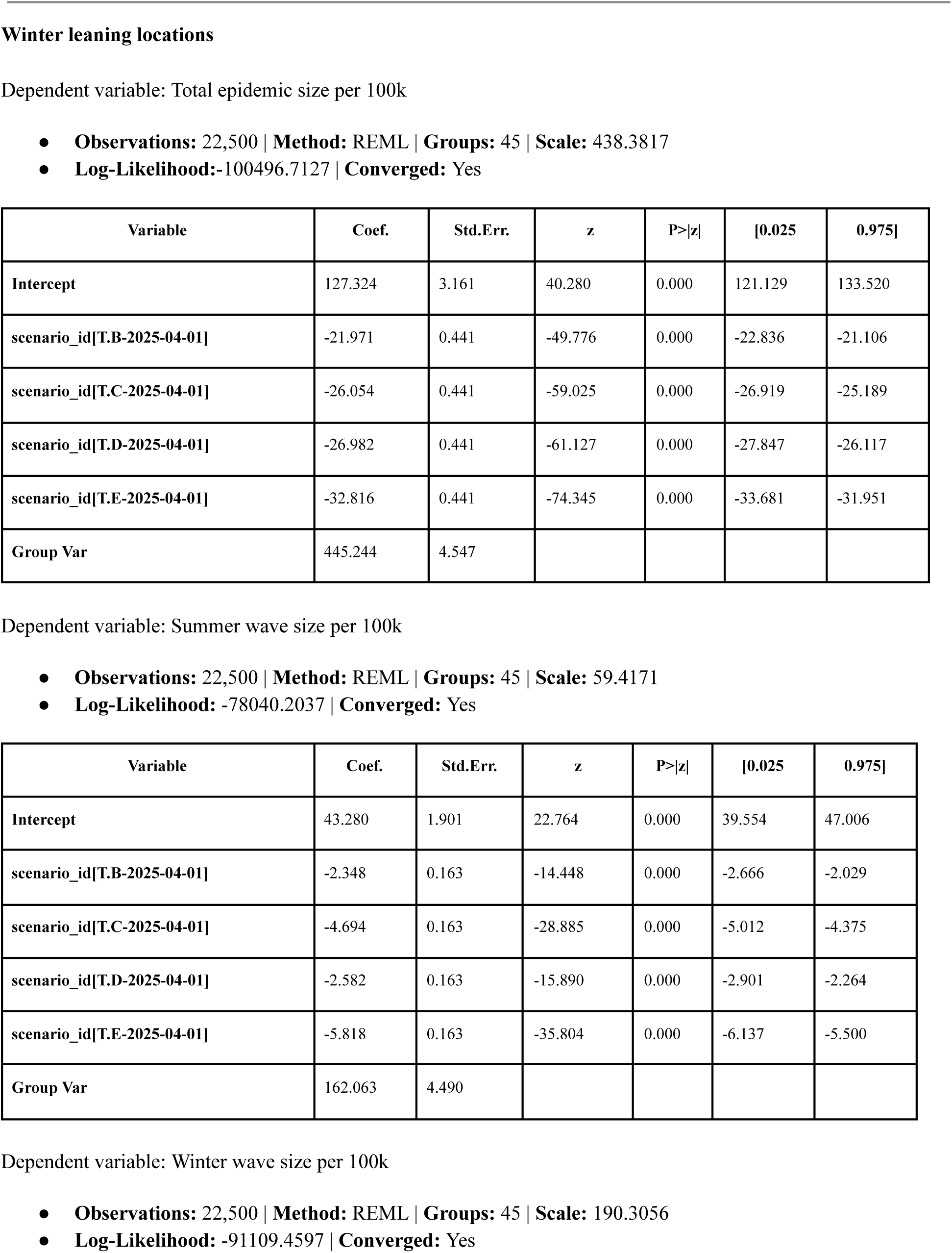
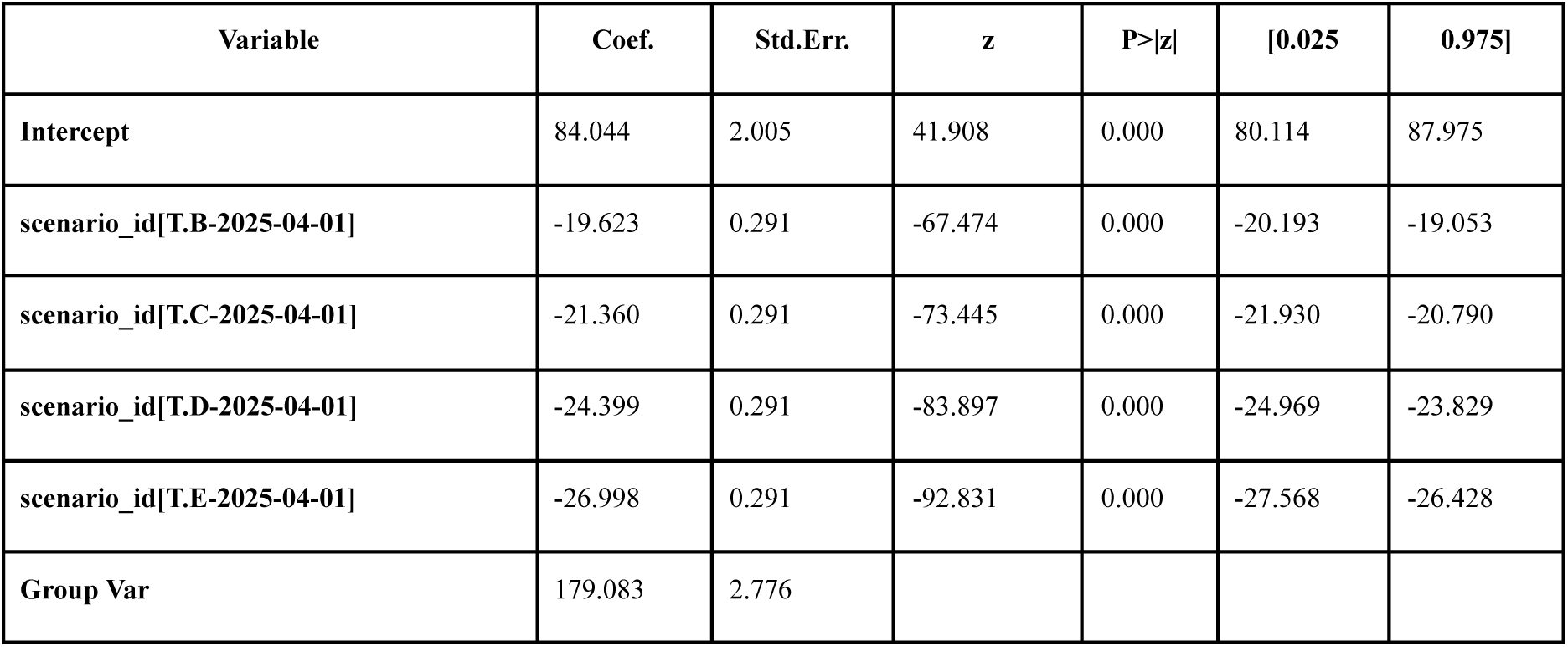
Mixed effects analysis results for Model 6 over the projection period (April 27, 2025 to April 25, 2026) using Scenario A as baseline, rest of the scenarios (B-E) as fixed effects and location (states) as random effects for both summer and winter leaning locations. The three dependent variables considered are: total, summer, and winter wave sizes per 100k population. Absolute values of the entries in the Coef. column for the scenario variables corresponds to the average number of hospitalizations averted per 100k population compared to Scenario A.

| Variable | Coef. | Std.Err. | z | P>\|z\| | [0.025 | 0.975] |
| --- | --- | --- | --- | --- | --- | --- |
| Intercept | 130.365 | 9.070 | 14.373 | 0.000 | 112.587 | 148.142 |
| scenario_id[T.B-2025-04-01] | -16.446 | 1.162 | -14.150 | 0.000 | -18.723 | -14.168 |
| scenario_id[T.C-2025-04-01] | -21.318 | 1.162 | -18.342 | 0.000 | -23.595 | -19.040 |
| scenario_id[T.D-2025-04-01] | -20.947 | 1.162 | -18.023 | 0.000 | -23.225 | -18.669 |
| scenario_id[T.E-2025-04-01] | -28.235 | 1.162 | -24.293 | 0.000 | -30.512 | -25.957 |
| Group Var | 571.171 | 15.203 |  |  |  |  |

| Variable | Coef. | Std.Err. | z | P>\|z\| | [0.025 | 0.975] |
| --- | --- | --- | --- | --- | --- | --- |
| Intercept | 72.004 | 5.554 | 12.964 | 0.000 | 61.119 | 82.890 |
| scenario_id[T.B-2025-04-01] | -3.518 | 0.632 | -5.568 | 0.000 | -4.756 | -2.280 |
| scenario_id[T.C-2025-04-01] | -7.522 | 0.632 | -11.905 | 0.000 | -8.760 | -6.283 |
| scenario_id[T.D-2025-04-01] | -3.920 | 0.632 | -6.205 | 0.000 | -5.158 | -2.682 |
| scenario_id[T.E-2025-04-01] | -9.708 | 0.632 | -15.366 | 0.000 | -10.946 | -8.470 |
| Group Var | 214.541 | 10.503 |  |  |  |  |

| Variable | Coef. | Std.Err. | z | P>\|z\| | [0.025 | 0.975] |
| --- | --- | --- | --- | --- | --- | --- |
| Intercept | 58.360 | 4.158 | 14.035 | 0.000 | 50.210 | 66.510 |
| scenario_id[T.B-2025-04-01] | -12.928 | 0.552 | -23.426 | 0.000 | -14.009 | -11.846 |
| scenario_id[T.C-2025-04-01] | -13.796 | 0.552 | -25.000 | 0.000 | -14.878 | -12.714 |
| scenario_id[T.D-2025-04-01] | -17.027 | 0.552 | -30.855 | 0.000 | -18.109 | -15.945 |
| scenario_id[T.E-2025-04-01] | -18.526 | 0.552 | -33.572 | 0.000 | -19.608 | -17.445 |
| Group Var | 119.969 | 6.727 |  |  |  |  |
| Intercept | 127.324 | 3.161 | 40.280 | 0.000 | 121.129 | 133.520 |
| scenario_id[T.B-2025-04-01] | -21.971 | 0.441 | -49.776 | 0.000 | -22.836 | -21.106 |
| scenario_id[T.C-2025-04-01] | -26.054 | 0.441 | -59.025 | 0.000 | -26.919 | -25.189 |
| scenario_id[T.D-2025-04-01] | -26.982 | 0.441 | -61.127 | 0.000 | -27.847 | -26.117 |
| scenario_id[T.E-2025-04-01] | -32.816 | 0.441 | -74.345 | 0.000 | -33.681 | -31.951 |
| Group Var | 445.244 | 4.547 |  |  |  |  |

| Variable | Coef. | Std.Err. | z | P>\|z\| | [0.025 | 0.975] |
| --- | --- | --- | --- | --- | --- | --- |
| Intercept | 43.280 | 1.901 | 22.764 | 0.000 | 39.554 | 47.006 |
| scenario_id[T.B-2025-04-01] | -2.348 | 0.163 | -14.448 | 0.000 | -2.666 | -2.029 |
| scenario_id[T.C-2025-04-01] | -4.694 | 0.163 | -28.885 | 0.000 | -5.012 | -4.375 |
| scenario_id[T.D-2025-04-01] | -2.582 | 0.163 | -15.890 | 0.000 | -2.901 | -2.264 |
| scenario_id[T.E-2025-04-01] | -5.818 | 0.163 | -35.804 | 0.000 | -6.137 | -5.500 |
| Group Var | 162.063 | 4.490 |  |  |  |  |

**Table S20.** Mixed effects analysis results for Model 6 over the projection period (April 27, 2025 to April 25, 2026) using Scenario A as baseline, rest of the scenarios (B-E) as fixed effects and location (states) as random effects for both summer and winter leaning locations.
| Variable | Coef. | Std.Err. | z | P>\|z\| | [0.025 | 0.975] |
| --- | --- | --- | --- | --- | --- | --- |
| Intercept | 84.044 | 2.005 | 41.908 | 0.000 | 80.114 | 87.975 |
| scenario_id[T.B-2025-04-01] | -19.623 | 0.291 | -67.474 | 0.000 | -20.193 | -19.053 |
| scenario_id[T.C-2025-04-01] | -21.360 | 0.291 | -73.445 | 0.000 | -21.930 | -20.790 |
| scenario_id[T.D-2025-04-01] | -24.399 | 0.291 | -83.897 | 0.000 | -24.969 | -23.829 |
| scenario_id[T.E-2025-04-01] | -26.998 | 0.291 | -92.831 | 0.000 | -27.568 | -26.428 |
| Group Var | 179.083 | 2.776 |  |  |  |  |

**Table S21.**
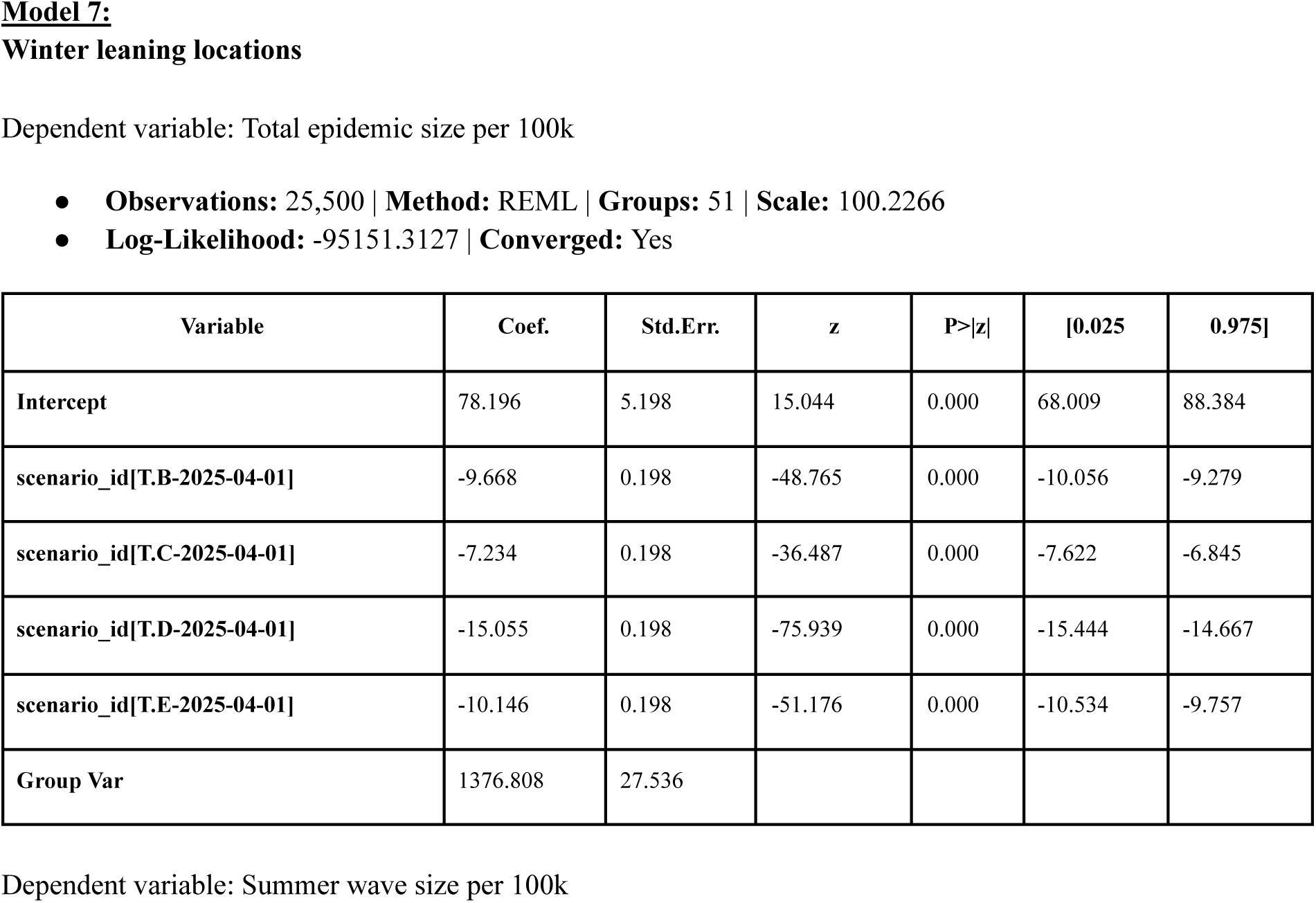
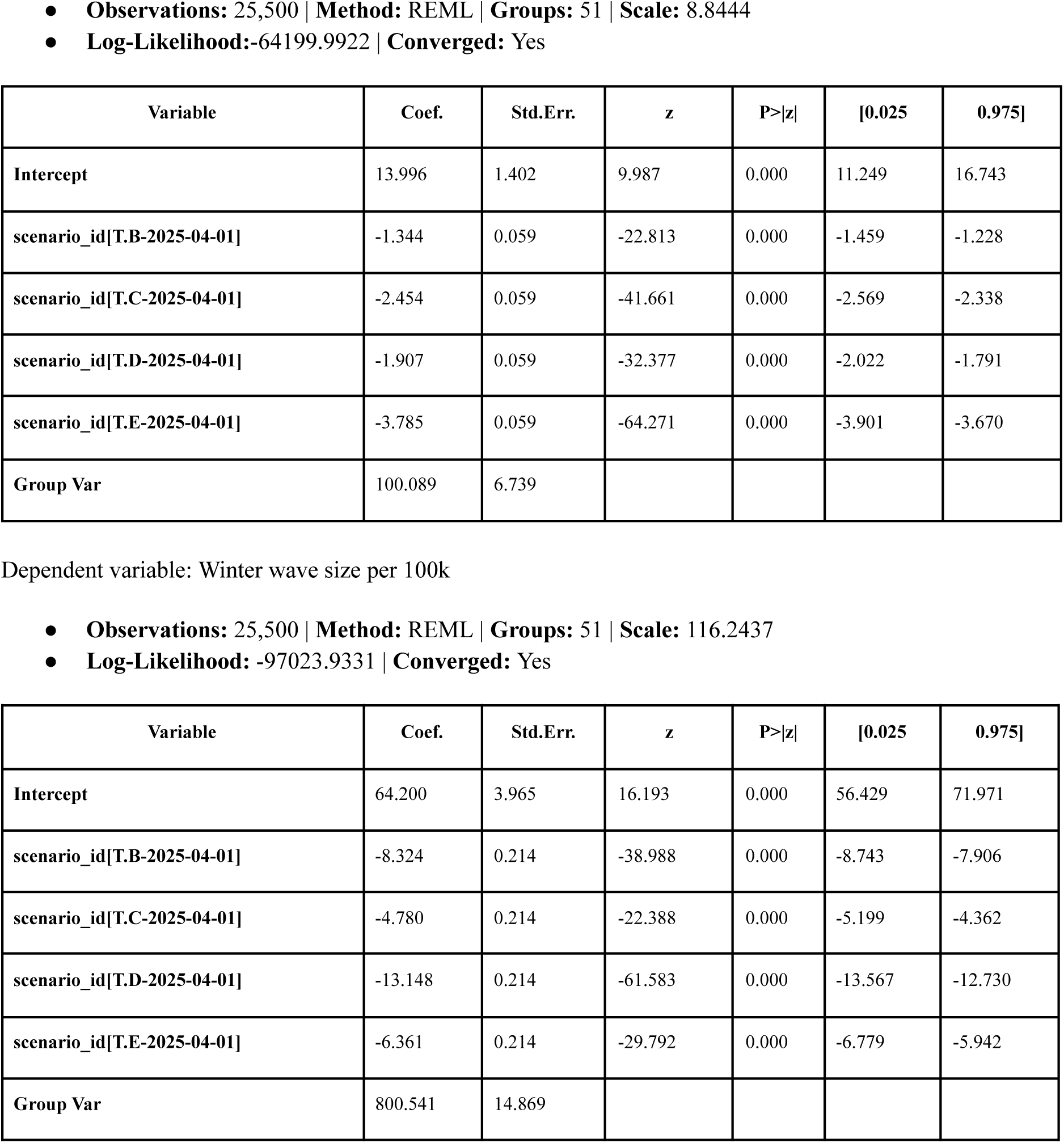
Mixed effects analysis results for Model 7 over the projection period (April 27, 2025 to April 25, 2026) using Scenario A as baseline, rest of the scenarios (B-E) as fixed effects and location (states) as random effects for winter leaning locations (model has no summer leaning locations). The three dependent variables considered are: total, summer, and winter wave sizes per 100k population. Absolute values of the entries in the Coef. column for the scenario variables corresponds to the average number of hospitalizations averted per 100k population compared to Scenario A.

| Variable | Coef. | Std.Err. | z | P>\|z\| | [0.025 | 0.975] |
| --- | --- | --- | --- | --- | --- | --- |
| Intercept | 78.196 | 5.198 | 15.044 | 0.000 | 68.009 | 88.384 |
| scenario_id[T.B-2025-04-01] | -9.668 | 0.198 | -48.765 | 0.000 | -10.056 | -9.279 |
| scenario_id[T.C-2025-04-01] | -7.234 | 0.198 | -36.487 | 0.000 | -7.622 | -6.845 |
| scenario_id[T.D-2025-04-01] | -15.055 | 0.198 | -75.939 | 0.000 | -15.444 | -14.667 |
| scenario_id[T.E-2025-04-01] | -10.146 | 0.198 | -51.176 | 0.000 | -10.534 | -9.757 |
| Group Var | 1376.808 | 27.536 |  |  |  |  |
| Intercept | 13.996 | 1.402 | 9.987 | 0.000 | 11.249 | 16.743 |
| scenario_id[T.B-2025-04-01] | -1.344 | 0.059 | -22.813 | 0.000 | -1.459 | -1.228 |
| scenario_id[T.C-2025-04-01] | -2.454 | 0.059 | -41.661 | 0.000 | -2.569 | -2.338 |
| scenario_id[T.D-2025-04-01] | -1.907 | 0.059 | -32.377 | 0.000 | -2.022 | -1.791 |
| scenario_id[T.E-2025-04-01] | -3.785 | 0.059 | -64.271 | 0.000 | -3.901 | -3.670 |
| Group Var | 100.089 | 6.739 |  |  |  |  |

**Table S21.** Mixed effects analysis results for Model 7 over the projection period (April 27, 2025 to April 25, 2026) using Scenario A as baseline, rest of the scenarios (B-E) as fixed effects and location (states) as random effects for winter leaning locations (model has no summer leaning locations).
| Variable | Coef. | Std.Err. | z | P>\|z\| | [0.025 | 0.975] |
| --- | --- | --- | --- | --- | --- | --- |
| Intercept | 64.200 | 3.965 | 16.193 | 0.000 | 56.429 | 71.971 |
| scenario_id[T.B-2025-04-01] | -8.324 | 0.214 | -38.988 | 0.000 | -8.743 | -7.906 |
| scenario_id[T.C-2025-04-01] | -4.780 | 0.214 | -22.388 | 0.000 | -5.199 | -4.362 |
| scenario_id[T.D-2025-04-01] | -13.148 | 0.214 | -61.583 | 0.000 | -13.567 | -12.730 |
| scenario_id[T.E-2025-04-01] | -6.361 | 0.214 | -29.792 | 0.000 | -6.779 | -5.942 |
| Group Var | 800.541 | 14.869 |  |  |  |  |

**Table S22.**
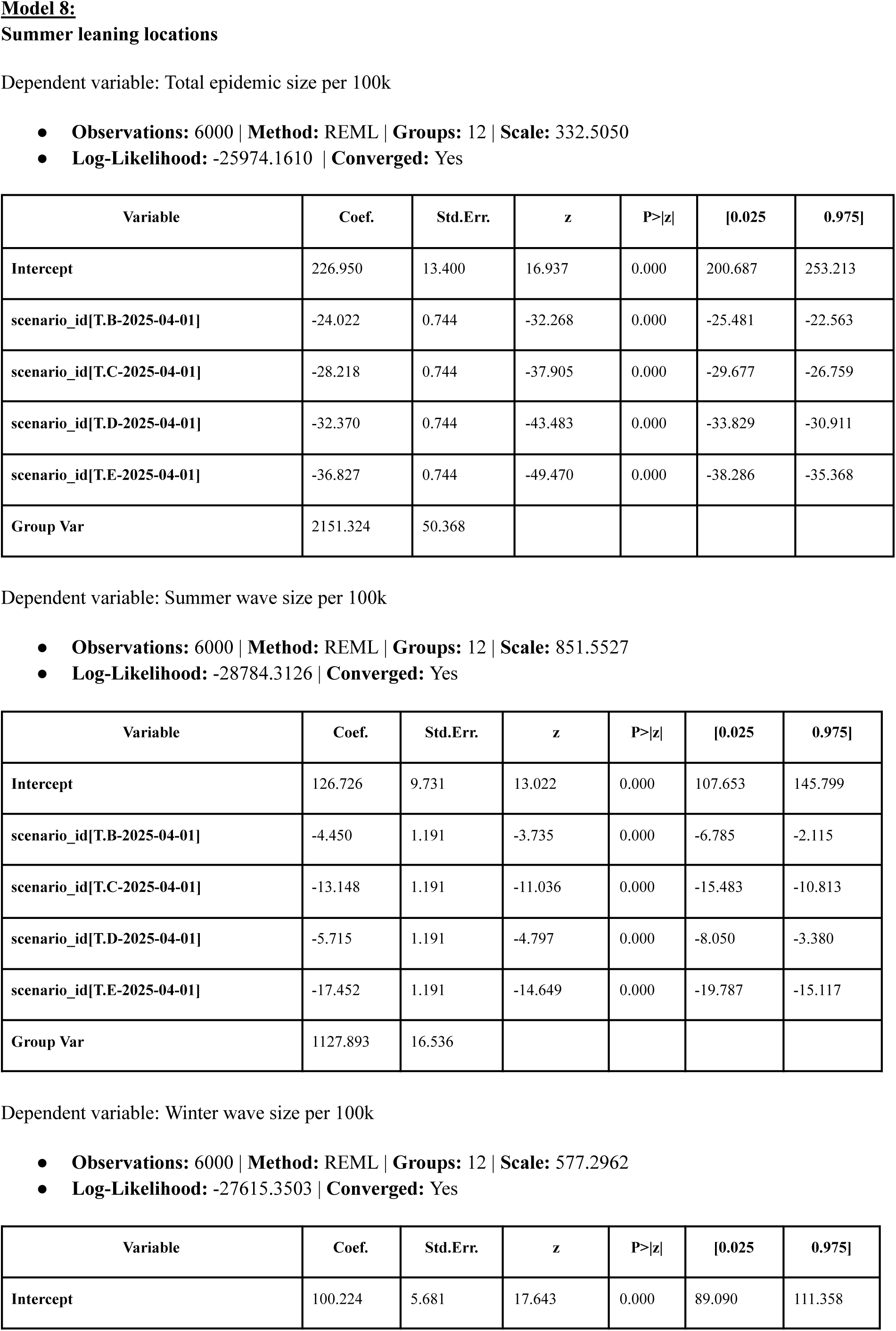
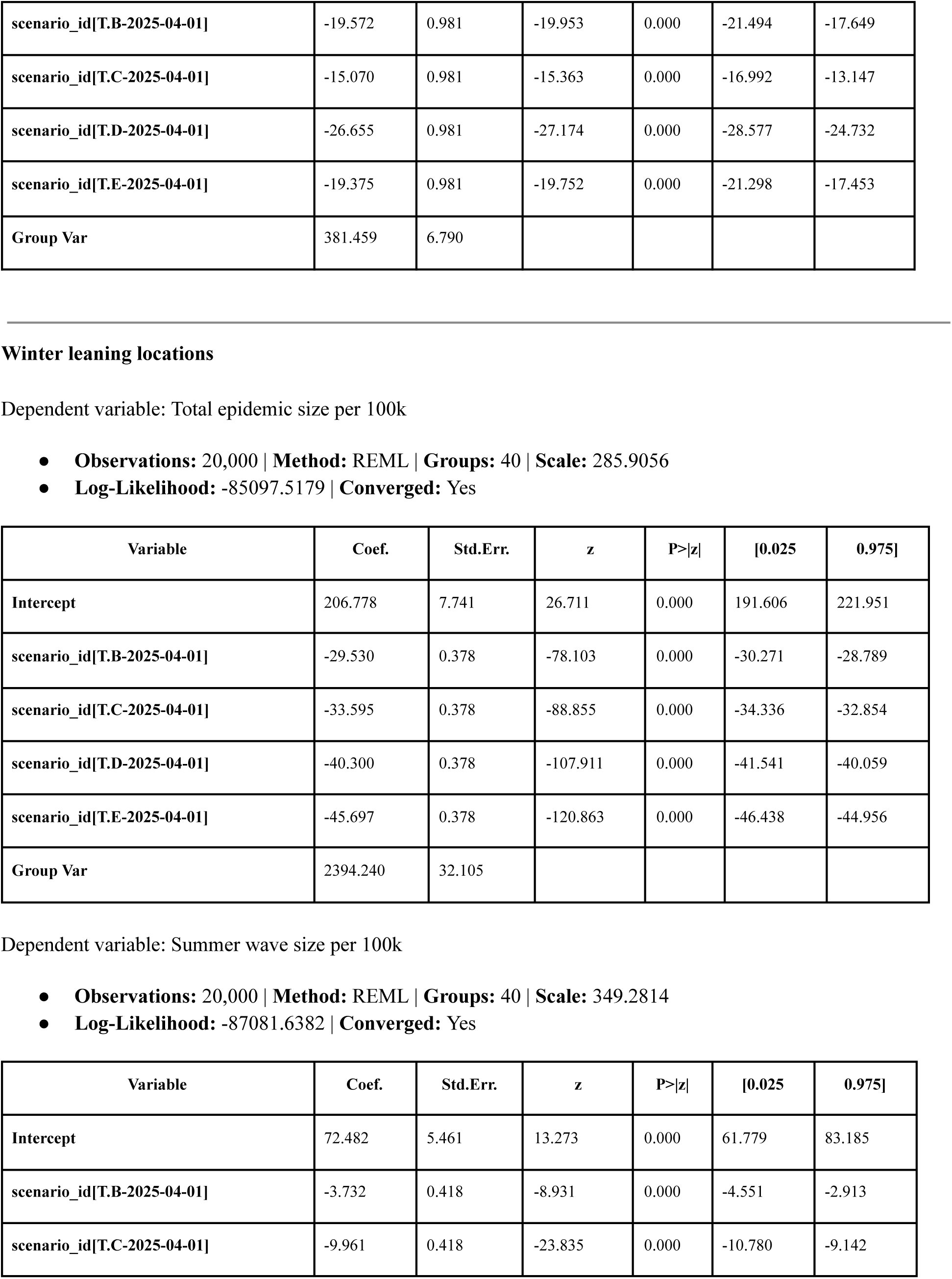
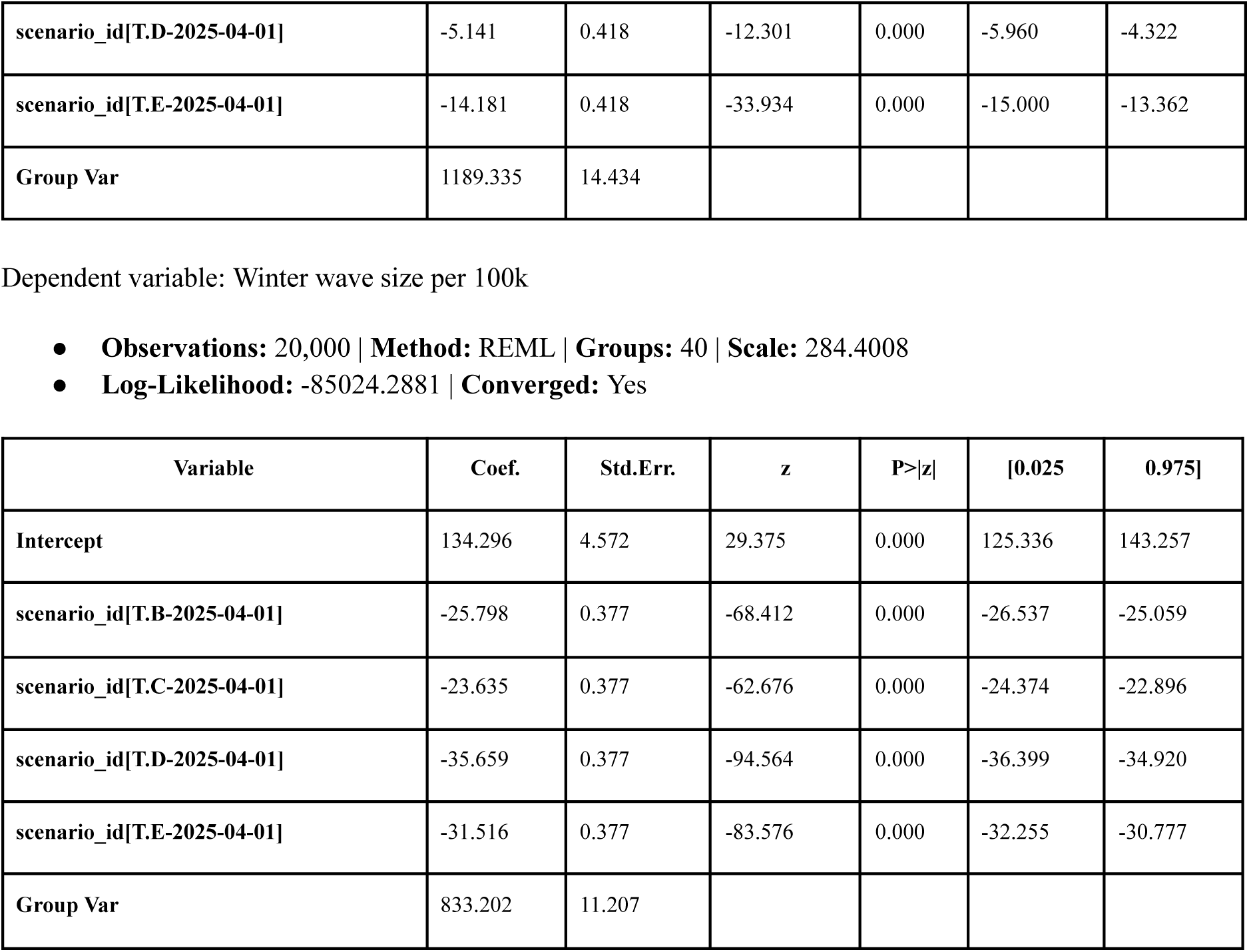
Mixed effects analysis results for Model 8 over the projection period (April 27, 2025 to April 25, 2026) using Scenario A as baseline, rest of the scenarios (B-E) as fixed effects and location (states) as random effects for both summer and winter leaning locations. The three dependent variables considered are: total, summer, and winter wave sizes per 100k population. Absolute values of the entries in the Coef. column for the scenario variables corresponds to the average number of hospitalizations averted per 100k population compared to Scenario A.

### 5 Code availability

All analyses were performed in *Python*^7^ (with additional libraries *NumPy, SciPy*, *statsmodels*, *pandas, matplotlib*) and *R*^8^ (with additional libraries *tidyverse*, *arrow, grid*, *broom*, *MMWRweek*). The analysis code is freely available at https://github.com/midas-network/covid19-scenario-hub_r19_manuscript.

## Notes

### Competing Interest Statement

The authors have declared no competing interest.

### Author Declarations

Study used only simulated data and publicly available data as mentioned in the data availability section.

### Summary of Updates

1. There were errors in the formatting of the abstract which are now fixed. 2. We forgot to add the statement that the study used only simulation data in the author declarations so this has now been added.

